# Information Bottlenecks in Forecasting COVID-19

**DOI:** 10.1101/2024.01.30.24302003

**Authors:** David Gamarnik, Muzhi Ma

## Abstract

Reliable short term and long term forecasting of the number of COVID-19 incidences is a task of clear importance. Numerous attempts for such forecasting have been attempted historically since the onset of the pandemic. While many successful short-term forecasting models have been put forward, predictions for mid-range time intervals (few weeks) and long-range ones (few months to half a year) appeared to be largely inaccurate.

In this paper we investigate systematically the question as to what extend such predictions are even possible given the information available at the times when the predictions are made. We demonstrate that predictions on the daily basis is practically impossible beyond the horizon of 20+ days, and predictions on the weekly basis is similarly impossible beyond the horizon of roughly half a year. We arrive at this conclusion by computing information bottlenecks arising in the dynamics of the COVID-19 pandemic. Such bottlenecks stem from the “memoryless” property of the stochastic dynamical systems describing COVID-19 evolution, specifically from the so-called mixing rate of the system. The mixing rate is then used to gage the rate at which the information used at a time when predictions are made no longer impacts the actual outcomes of the pandemic.

## Introduction and the summary of the results

### Efforts in forecasting COVID-19

Efforts to forecast the number of COVID-19 incidences as well as related metrics such as hospitalization, severity and mortality began pretty much since the beginning of the pandemic in Spring of 2020. Related work includes (*1–56*). The importance of such forecasts needs no justification: knowing the number of incidences can inform health care planners to provision health care supply and treatment accordingly. The models used for predictions ranged widely from some simple statistical estimations to very complex epidemic models based on a multitude of parameters. In order to gage the success level of such predictions, a hub was created which compiled the forecasts put forward by various researchers and organizations https://covid19forecasthub.org. While the models used by forecasters and reported in this hub incorporated a variety of types of data inputs, the number of incidences of COVID-19, mortality and hospitalization were used almost invariably. A large number of models also employed the mobility data as a data input. The hub contains predictions from over 90 research groups. Furthermore, a meta model was constructed which combined the predictions of individual models and a meta study was conducted for evaluating the accuracy of these predictions (*1*).

One of the conclusions of this study, (which perhaps was hardly surprising), was that long term predictions, specifically predictions of the number of incidences of COVID-19 beyond a half a year horizon were quite inaccurate, and could be hardly distinguished from pure guessing. Furthermore, for about one third of the competing research teams, their predictions were less accurate than the so-called baseline model. The baseline model is a very simple model which predicts the metric of interest based on the currently observed numbers. Thus failing to predict better than the baseline implies not even being able to predict whether the metric of interest will *increase* or *decrease* over the time horizon of interest, let alone providing more refined estimates.

Failure to provide accurate predictions for COVID-19 was also well documented in a paper distinctively called “Forecasting for COVID-19 has failed” (*2*). The paper lists a range of instances where predictions put forward by many research teams vastly disagreed with actually observed numbers, sometimes in fact by orders of magnitude.

### Informational limits in long-term forecasting

The mixed success or even the outright failure to conduct accurate predictions of COVID-19 in the long term raises a question as to what extent such predictions are even feasible. In this paper we propose an approach for understanding and estimating the *limits* of predictability of COVID-19 incidences in the long term. Based on our approach we conclude that predicting the number of incidences on the daily basis, namely predicting the number of COVID-19 incidences to occur *x* days from the date when the prediction is being made, is nearly impossible when the time horizon satisfies *x ≥* 10 or *x ≥* 20 (depending on the choice of the model). Similarly predicting on the weekly basis, namely predicting number of incidences to occur during the *x*-th week away from the time when the predictions are made, is nearly impossible when *x ≥* 20 or *x ≥* 30 (again depending on the choice of the model).

Our claims are based on two related methods. These methods are described below briefly, and at a more detailed technical level in the next section. We also discuss why standard statistical “impossibility” results are not applicable in the setting of COVID-19 predictions, forcing us to resort to models more fitting in capturing salient features of the pandemics.

Every data driven predictive method is based on constructing some way of mapping the data available at a time when the prediction is being made, to the # occurrences of the observable of interest, which is predicted to occur. We denote this mapping generically as *F* (*S*_*T*_) = *O*_*T* +*t*_, where *S*_*T*_ is the state (data) collected up to time period *T*, say fatalities and hospitalization data up to date *T, O*_*T* +*t*_ is the observable of interest which is being predicted at a future time *T* + *t*, say the number of COVID-19 incidences, and *t* is the horizon for prediction, say *t* = 8 weeks from the date *T* when the prediction was made.

The two models which we adopt for our study correspond to two types of mappings, which we denote by *F*_*I*_ and *F*_*II*_. Both models are intended to capture as much as possible of the generality of the dynamics of COVID-19 pandemic, including its time dependent aspects as well as its dependence on observable data. The main conclusion we arrive at in studying these models as is follows: when the prediction horizon *t* is large, the dependence of the mapping on data state *S*_*T*_ is almost non-existent, in the sense that for every two hypothetical states *S*_1_ and *S*_2_ describing two hypothetical data inputs feeding the model, the difference *F* (*S*_1_) *− F* (*S*_2_) is practically indistinguishable from zero. Put it differently both models *F*_*I*_ and *F*_*II*_ of the evolving dynamics of the pandemic have the property that the predictions being made for a long term horizon will be nearly the *same*, regardless of the data used to populate the model. The statistical properties of the observable *O* of interest for the time for which the predictions are made are therefore almost *entirely* an artifact of noise, which accumulates between the time of prediction and the time for which the prediction is made. This is an important conclusion which explains the informational barriers for conducting meaningful long-term predictions. The data incorporated into a predictive model is used to make the predictions as accurate as possible. Naturally then it is paramount that the predictions depends on the data input in a significant way, since otherwise, the overall predictive power of the data is negligible.

### Two models of the evolution of the pandemics

The first of the two aforementioned models giving rise to the mappings *F*_*I*_ is a Markov chain model, which is a canonical choice for modeling uncertain quantities evolving in time in some stochastically dependent way. We use historical data to estimate the transition rates of the Markov chain. From the estimated transition rates of the chain we obtain an estimation of the associated spectral gap *λ*_2_ *<* 1 of the chain (see the next section for details). Standard theory of Markov chains postulates that *λ*_2_ is rate of the memory loss of a chain (the so-called mixing rate), meaning that the state of the chain at time 0 impacts the state of the chain at time *t* by magnitude which is at most order 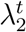. In particular, the impact degrades at a geometric rate as a function of *t*. This loss of memory property of Markov chains (one of its most basic and celebrated properties) thus creates a sort of information bottleneck in our ability to predict the state of the chain at time *T* + *t* given the current state at time *T*, when the value of 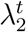 is very small. Based on our estimations we discover that the values of 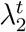 for the time periods *t* corresponding to long term predictions, are indeed very close to zero, and thus the Markov chain describing the dynamics of the pandemic indeed exhibits of almost entire loss of memory.

The spectral gap based reasoning is just an indirect indication of the memory loss. We have also computed it directly as follows. We have computed the impact of the current states *X*_*T*_ on the probability tails, namely, how much the state of the chain at the time of observation (for example the currently observed rates of incidences, hospitalization, etc) impacts the likelihood that the number of incidences *t* days in future will be at least say *x*. Consistently with the spectral gap based estimations, we have found that the state values *X*_*T*_ at the time of observation hardly impact the probability tails beyond the time horizon for predictions. In summary, the Markov chain based model of the evolution of the pandemic exhibits a memory loss making statistical predictions beyond a certain time horizon impossible.

While we believe that Markov chain based model of the evolution of the COVID-19 pandemics is justifiable (more on this in the next section), it suffers from one important limitation: the time homogeneity of the underlying chains. Namely, in our assumptions, the likelihood (probability) of moving from state *X*_*t*_ = *s* to state *X*_*t*+1_ = *s*′ in one step of the chain’s transition depends on the state *s*, but not on the time *t*. This is admittedly objectionable, since presumably the dynamics of the pandemics may be influenced strongly by additional factors not incorporated by states and which are time dependent. These might include season and season related factors such as school times, holidays, temperature, etc.

Our second model addresses these limitations. We consider a non-stationary Markov chain for which the transition rates are possibly time dependent (with the idea that the unobserved latent factors impact transition rates at each time *t*). Non-stationarity presents a serious parameter estimation challenge. For each time period *t* we have only one observed transition *X*_*t*_ *→ X*_*t*+1_ rendering statistical estimation of transition likelihoods for each time period impossible, unless some additional structural and regularization assumptions on the chain are adopted, and this is what we do. Specifically, we adopt the assumption that our Markov chain is a linear time dependent (aka inhomogeneous) auto-regressive process regularized by the assumption that the parameters of the process evolve in some controlled way. We elaborate on this in the next section. We estimate the parameters of this process using the least squares method. The estimated parameters are then used to populate the second model *F*_*II*_. The statistical soundness of this approach based on the computational learning theory, specifically the theory of Vapnik-Chervonenkis dimension, can be found in (*57*). We then use our model to assess the sensitivity of *F*_*II*_ on its argument *S*_*T*_. Once again we discover that the sensitivity of *F*_*II*_ on its initial data condition is very minimal and degrades rapidly as a function of *t*. The future values of observables *O*_*T* +*t*_ = *F*_*II*_(*S*_*T*_) depend very minimally on the current state *S*_*T*_, namely the data used to produce the forecasts.

### Generalizations and alternative methods for establishing limits of predictability

There exists a rich plethora of statistical/information theory based methods for establishing limits for data driven statistical predictability. The methods roughly evolve around two related concepts called mini-max theory in the field of statistics (*58*) and Fano’s inequality in the information theory (*59*). Both methods however are premised on data exhibiting independent and identical distribution (i.i.d.), the property which profoundly is not the case in our setting. The data used in the predictive models discussed in (*1*) as well as in our setting, such as incidence, hospitalization, mortality and mobility, all exhibit auto-correlations. Arguably, one then could try to use time series type models to study non-predictability, such as auto-regressive processes – a standard model in the field of econometrics, which exhibits auto-correlation. The problem with standard auto-regressive models is the time homogeneity of the parameters, which we wish to avoid. Thus we allowed for our second model to exhibit the time dependence of the parameters, but in a regularized way, effectively giving a rise to an inhomogeneous auto-regressive process. This is essentially our model *F*_*II*_ discussed above.

It is entirely possible that more general models can be used for establishing the limits of predictability, such as, for example, models based on latent (hidden) variables, aka Hidden Markov Model and related models. Even broader, one could try to elicit the help of general dynamical systems and using chaos type properties to study the limits of predictability. Such limits are well-known for example in the context of weather prediction models based on Lorenzian dynamics. The relevance of these and related models for predictability of COVID-19 is an interesting question for further research.

## Methods and models

We now turn to the detailed description of our two models *F*_*I*_ and *F*_*II*_ as well as the data we use to populate the parameters of these models. We begin with the data description.

### Data

Consistently with the majority of models analyzed in (*1*) we use four types of data: Incidence, Mortality, Hospitalization and Mobility. This data was retrieved from the same source COVID-19 Forecast Hub as was used in (*1*). The data spans time period beginning with March 29, 2020 and ending April 9, 2022, with a total of 742 days (i.e. 106 weeks). A summary statistics of the data on daily and weekly basis is provided in Table 1-2.

**Table 1:** Statistics of Daily Data.

| Data | Mean | Median | Std | Max | Min |
| --- | --- | --- | --- | --- | --- |
| Incident Cases | 108180.3 | 59551.0 | 148230.9 | 1383795 | 4236 |
| Incident Deaths | 1324.1 | 1046.0 | 962.9 | 4431 | 8 |
| Incident Hospitalizations | 6474.6 | 5276.0 | 5164.3 | 23479 | 0 |
| Mobility Rate | 7.7 | 7.0 | 4.7 | 27 | -2 |

**Table 2:** Statistics of Weekly Data.

| Data | Mean | Median | Std | Max | Min |
| --- | --- | --- | --- | --- | --- |
| Incident Cases | 757261.8 | 460875.0 | 952651.2 | 5654028 | 81687 |
| Incident Deaths | 9268.5 | 7981.5 | 5315.7 | 23447 | 1583 |
| Incident Hospitalizations | 45322.5 | 36283.0 | 35971.1 | 154002 | 16 |
| Mobility Rate | 8.24 | 7.8 | 3.7 | 20.2 | 4.0 |

### Number of incidents and mortality

During the COVID-19 pandemic in the US, data on cases and deaths were collected by state and local governmental health agencies and aggregated into standardized, sharable formats by third-party data tracking systems. Early in the pandemic, the Johns Hopkins Center for Systems Science and Engineering (CSSE) developed a publicly available data tracking system and dashboard. CSSE collected daily data on cumulative reported cases at the county, state, territorial, and national levels and made these data available in a standardized format. Daily incident cases and mortality were inferred from this time-series as the difference in successive reports of cumulative cases, see Figures 1-4. Weekly values are defined and aggregated based on daily totals from Sunday through Saturday, according to the standard definition of epidemiological weeks used by the CDC, see Figures 25-28 in Supplementary materials.

**Figure 1.**
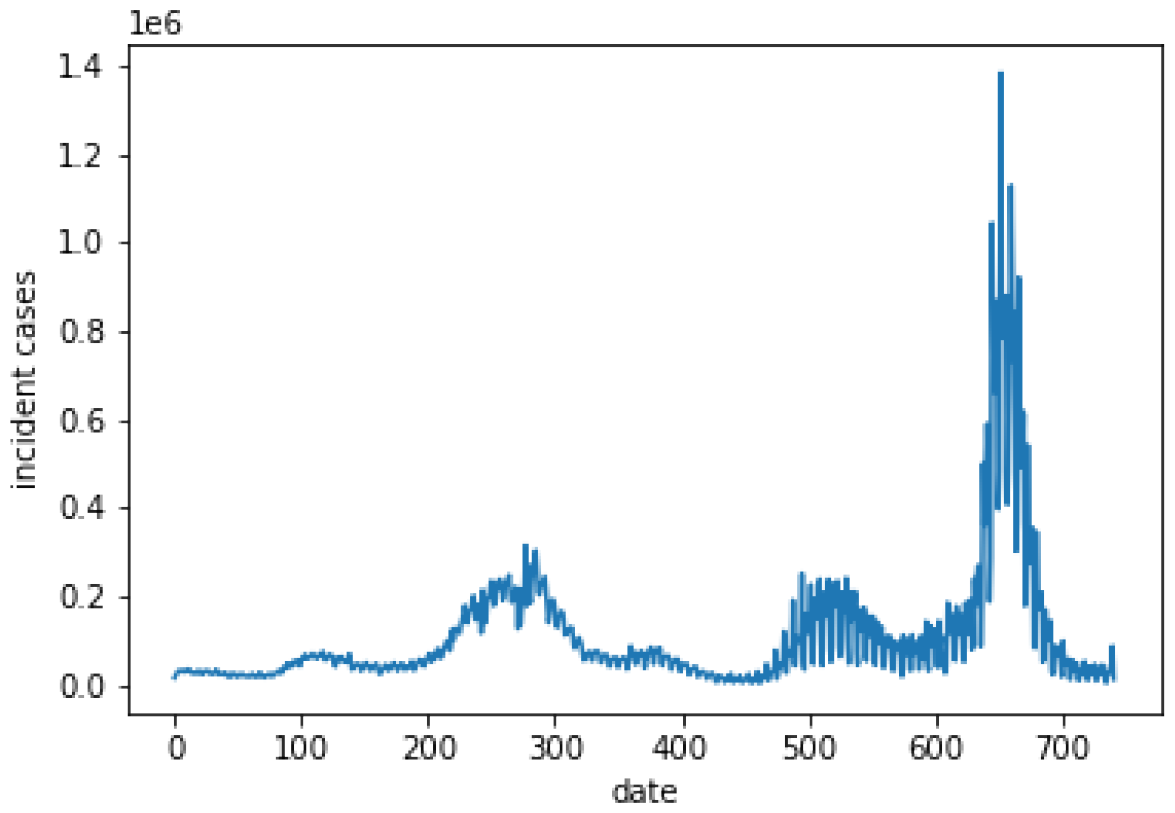
Time Series of Daily Incident Cases.

**Figure 2.**
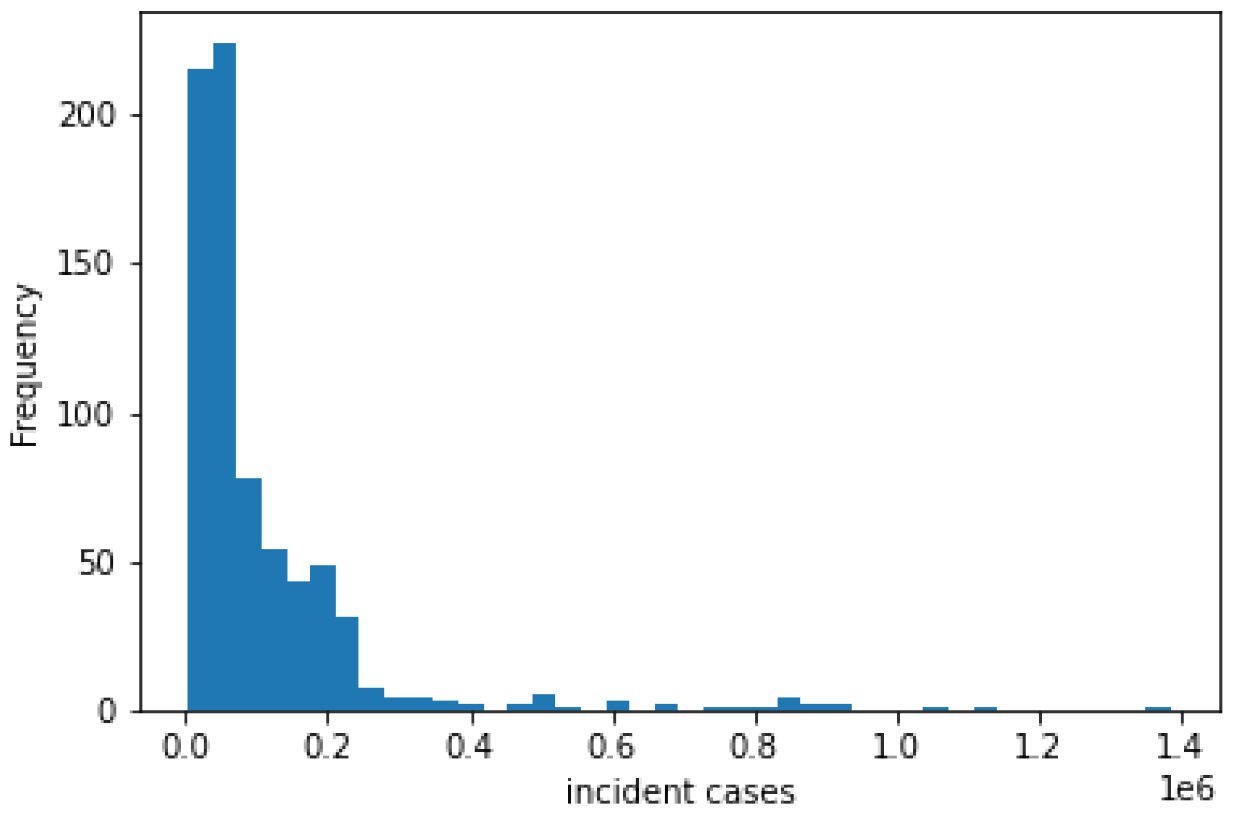
Distribution of Daily Incident Cases.

**Figure 3.**
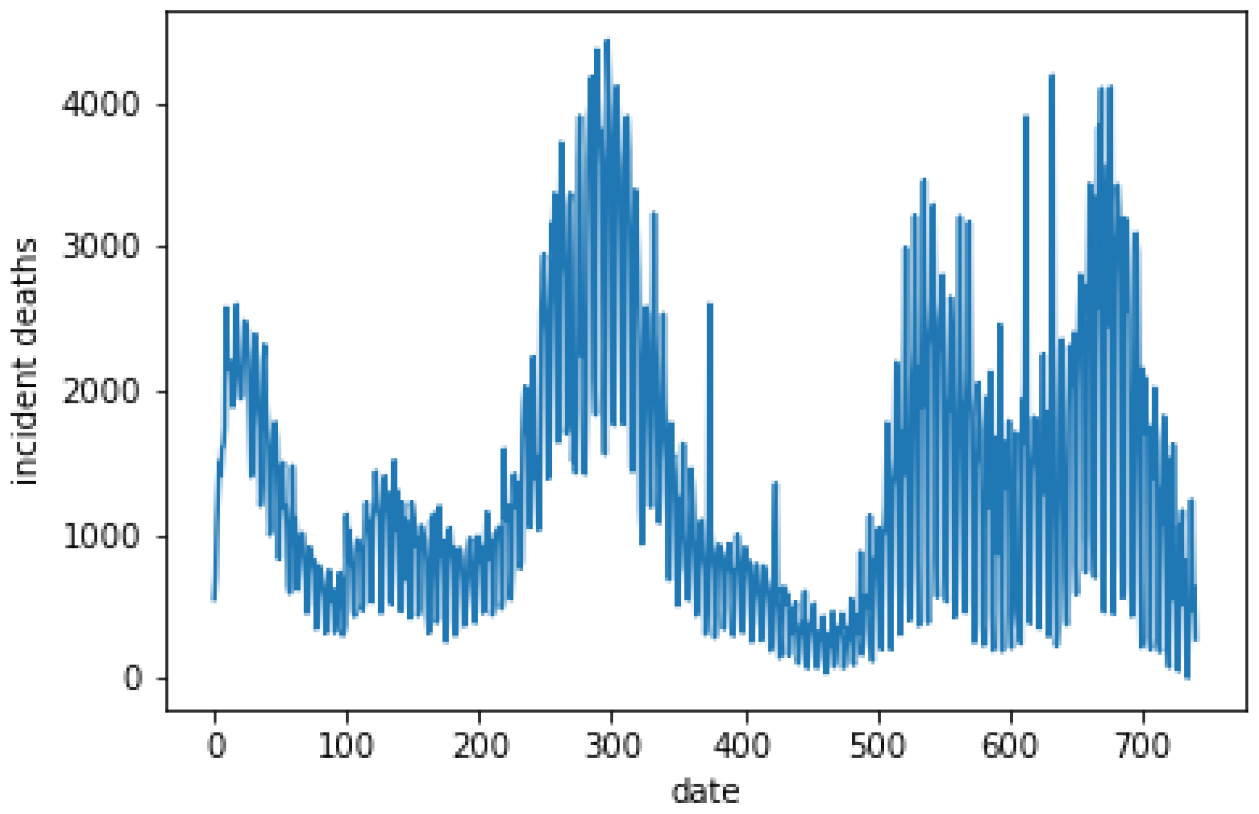
Time Series of Daily Incident Deaths.

**Figure 4.**
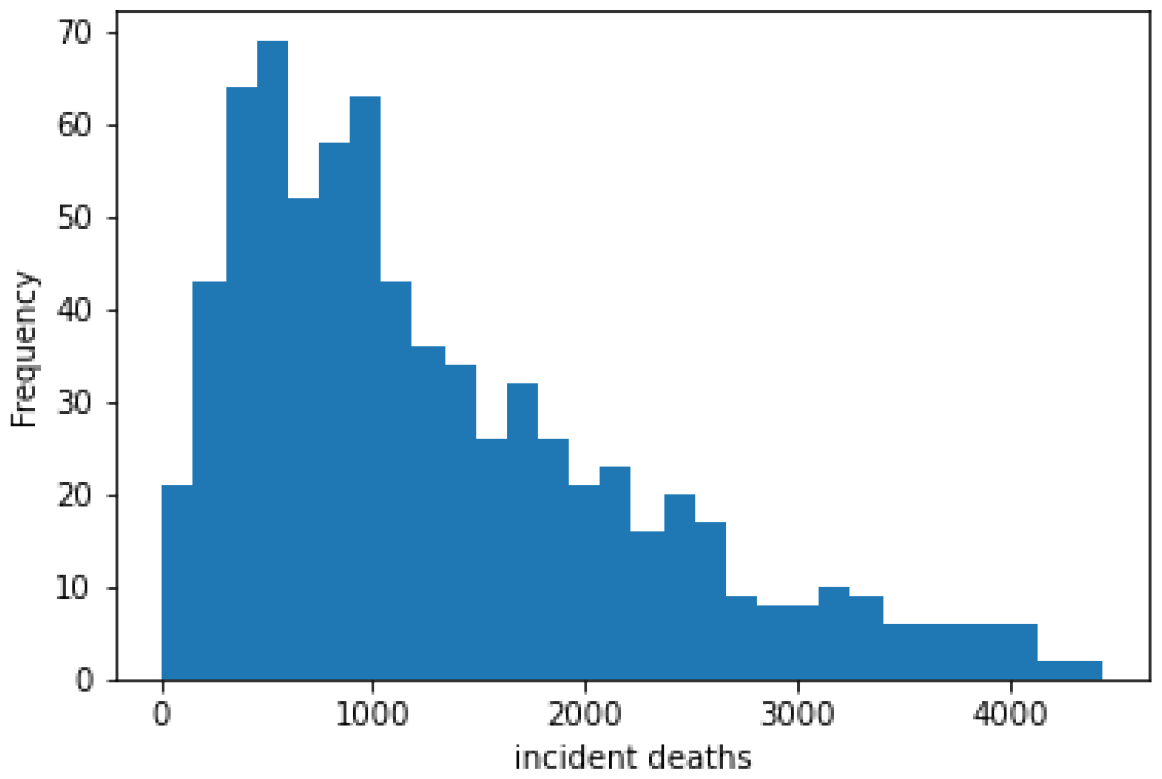
Distribution of Daily Incident Deaths.

### Hospitalizations

CDC records daily hospitalization data through COVID Tracking Project. The dataset indicates the number of newly-admitted COVID-19 patients across the US each day. The patients include both adult and pediatric patients. see Figures 29-32 in Supplementary materials.

### Mobility

During the pandemic, Google collected mobility information from its app users. The daily mobility rate reports how much the length of stay at residential area each day changes compared to a baseline. The baseline is the median value, for the corresponding day of the week, during the 5-week period Jan 3 - Feb 6, 2020. The data represent the percentage of change compared to the baseline. The weekly mobility rate is defined as the daily average mobility rate within a week. See Figures 33-36 in Supplementary materials.

We denote by *I*_*t*_, *D*_*t*_, *H*_*t*_, *M*_*t*_ the number of incidences, mortality, hospitalization and mobility rate observed on day *t*, respectively, as described above. Day *t* = 0 corresponds to March 29, 2020 and the last day *t* = *T* corresponds to April 9, 2022. Thus *T* = 742. We use a similar notation for data on weekly basis. That is, for example *I*_*t*_, *t ∈* [0, *T*] is the number of incidences which occurred on week *t* of the pandemic, where *T* is now 106.

## Models

We now turn to description of the two models under the discussion.

### Model I. Homogeneous Markov chain

The first model *F*_*I*_ is based on a Markov chain description of the processes *I*_*t*_, *D*_*t*_, *H*_*t*_, *M*_*t*_, *t ∈* [0, *T*] where each individual process is postulated to be a Markov chain. We illustrate the idea only for the incidences process *I*_*t*_. The treatment of the remaining three processes is similar. We perform a discretization of the observable *I*_*t*_ in order to have a non-trivial number of observations per transition in our chain. This is done as follows. Let *I*_min_ = min_*t∈*[0,*T*]_ *I*_*t*_ and *I*_max_ = max_*t∈*[0,*T*]_ *I*_*t*_ denote the smallest and the largest values of incidences per day observed historically in the described period. As it turns out *I*_min_ = 4236 and *I*_max_ = 138795, see Table 1. We fix a parameter *n* which describes the discretization level (which we will vary), and split the interval [*I*_min_, *I*_max_] into *n* equal size subintervals *J*_1_, *J*_2_, …, *J*_*n*_, so that *J*_*i*_ = [*I*_min_ + (*i −* 1)(*I*_max_ *− I*_min_)*/n, I*_min_ + *i*(*I*_max_ *− I*_min_)*/n*] for *I* = 1, …, *n*. We declare *I*_*t*_ to be in state *i* at time *t* if *I*_*t*_ belongs to the interval *J*_*i*_ at time *t*. We consider a Markov chain *Ī*_*t*_ on the state space 1, 2, …, *n*. This Markov chain postulates the dynamics of the stochastic process (*I*_*t*_, *t ∈* [0, *T*]) or the process (*Ī*_*t*_, *t ∈* [0, *T*]) to be exact. The transition matrix *P* = (*P*_*ij*_, 1 *≤ i, j ≤ n*) is estimated from the data in a straightforward way: the value *P*_*ij*_ is estimated as a fraction of days *t ∈* [0, *T*] such that *I*_*t*_ *∈ J*_*i*_ (i.e. *Ī*_*t*_ = *i*) and *I*_*t*+1_ *∈ J*_*j*_ (i.e. *Ī*_*t*+1_ = *j*). Namely, it is the number of days such that on this day the number of incidences was in the interval *I*_*i*_ and the number of incidence on the next day was in the interval *I*_*j*_.

We a consider the same model on a weekly basis. Here *I*_*t*_ corresponds to the number of incidences which occurred in week *t* starting from the pandemic (the first week being the week of March 29, 2020 and last week being April 9, 2022). The advantage of analyzing the weekly data is the absence of the day of the week (weekday vs weekend) heterogeneity. The disadvantage is the reduced data size (106 vs 742).

There is a natural statistical trade-off of making *n* too small vs too large. For smaller values of *n* the approximation implied by the rounding the values to the end points of each interval can be potentially significant. The upside though is a larger number of observations per each pair of states *i, j*. Conversely, for large *n*, the interval approximation is more granular, but the number of observations per each pair *i, j* might not be significant enough. For the daily incidences process we vary *n* in the range *n* = 2, 3, …, 10 and for the weekly incidences we vary *n* in the range *n* = 2, …, 5. We observe a fair amount of insensitivity of our results to the choice of the discretization level *n*, as we report in the next section.

Once our *n × n* matrix transition matrix *P* is estimated, we compute its spectral radius *λ*_2_(*P*) *<* 1 which is the second largest eigenvalue. The values of *λ*_2_(*P*) for daily and weekly data for processes *I*_*t*_, *D*_*t*_, *H*_*t*_, *M*_*t*_ and their implications are reported in the next section.

The spectral radius *λ*_2_ provides an upper bound on the memory loss, specifically the rate of convergence to stationarity. More specifically, denote by *π* the (unique) stationary distribution of our chain *P*, that is the unique probability vector solving *π*^*T*^ *P* = *π*^*T*^. The uniqueness was verified directly by computing *π* itself which is straightforward to do. Standard theory of Markov chain postulates that the so-called total variation distance from the transient distribution to the stationary distribution is upper bounded in terms of *λ*_2_ is follows. In general, given two discrete probability distributions *μ*_*i*_, *ν*_*i*_ supported on the set *i* = 1, 2, …, *N* the total variation distance between them is defined as Σ_*i*_ |*μ*_*i*_ *− ν*_*i*_| (here we drop a factor 1*/*2 commonly used in this definition for convenience). Let *Ī*_*t*_ be the Markov chain *P* on state space 1, …, *n* constructed above. Then, for some problem independent constant *C*, the following holds for the total variation distance between the transient distribution and the stationary distribution 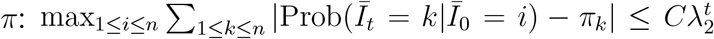. Here Prob(*A*|*B*) denotes the probability of the event *A* conditioned on the event *B*. Namely, the maximum over total variation distances is upper bounded by 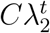 where the maximum is taken over all possible initial states *i*. We denote this maximum total variation value by *TV* (*t*) (step *t* maximum total variation distance). In light of the fact that the chain *P* was explicitly constructed from which the stationary vector *π* was easy to compute, we also computed *TV* (*t*) directly for various time horizons *t*.

Additionally, we have computed directly the following metric *TV*_2_(*t*) = max_1≤*i,j*≤*n*_ Σ_1*≤k≤n*_ |Prob(*Ī*_*t*_ = *k*| *Ī*_0_ = *i*) *−* Prob(*Ī*_*t*_ = *k*| *Ī*_0_ = *j*)|. This measure is trivially at most twice *TV* (*t*), but is somewhat more meaningful in our context. Indeed, this metric *TV*_2_ measures directly the following memory sensitivity property. Prob(*Ī*_*t*_ = *k*| *Ī*_0_ = *i*) is the likelihood that the chain is in state *k* at time *t*, given the information that it is in state *i* now (time zero). The Metric *TV*_2_ then measures the sensitivity of this likelihood to the initial information (current state). If *TV*_2_ is small it implies that the current state has a very small impact on this likelihood, and thus has a very limited predictive power. We report *TV*_2_(*t*) for a range of values of *t* in the next section as well. As we will see, this value drops to very small values when the time horizon *t* is large, specifically less than 1% for *t* at least 40 days and 40 weeks, (depending on whether *I*_*t*_ corresponds to daily or weekly observations).

Finally, we compute and report estimation of the tail events and in particular the sensitivity of the tail events to the information available at the time when the forecasting is being made. Given a threshold value *θ* we consider the probability Prob(*Ī*_*t*_ *≥ θ*| *Ī*_0_ = *i*). Namely, the likelihood of observing at least *θ* incidences of COVID-19 during the time period *t*, when the current incidence level is *Ī*_0_ = *i*. To measure the sensitivity, we introduce *TV* (*t, θ*) = max_1*≤i,j≤n*_ |Prob(*Ī*_*t*_ *≥ θ*| *Ī*_0_ = *i*) *−* Prob(*Ī*_*t*_ *≥ θ*| *Ī*_0_ = *j*)|. We estimate this sensitivity measure for threshold *θ* corresponding to a fraction of the largest historically observed numbers, namely *θ* = *ρMax*(*I*) where *Max*(*I*) is the largest observed incidences (per day or per week) over the entire period [0, *T*], and *ρ* is a fixed percentile level (such as say *ρ* = 0.7 or *ρ* = 0.8). In other words, *TV* (*t, θ*) measures the extent to which the likelihood of observing a spike in the incidences (at least *ρ*-fraction of maximum) depends on the current level of incidences. We estimate this measures both for incidences *I*_*t*_ and mortality *D*_*t*_. In this case the rate of the information loss is even stronger. The value less than 1% is achieved within 10 to 25 days and weeks (for both aggregation levels).

### Model II. Heterogeneous regularized auto-regressive process

As discussed earlier, the Model I suffers from important modeling limitations. First, it postulates the dynamics of the four observables as a homogeneous Markov chain, thus ignoring potential heterogeneity associated with various time dependent events, such as seasonality, government intervention, etc. The second source of limitation is that each of the processes *Ī*_*t*_, 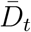 is considered in isolation. Thus for example the value of mortality 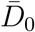 at the observation time *t* = 0 has no impact on predicting the incidence level *Ī*_*t*_ at a future time *t*, and vice verse. As such, our conclusions about the loss of information and the ensuing forecasting limitations are no more than indicative of those.

We now turn to a far richer model which fixes both of these isssues. We call our model heterogeneous regularized auto-regressive process. Introducing a shorthand notation for the four dimensional observable *X*_*t*_ = (*I*_*t*_, *D*_*t*_, *H*_*t*_, *M*_*t*_), *t ∈* [0, *T*], we now postulate the dynamics underlying the stochastic law of the process *X*_*t*_. First, since the processes *I, D, H, M* evolve on a different scale, and, furthermore, the units of these four observables are not identical, we introduce a unit free version of this process, by considering instead the processes 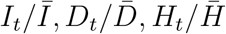 and 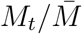, where for each observable *O*_*t*_, 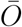 denotes simply the average of this observable over the entire time period *T*. We then define *X*_*t*_ to be instead the unit free version of the same process: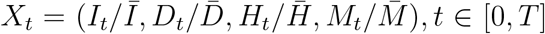, *t ∈* [0, *T*]. We postulate that the dynamics of this process *X*_*t*_ is described by a probabilistic law *X*_*t*+1_ = *F*_*t*_(*X*_*t*_) + *ϵ*_*t*_, where *F*_*t*_ is an (importantly) time-dependent law which translates the observables at time *t* to the observables at time *t* + 1 and *E*_*t*_ is a zero mean idiosyncratic noise. We specifically postulate a linear model of the form *X*_*t*+1_ = *A*_*t*_*X*_*t*_ + *β*_*t*_ + *ϵ*_*t*_. Here *A*_*t*_ is a time dependent 4 *×* 4 matrix and *β*_*t*_ is a time dependent 4-dimensional vector.

Building a meaningful model of this form is impossible without any regularization assumptions on *A*_*t*_ and *β*_*t*_ as we have only one observation of *X* per each time instance *t*. We postulate a very natural restriction on possible values of *A*_*t*_, *β*_*t*_, which state that *F*_*t*_(*x*) and *F*_*t*+1_(*x*) should not differ drastically on the same input *x*. This represents a very simple principle that the mechanism *A*_*t*_, *β*_*t*_ driving the dynamics of the pandemic should not be fluctuate widely on the day to day and week to week basis.

Thus we fix two parameters *τ*_*A*_ and *τ*_*β*_, and postulate that for each *t* we have a bound

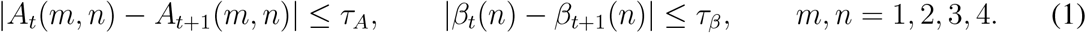

The parameters *τ*_*A*_, *τ*_*β*_ will be estimated from the data as described below. First for any fixed values *τ*_*A*_, *τ*_*β*_ and for our historical data series *X*_*t*_, *t ∈* [0, *T*], we obtain *A*_*t*_, *β*_*t*_ by solving the squared loss constrained minimization problem

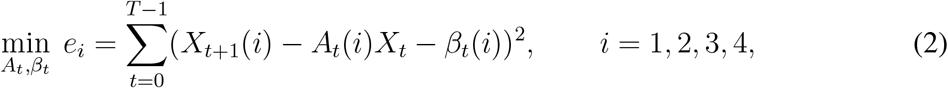

subject to the constraints (1). Here *X*_*t*+1_(*i*) and *β*_*t*_(*i*) denotes the *i*-th element of *X*_*t*+1_ and *β*_*t*_, respectively. In addition, *A*_*t*_(*i*) denotes the *i*-th row of *A*_*t*_. Note that *e*_1_, *e*_2_, *e*_3_, *e*_4_ denote the errors of incidences, mortality, hospitalization and mobility processes respectively. In this optimization problem *A*_*t*_, *β*_*t*_ are viewed as variable. Solving this optimization problem is straight-forward due to convexity of the objective function. Further, to obtain the best adjustment of the parameters *τ*_*A*_, *τ*_*β*_, we conduct a search over the choices of those which provide the smallest value of the objective function, thus providing the best fit. More precisely, if *O*(*τ*_*A*_, *τ*_*β*_) denotes the value of the objective function (2) as a function of *τ*_*A*_ and *τ*_*β*_, we conduct a search of the values of *τ* and *β* which give the smallest value of *O*(*τ, β*). This is done in straightforward by going the all choices of these parameters with some small discretization steps. The optimal values of *τ* ^*\**^ and *β*^*\**^ are reported in the next section. This approach is adopted for both for daily and weekly data, and also across various states US, along with US data as a whole. The associated optimal values of sequences *A*_*t*_, *β*_*t*_, *t ∈* [0, *T*] are denoted by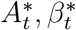. Along with 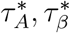 they represent the best fit dynamical system representation of the pandemics evolution. For any two times instances *t*_0_, *t*_1_ *∈* [0, *T*] and any data configuration 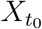 at time *t*_0_ we can compute the implied prediction 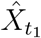 of the actual observable 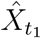 at time *t*_1_ by iterations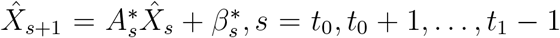. We note that each of these is a function of the observable 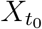 at time *t*_0_ and write 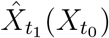 in order to emphasize this.

Our next step is similar to the one before: we investigate to what extend the dynamics is sensitive to the observations 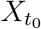. If the data analyst is currently positioned in time *t*_0_, she uses the observable 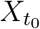 to make predictions 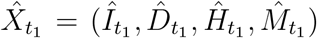 for the observables at time *t*_1_. If the sensitivity (to be defined now) is very small, it means that most of the information embedded in the data 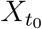 is “lost” and most of the future realization 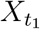 of the observable is due to the accumulation of the idiosyncratic noise values *ϵ*_*s*_, *s ∈* [*t*_0_, *t*_1_]. The sensitivity for the observable corresponding to the number of incidences is defined as max_*X,Y*_ 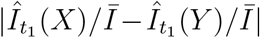 where *X* and *Y* vary over all possible choices of four dimensional vectors with coordinates in [*I*_min_, *I*_max_], [*D*_min_, *D*_max_], [*H*_min_, *H*_max_], [*M*_min_, *M*_max_] respectively. Here we recall that min and max values correspond to the smallest and largest values of the four observables observed over the entire period [0, *T*]. We denote this maximum value by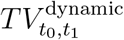. This is similar to the total variation and related measures of distances discussed earlier in the context of homogeneous Markov chains. We report its values in the following section. In particular, we find indeed that the value is very small when compared to the average *Ī*, once the forecasting horizon (*t*_0_, *t*_1_) is more than a couple of weeks for the daily observations and more than six to several months for weekly observations. This indicates that indeed the loss of information imbedded in the data 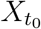 used for prediction is very significant to the point that it hardly provides any forecasting value.

We stress one very important point here. The values 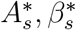 for *s ≥ t*_0_ are not in fact available to the data analyst at time *t*_0_, as those depend on the future realizations *X*_*s*_, *s ≥ t*_0_ of the observables. So in fact, while the series *A*_*s*_, *β*_*s*_ represents the best prediction of the future as a function of the current observable 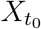, this cannot be implemented in real life. What we show, however, is that such predictions would be rendered useless beyond certain time horizons *even* if the values 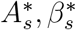 were given to the predictor in the hindsight, that is even for the predictor who is equipped with the clairvoyante power of having the access to the future values of *A*_*s*_ and *β*_*s*_.

## Results

### Homogeneous Markov chain based estimations

We now turn to the results of our computations based on the data and models described in the section Methods and models. Recall that our first metric of interest is the spectral radius *λ*_2_(*P*) of the associated Markov chain. We have computed it for the Markov chain process *I*_*t*_ corresponding the dynamics of the incidences, daily and weekly, at different discretization levels *n*. The spectral radius values associated with the process of incidences is reported on Figure 5 with discretization value *n* ranging from 2 to 10 for daily observations and the discretization value *n* ranging from 2 to 5 for weekly observations. Finer discretization becomes problematic due to insufficiency of data per chain transition observations (though finer discretization were achievable for mortality estimations). The largest, namely the most conservative value of the gap is *λ*_2_(*P*) = 0.868 when *n* = 10. While it is possible that the value can increase as *n* increases, we anticipate based on the plot that it will increase only marginally. With value *λ*_2_ = 0.868 as our estimation of the correct spectral radius, the rate of information decay (mixing) is then 0.868^*t*^ per *t* transitions of the chain. This is of course just a crude quantitative assessment of the rate of the information loss. In order to assess the actual loss of information, we report the metrics *TV*_2_(*t*), which we recall is the total sensitivity metric, and *TV* (*t, ρMax*(*I*)), namely the tail sensitivity metric. The values of *TV*_2_(*t*) for daily incident processes are report on Figure 6 for discretization levels *n* = 4, …, 10. We see that the value drops below 0.01 when *t* is in the range from 30 to 50 days. This means that the predictive power of the information of the number of incidences observed on day *X* loses 99% of its value for predicting beyond *X* + 30 to *X* + 50 days ahead. The results on a weekly basis are similar and reported on Figure 7: 99% information is lost within 30 to 40 weeks. While 40 weeks appears excessively long prediction period and has not been attempted, to the best of our knowledge, in the same plot we see that 90% of information is lost already within 15 to 25 weeks which is very much within ranges for which forecasting were produced in the past.

**Figure 5.**
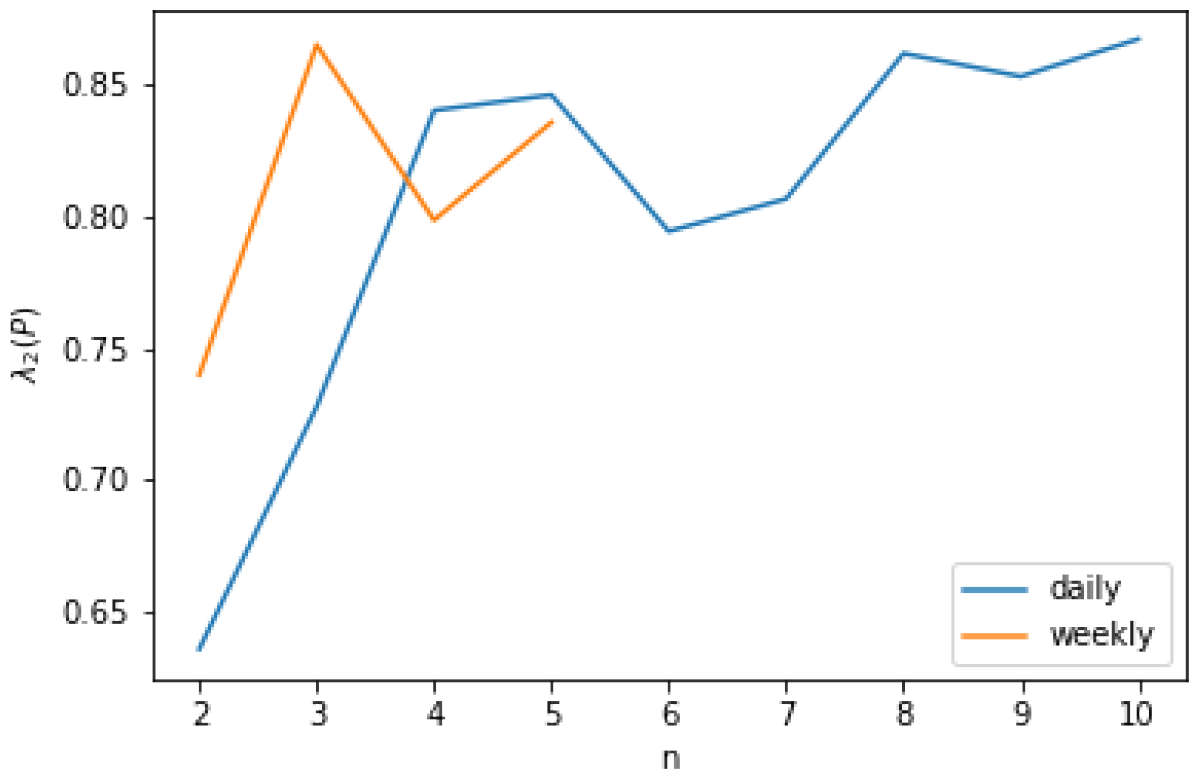
Spectral radius for incidences.

**Figure 6.**
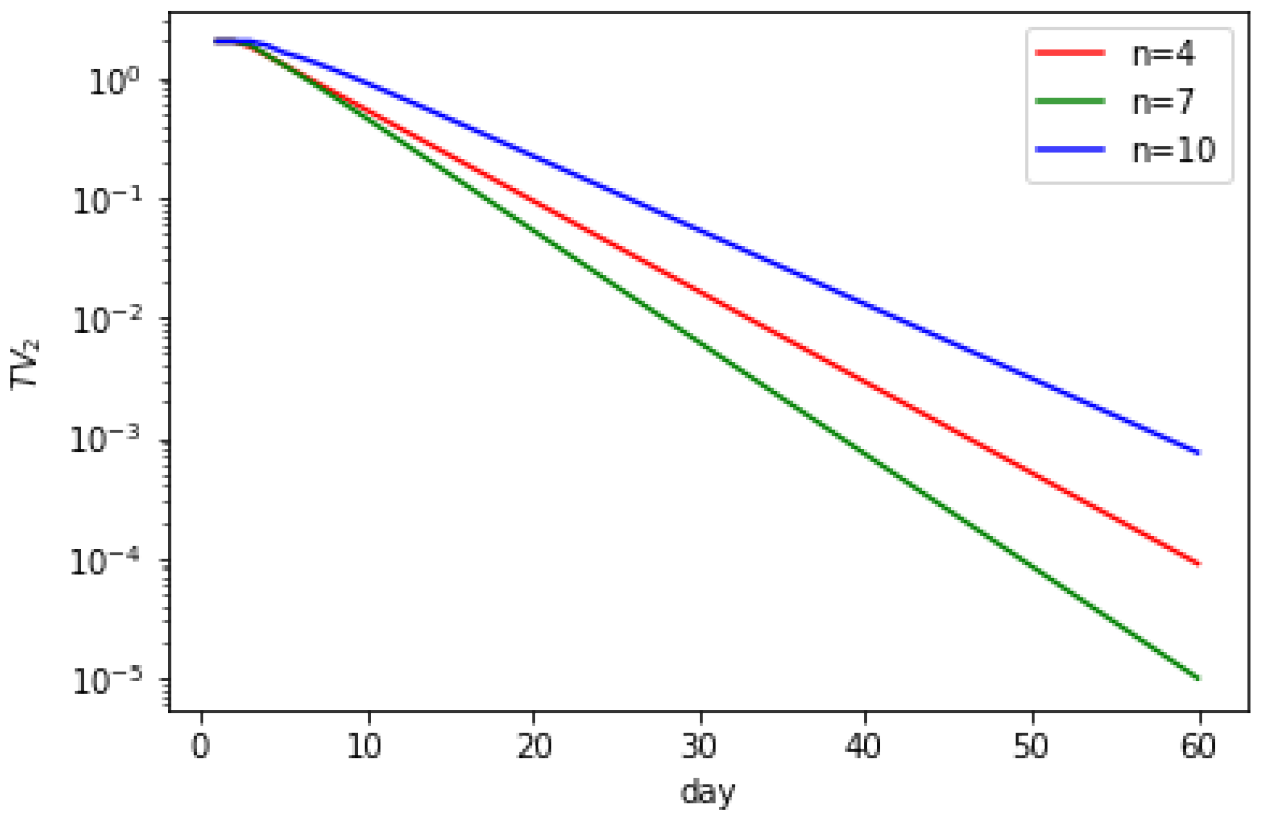
*TV*_2_(*t*) for daily incidences.

**Figure 7.**
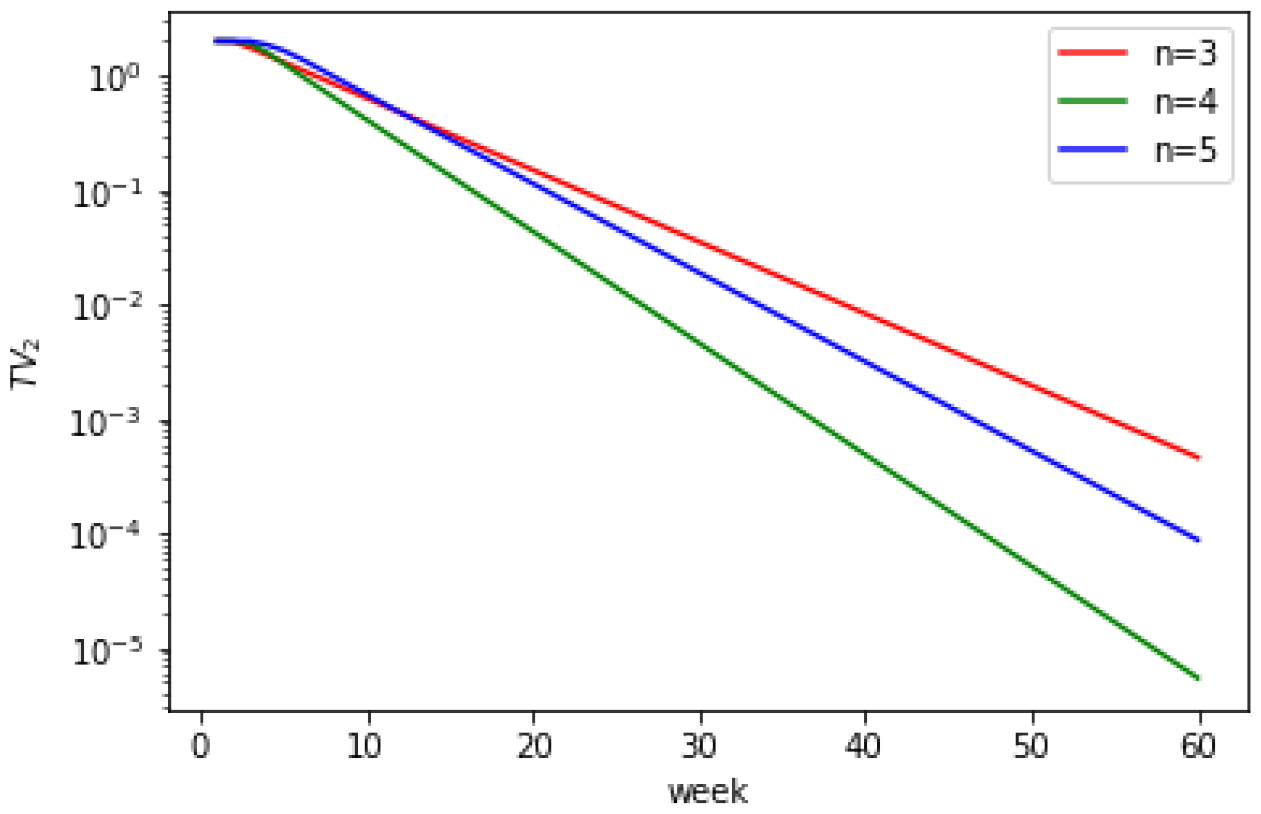
*TV*_2_(*t*) for weekly incidences.

The results are even more compelling when we look at the metric *TV* (*t, ρMax*(*I*)) which is reported for value *ρ* = 0.7 on Figure 8 and *ρ* = 0.8 on Figure 9 for daily incidences, and on Figures 10 and 11 for weekly incidences for the same choices of *ρ*. Recall that *TV* (*t, ρMax*(*I*)) measures the power of the observable *I* on day *X* to predict the likelihood of seeing *ρ*-percent historically high incidences on day *X* + *t*. For daily incidences the value of this metric drops below 0.01 within 15 to 25 days based on *ρ* = 0.7, and within 10 to 20 days based on *ρ* = 0.8. Namely, the information embedded in *I* on day *X* amounts to at most 1% of the outcome of *I* on day *X* + *t*. Recall that, as we have explained earlier, this means that *any* amount of change in *I* on day *X* will result in change of *I* on day *X* + *t* which is at most 0.01 times the probability of an outcome. For the weekly data the results are similar: the value drops below 0.01 within 20 to 25 weeks for *ρ* = 0.7 and *ρ* = 0.8.

**Figure 8.**
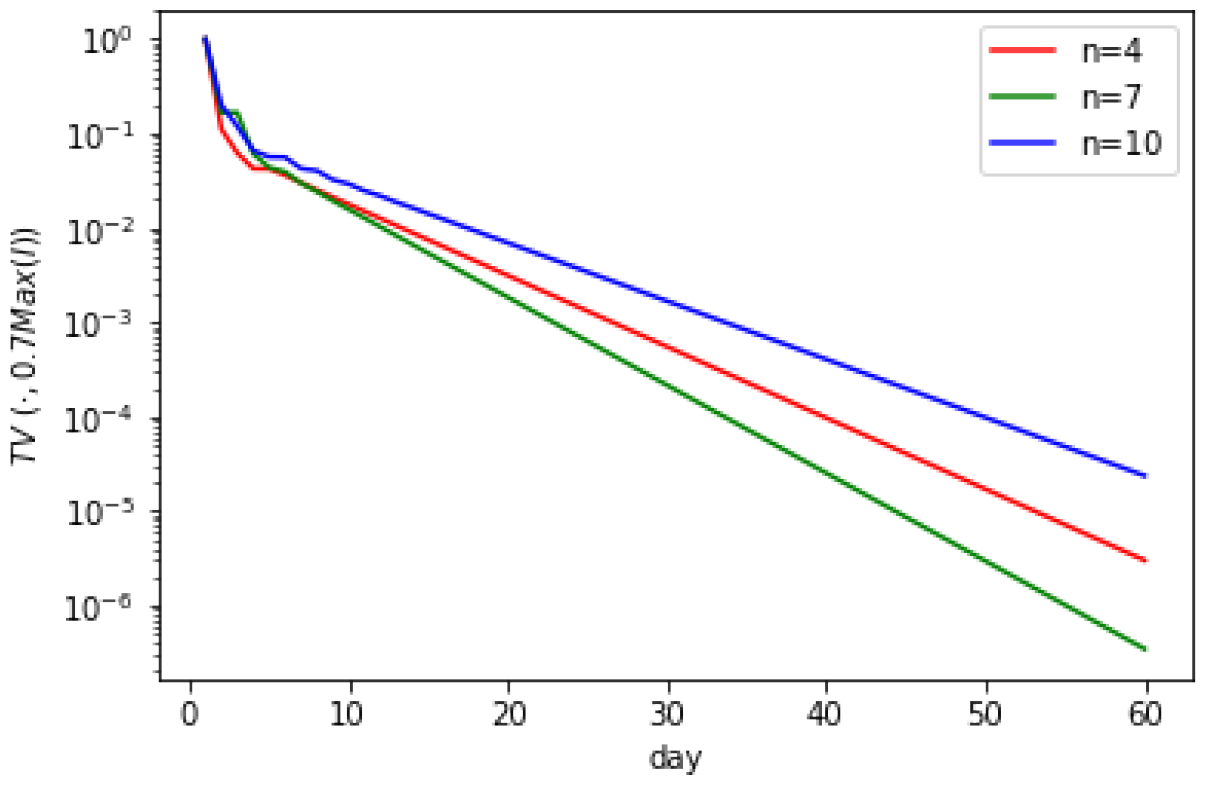
*TV* (*t*, 0.7*Max*(*I*)) for daily incidences.

**Figure 9.**
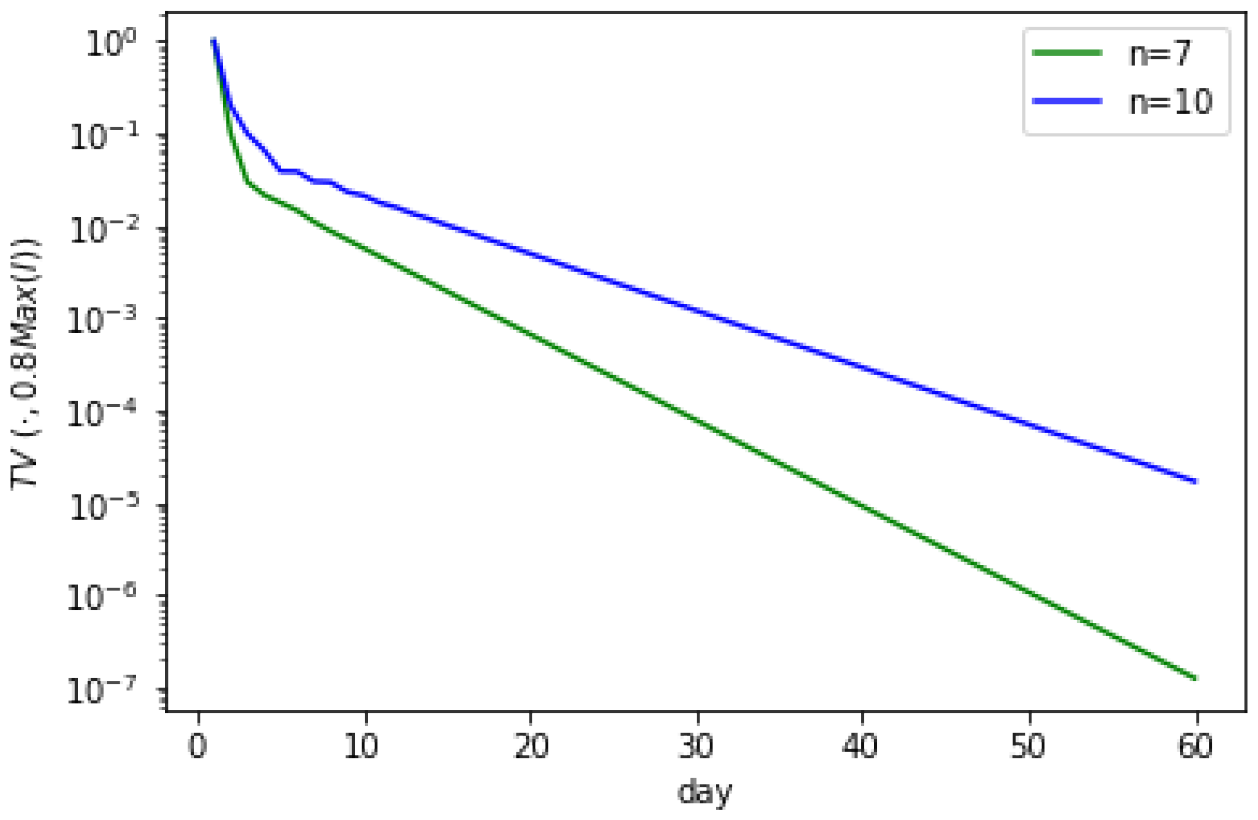
*TV* (*t*, 0.8*Max*(*I*)) for daily incidences.

**Figure 10.**
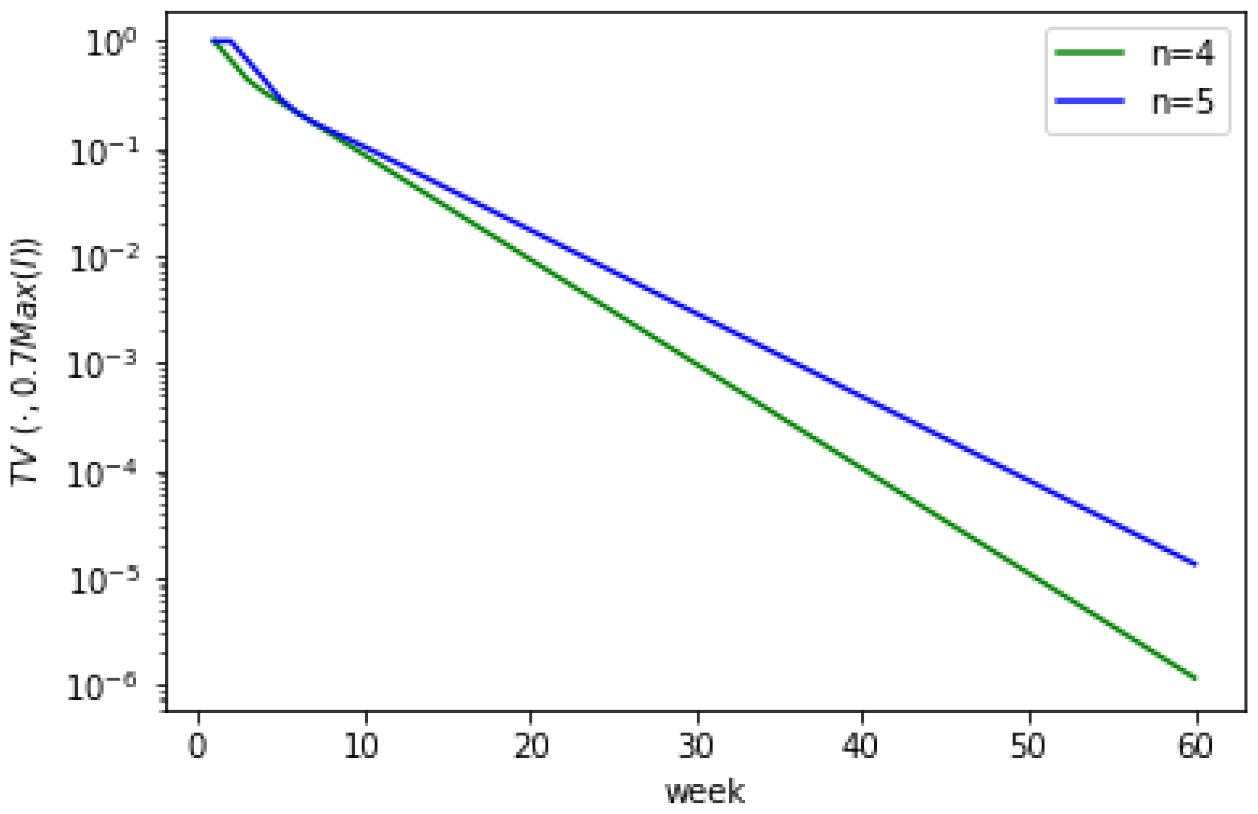
*TV* (*t*, 0.7*Max*(*I*)) for weekly incidences.

**Figure 11.**
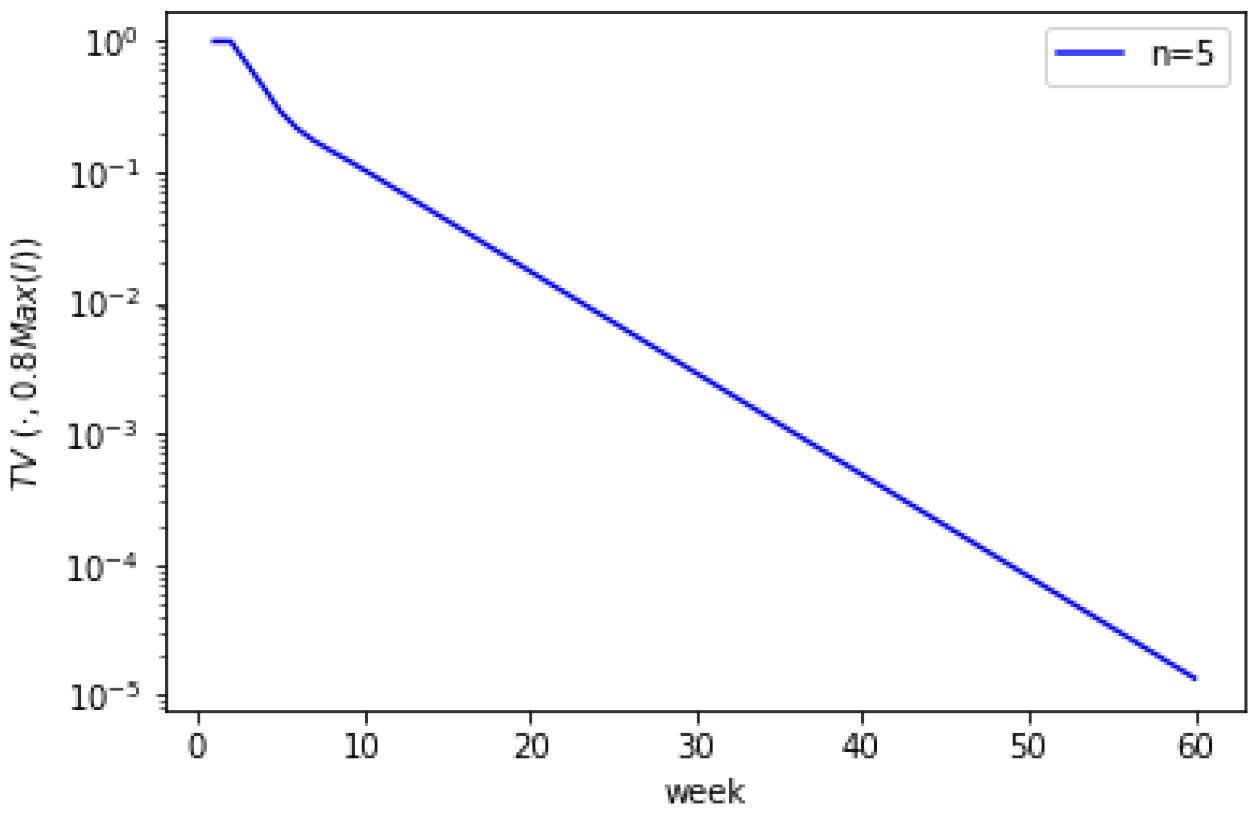
*TV* (*t*, 0.8*Max*(*I*)) for weekly incidences.

We conducted similar computations with incidences (*I*) replaced by mortality (*D*). We found that spectral gaps can be estimated based on somewhat finer discretization and in fact the estimations appear to settle when when the discretization is about *n* = 10 both for daily and weekly mortality, see Figure 12. We found that metrics *TV*_2_ and *TV* settle to values less than 0.01 within about 10 days, Figures 13-15. For weekly observables, however, the values are significantly higher and by roughly 30 weeks they only drop to 0.1, signifying a loss of 90% (as opposed to 99% for daily observations), see Figure 16-18. This is also consistent with estimations of the spectral radius, which settles to value approximately 0.75 for daily mortality observation, but a pretty high value of approximately 0.95 for weekly mortality observations. Whether this suggests that mortality observed on a weekly basis can be predicted more accurately than incidences observed on a weekly basis, is an interesting question for a future research.

**Figure 12.**
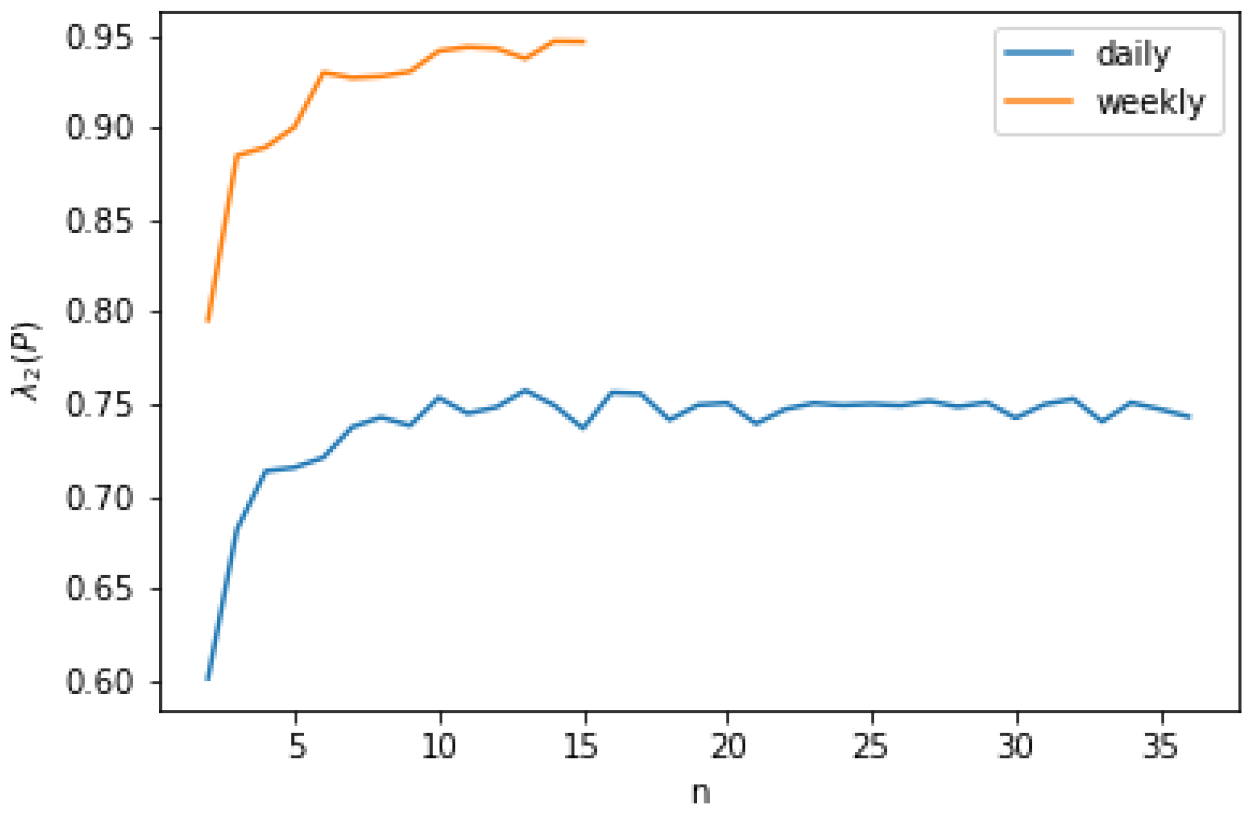
Spectral radius for mortality.

**Figure 13.**
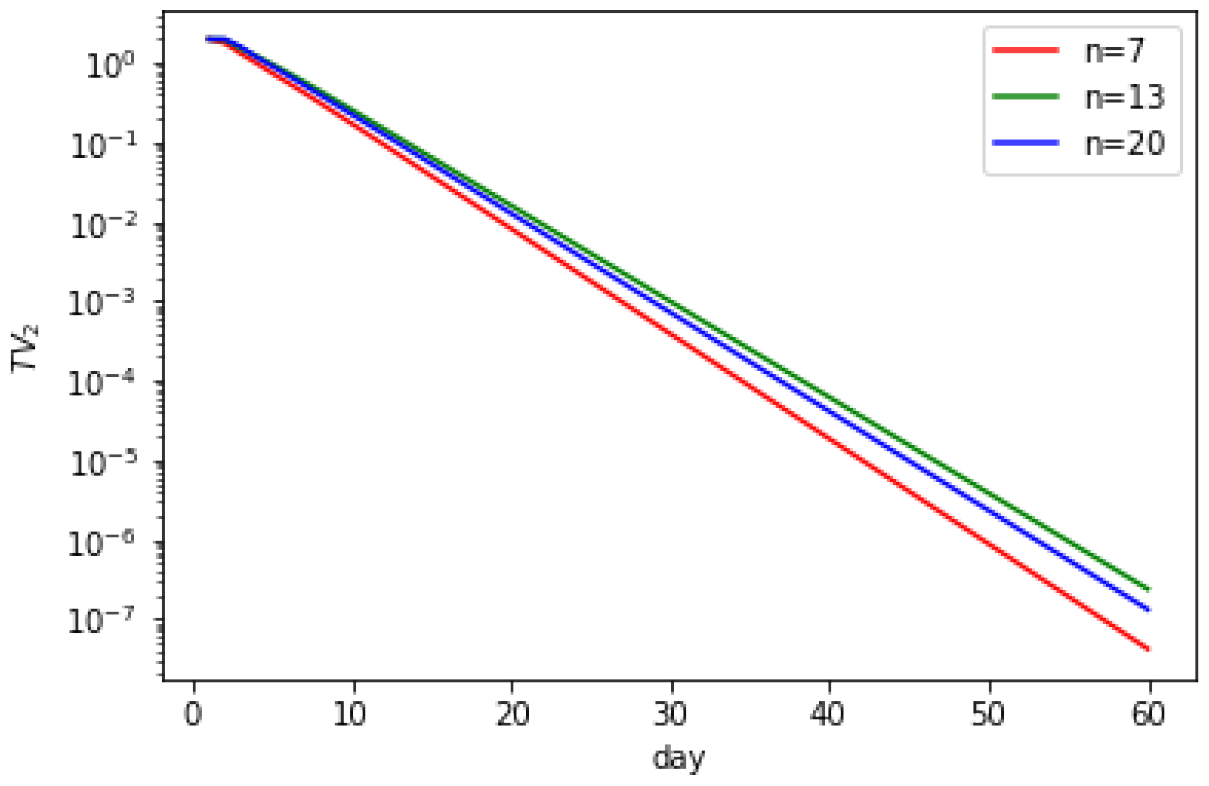
*TV*_2_(*t*) for daily mortality.

**Figure 14.**
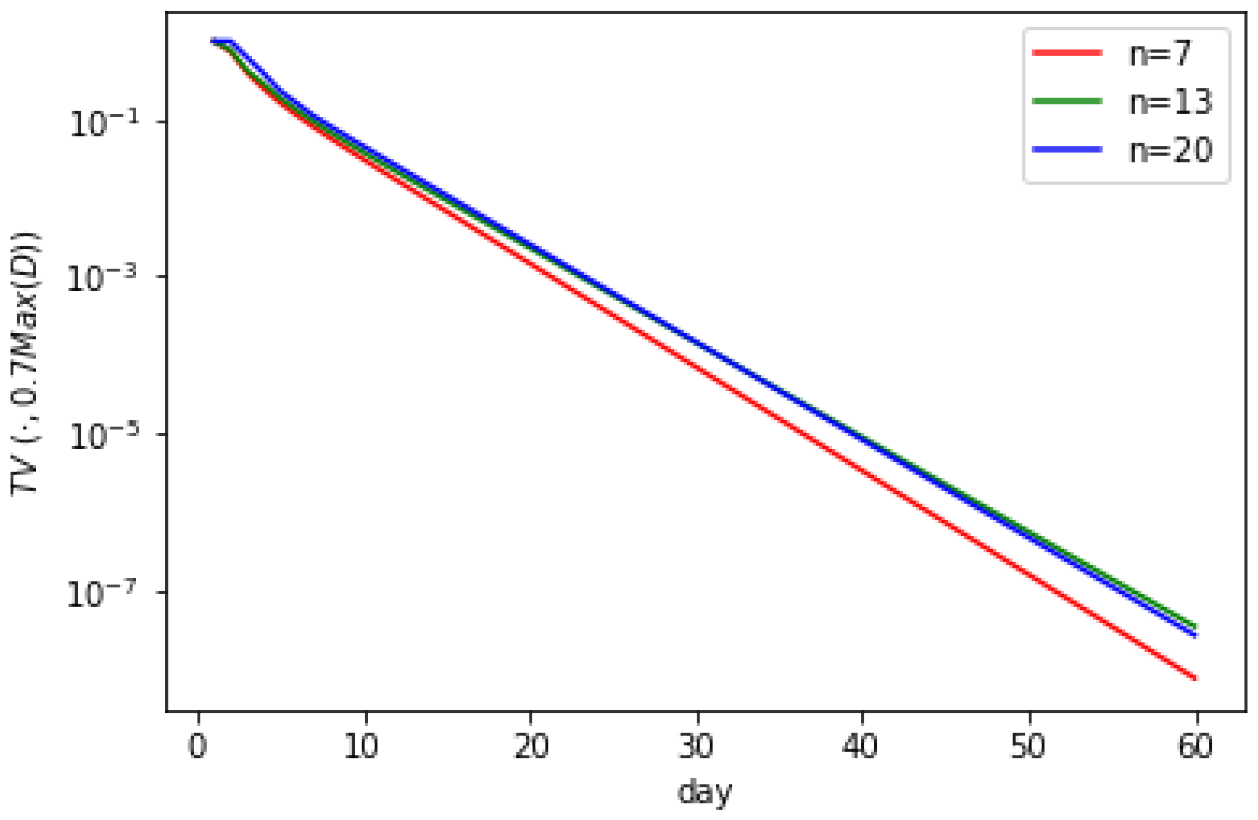
*TV* (*t*, 0.7*Max*(*D*)) for daily mortality.

**Figure 15.**
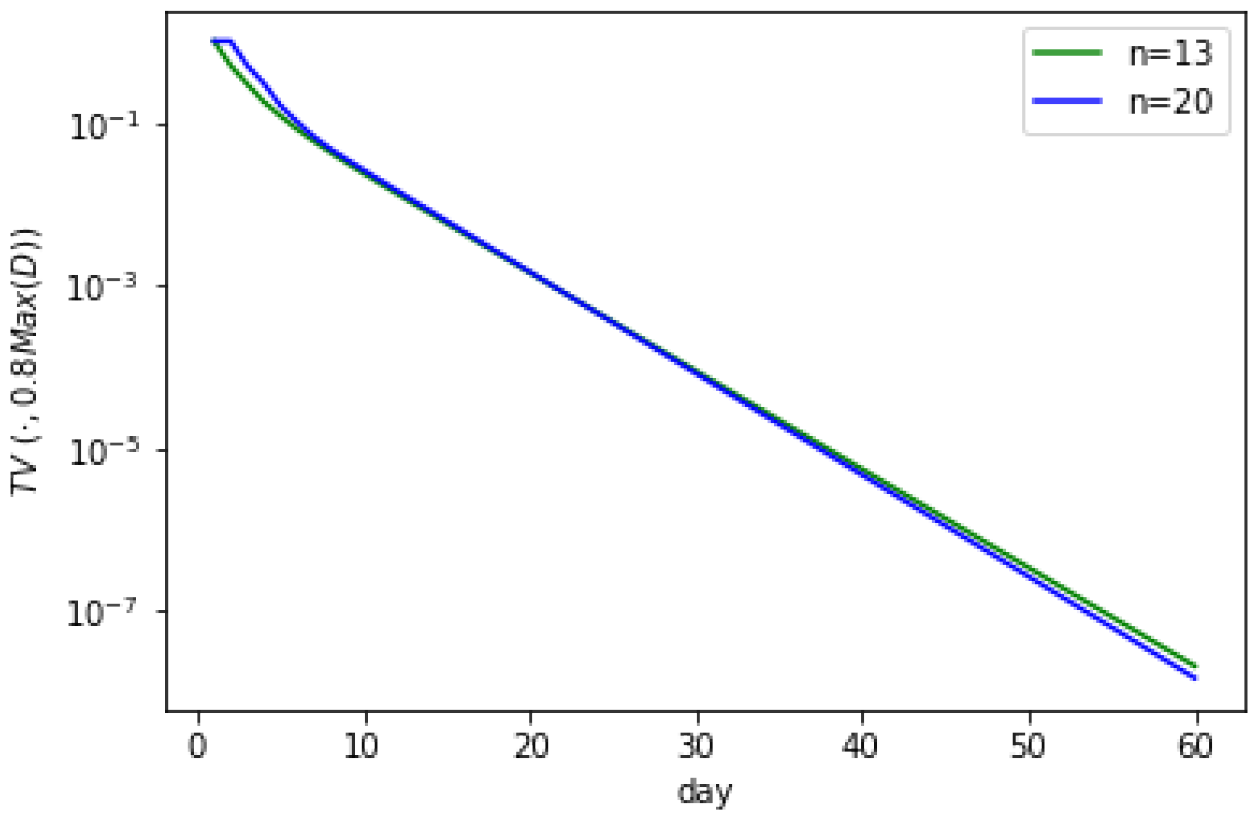
*TV* (*t*, 0.8*Max*(*D*)) for daily death.

**Figure 16.**
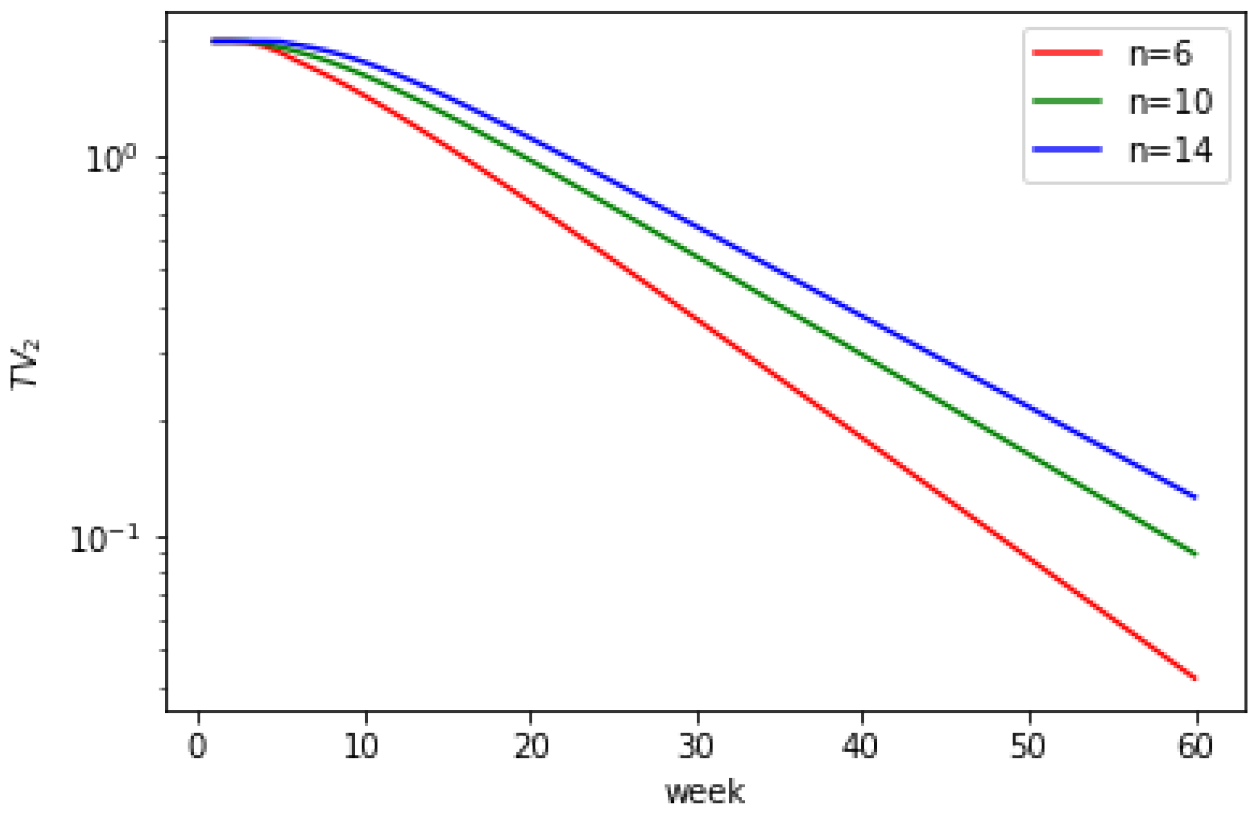
*TV*_2_(*t*) for weekly mortality.

**Figure 17.**
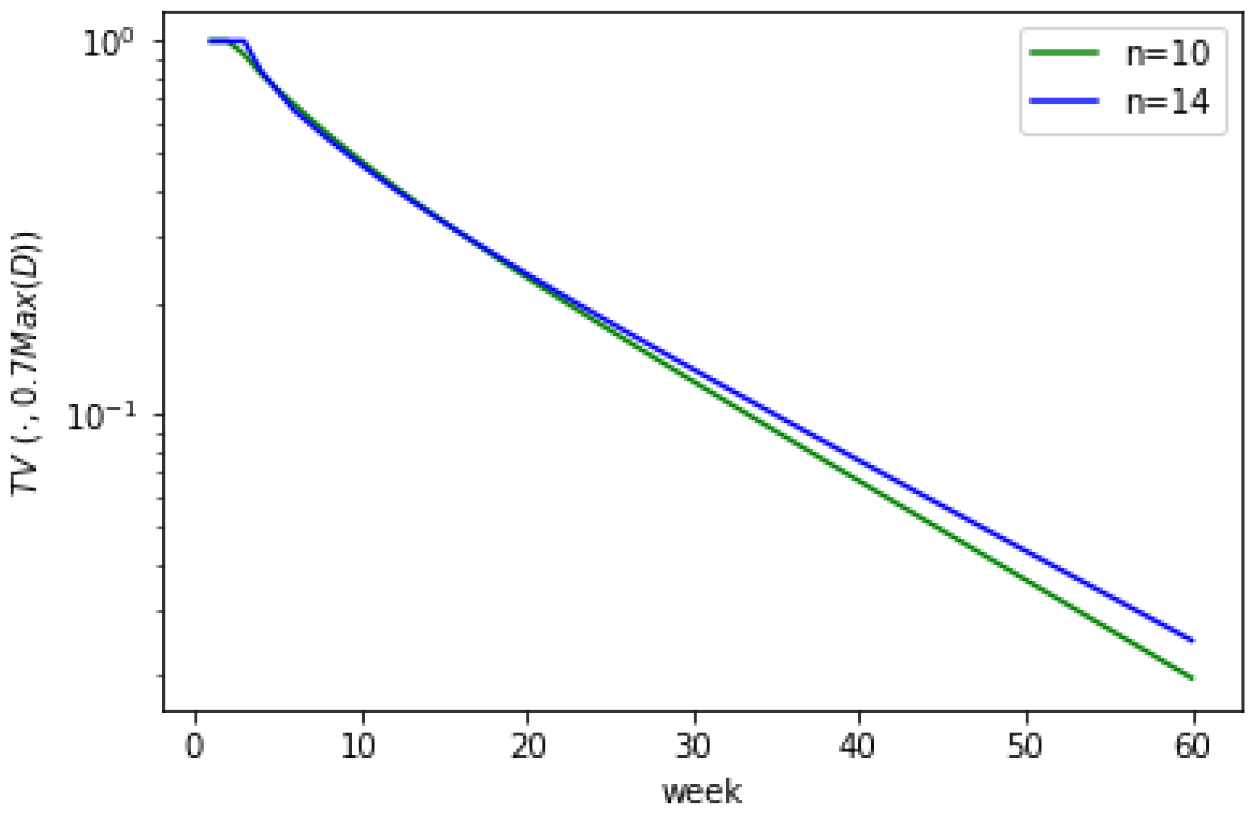
*TV* (*t*, 0.7*Max*(*D*)) for weekly mortality.

**Figure 18.**
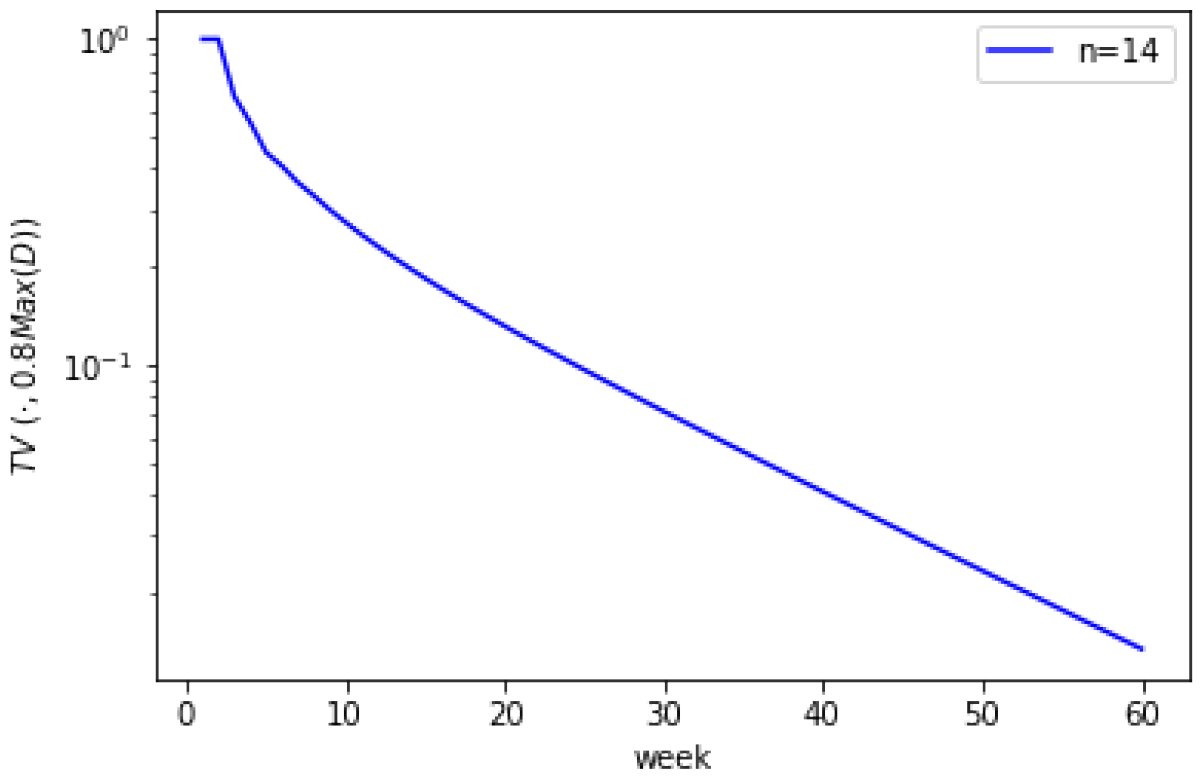
*TV* (*t*, 0.8*Max*(*D*)) for weekly death.

### Estimations based on heterogeneous regularized auto-regressive process

Next we turn to reporting our results based on the second Model II, namely inhomogeneous regularized auto-regressive processes.

First we describe the process of estimation of parameters *τ*_*A*_ and *τ*_*β*_. The simplest approach for estimating them would be to find values of these parameters which make the value of the optimization problem (2) the smallest. But this would correspond to the in-training data subject to the overfitting issue. Rather, instead we split the entire data series (per day and per weekly) into two equal parts, the first corresponding to the even days (weeks) and the second corresponding to the odd days (weeks). The first (even) data set is used for training, and the second (odd) data set is used for testing. Note that if *τ*_*A*_, *τ*_*β*_ are too large, the training data can be perfectly fitted with *A*_*t*_, *β*_*t*_, and the training error will be 0. Such cases correspond to overfitting. Thus, we set an upper bound on *τ*_*A*_, *τ*_*β*_, i.e. (*τ*_*A*_, *τ*_*β*_) *∈* [0, 10]^2^. Our experiment shows that when going beyond this upper bound, the resulting training error is always zero, which is very suboptimal. Moreover, we require the optimal *τ*_*A*_, *τ*_*β*_ to have training error at least 10^*−*10^. For each choice of *τ*_*A*_, *τ*_*β*_ within [0, 10], we solve the optimization problem (2) using the training data. This is done by grid search, that is a finite subset of points in [0, 10]^2^ which covers all points within a distance 0.01. For incidences process, we solve for 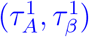 with the smallest testing error *e*_1_. For mortality and hospitalization process, we solve for 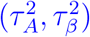 and 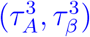 with the smallest testing error *e*_2_ and *e*_3_, respectively. We found the optimal values to be 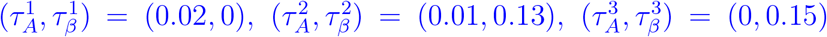 for daily observations and 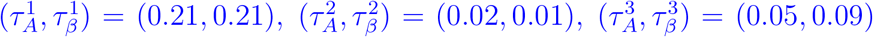 for weekly observations.

Next we compute the point forecast sensitivity metric 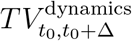 which we have described in the Subsection “Model II. Heterogeneous regularized auto-regressive process”. We do this for time lags of Δ = 1, 3, 5, 10, 20 and 30 days and report the values of this metric for all times *t*_0_ within the entire range *t*_0_ *∈* [0, 742]. Figures 19 and 20 report our results for the normalized Incidences metric *Ī*_*t*_ on daily and weekly basis respectively. The horizontal dashed lines correspond to medians of the respecting curves across the entire range. For daily observations we see the drop to below roughly 0.01 value with Δ = 10 days lookahead. As before, this means that less than 1% of information about the number of incidences 10 days ahead of predicting time is explained by the data used for the prediction, and the remaining 99% is due to the noise which accumulates within these 10 days. We stress again that this is the case even when the predictor is *equipped with the foresight power* of knowing the actual drivers *A*_*t*_, *β*_*t*_ of the dynamics for these future 10 days of the predicting horizon.

**Figure 19.**
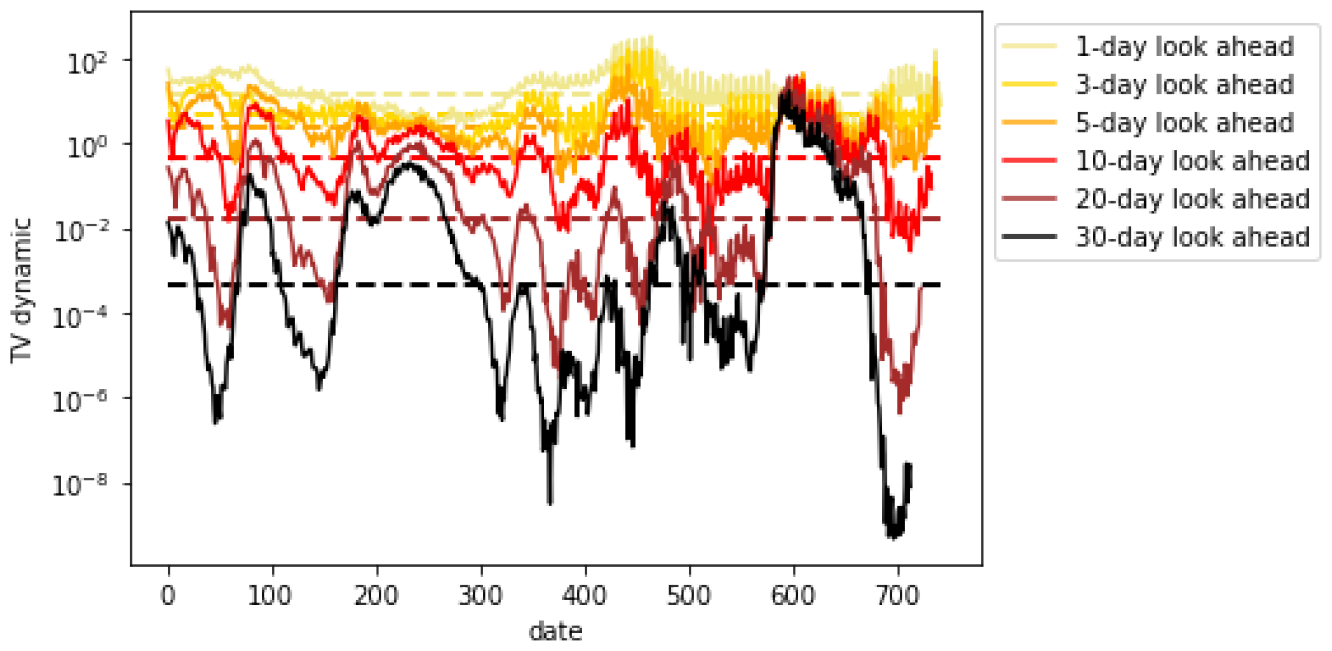
*TV* ^*dynamic*^ for daily cases.

**Figure 20.**
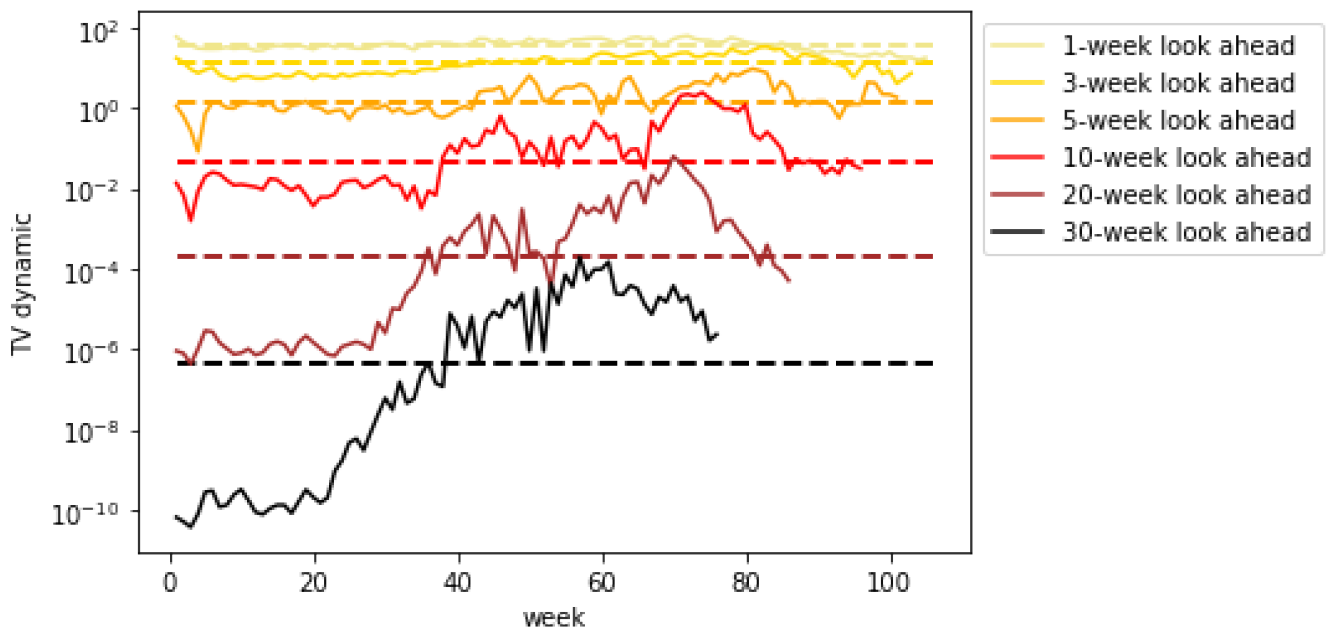
*TV* ^*dynamic*^ for weekly cases.

For the weekly observations, the loss of information is not as dramatic, but it is still significant. We find that the metric 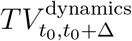 drops to the value below roughly 0.03 within Δ = 30 weeks period. Whether this means that predicting the number of weekly incidences for the horizon of *t <* 30 weeks ahead is feasible, is yet to be seen. But based on our result we conclude that predicting beyond 30 weeks does not appear possible.

Our estimation of information loss for predicting mortality is similar (reported on Figures 21 and 22): 99% information loss within 10 days for daily observations and about 97% information loss within 30 weeks. The results for hospitalizations are similar and reported on Figures 23-24. We have also conducted a similar analysis for several individual states. While, the results paint a more mixed picture, likely reflecting smaller amounts of aggregated data per states as opposed to national data, the general trend is similar: the information loss is clear beyond certain horizon level which depends on the level of aggregation (daily vs weekly).

**Figure 21.**
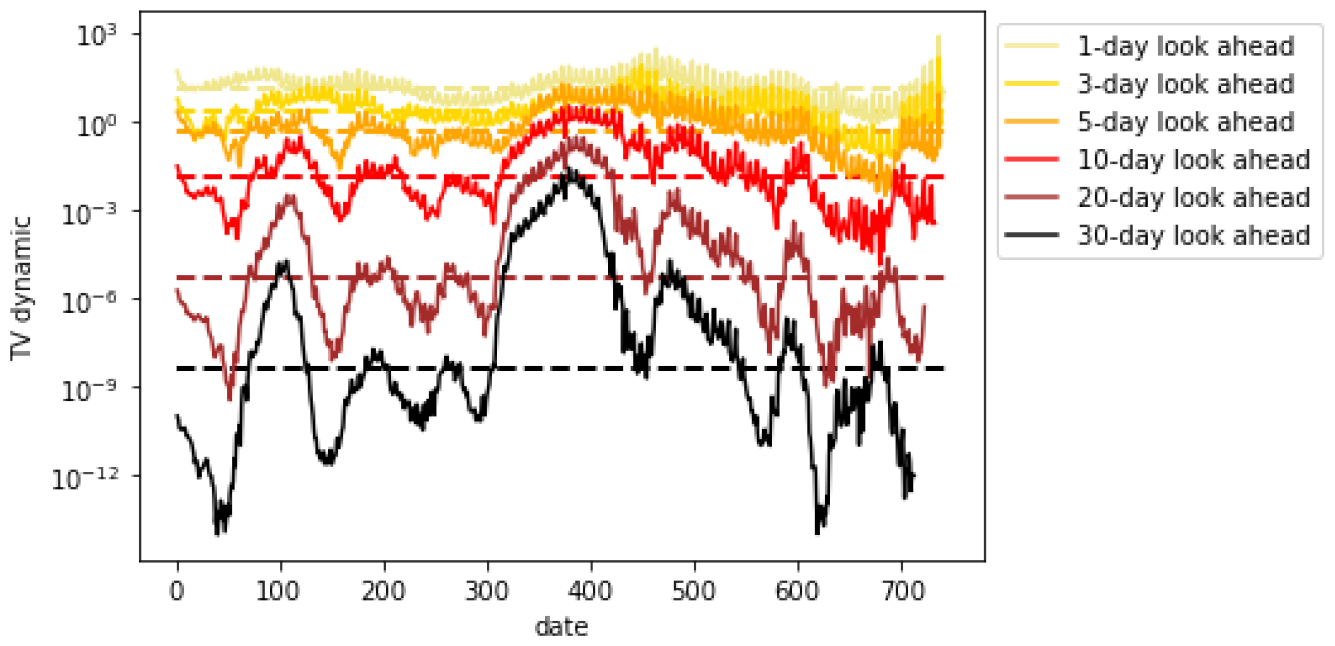
*TV* ^*dynamic*^ for daily mortality.

**Figure 22.**
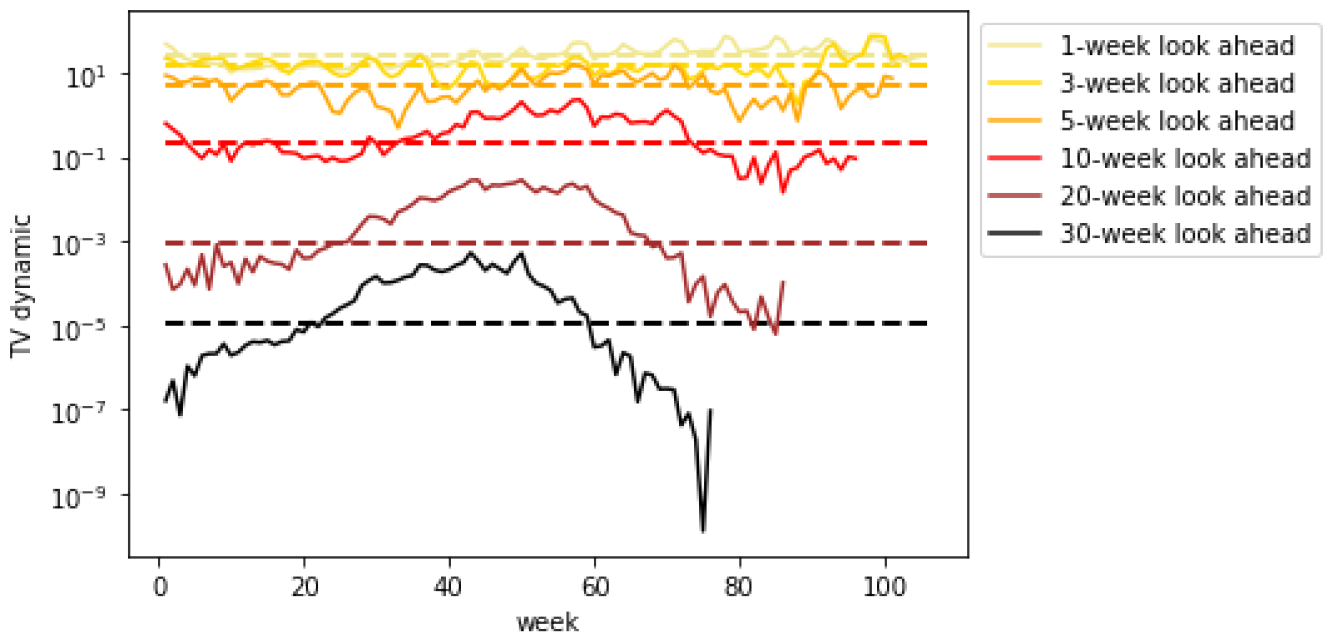
*TV* ^*dynamic*^ for weekly mortality.

**Figure 23.**
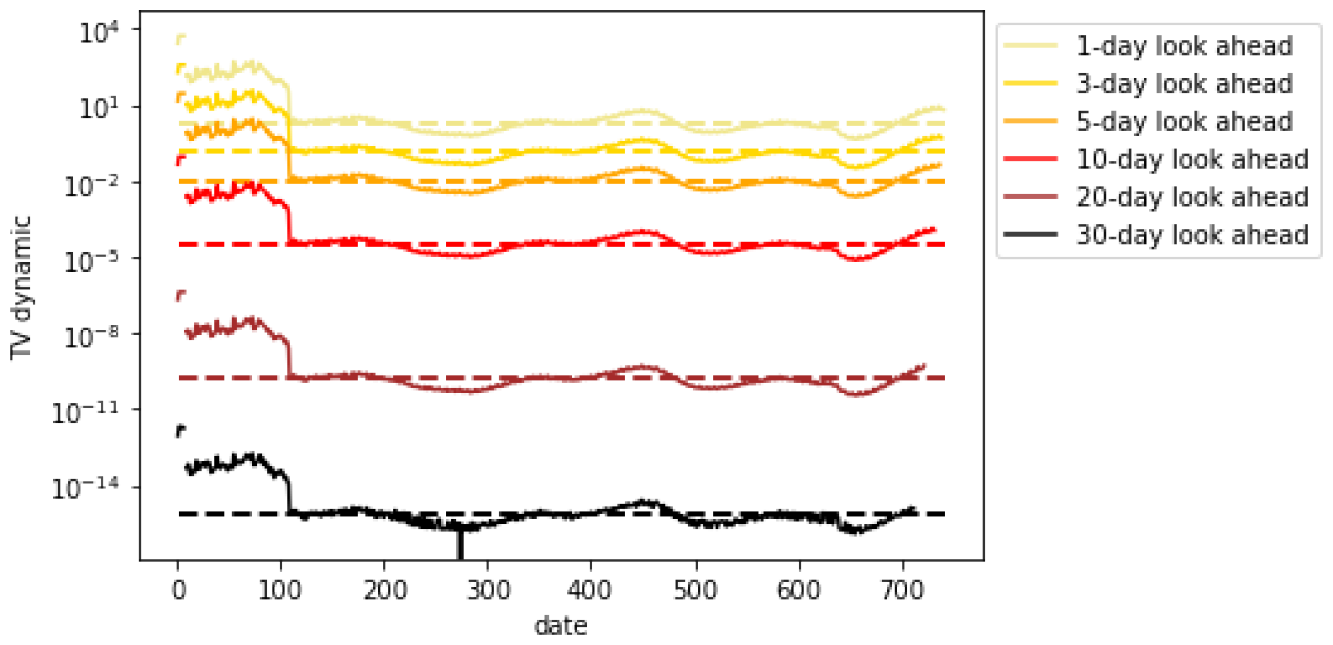
*TV* ^*dynamic*^ for daily hospitalizations.

**Figure 24.**
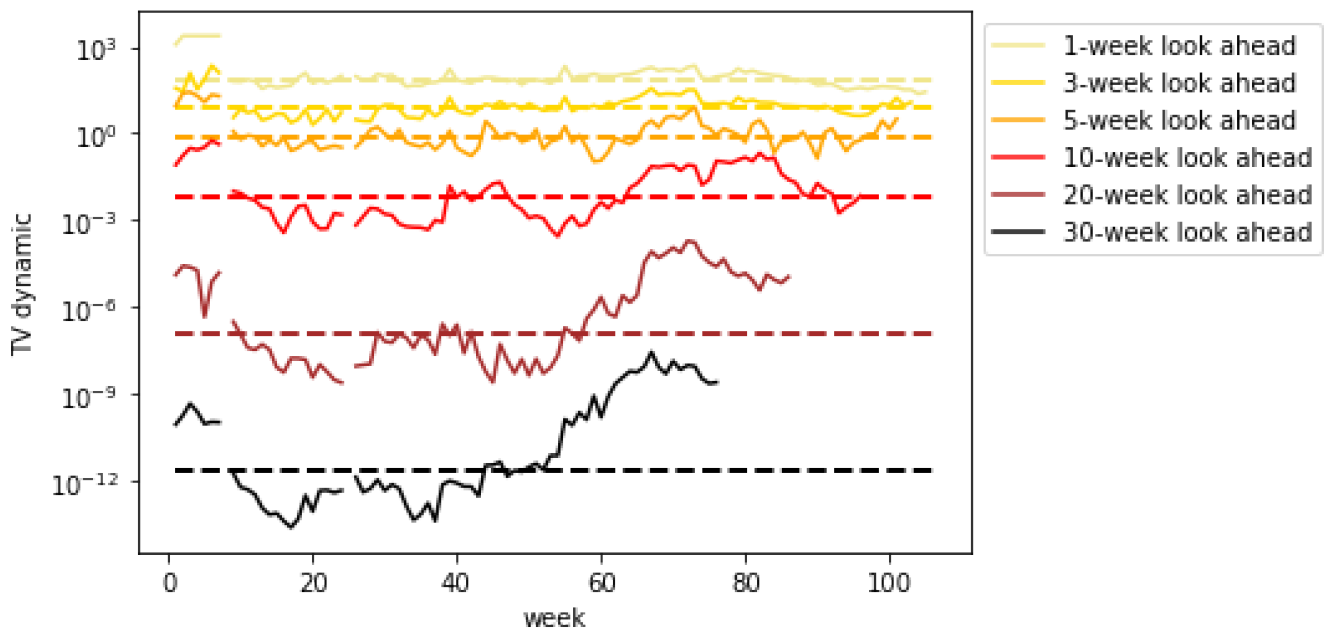
*TV* ^*dynamic*^ for weekly hospitalizations.

## Conclusions

The following question was investigated in the present paper: to what extend the future rates of COVID-19 incidences can be accurately forecasted? While the existing empirical studied showed a very limited success in forecasting beyond certain time horizons, and put forward forecasts which turned out to be orders of magnitude incorrect, to the best of our knowledge there are no studies devoted to understanding barriers for long term forecasting. Our paper is thus the first one of this kind. Our main conclusion is that predicting the number of incidences on daily basis given the information available at the time when the prediction is being made, is impossible beyond roughly 20 or so days horizon, and on weekly basis beyond 30 or so weeks horizon.

While the results we have reported applied exclusively to COVID-19 dynamics, we believe that the methods we have introduced in this paper are broad enough to understand limits of predictability in many other fields were predictions are routinely conducted, including areas of economics, finance and health care. The shortage of methods establishing limits of predictability is partially explained by the fact that these limits are usually tied with concrete predictive methodologies. Across statistical and machine learning applications pretty much every predictive methods comes with an understanding of its limitations in certain stylized setting usually involving independent identically distributed data. In our work we depart from this setting and conduct a study which is not tied to some particular predictive methodology, and instead applies to a broader class of methods. Specifically, we stress that the heterogeneous regularized autoregressive process we have adopted in our paper, which was our main framework to understand barriers for forecasting, *is not* a predictive methodology since it is based on affine process drivers *A*_*t*_, *b*_*t*_ which *are not* estimable at the time when the predictions are made. We believe that the general the theory of understanding limits of predictability should be based on considering widest possible model classes, in particular beyond those which are known to succeed in predicting for short term horizons, as this makes our limitation arguments even more compelling.

## Data Availability

All data produced in the present study are available upon reasonable request to the authors

## Acknowledgments

**Figure 25.**
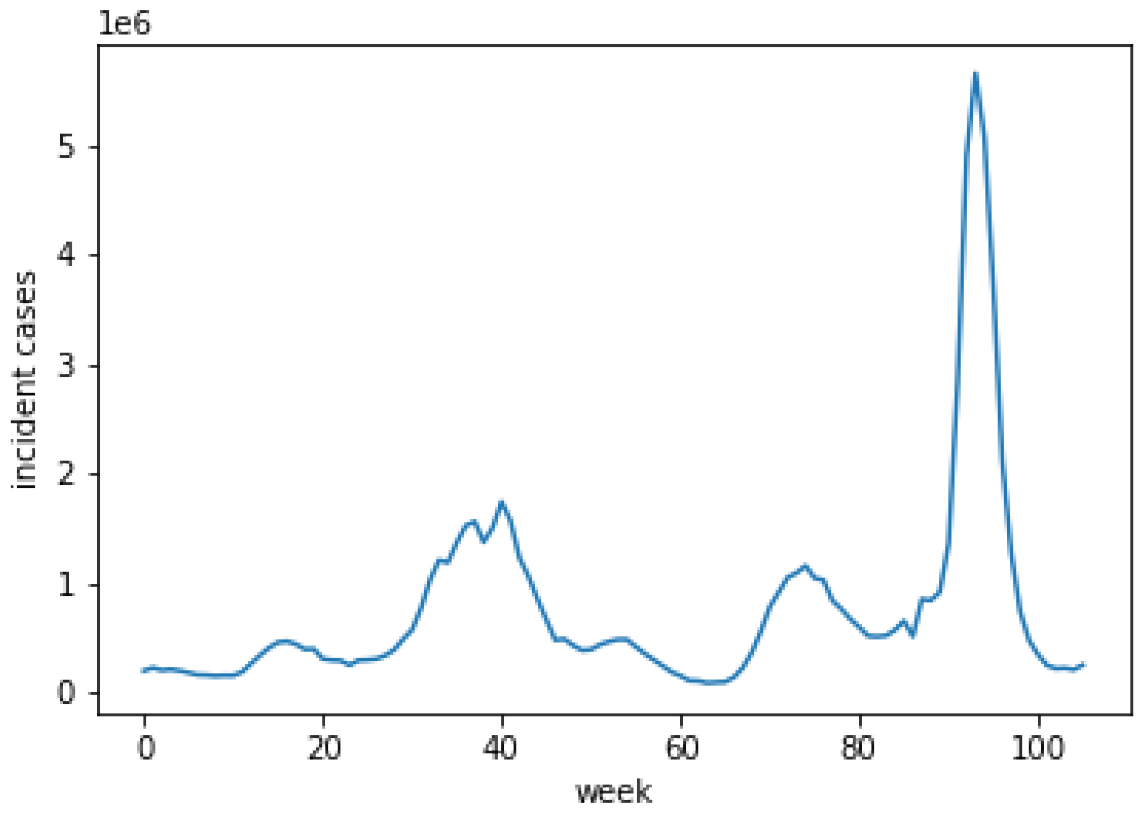
Time Series of Weekly Incident Cases.

## Supplementary materials

### Homogeneous Markov chain based estimations

**Figure 26.**
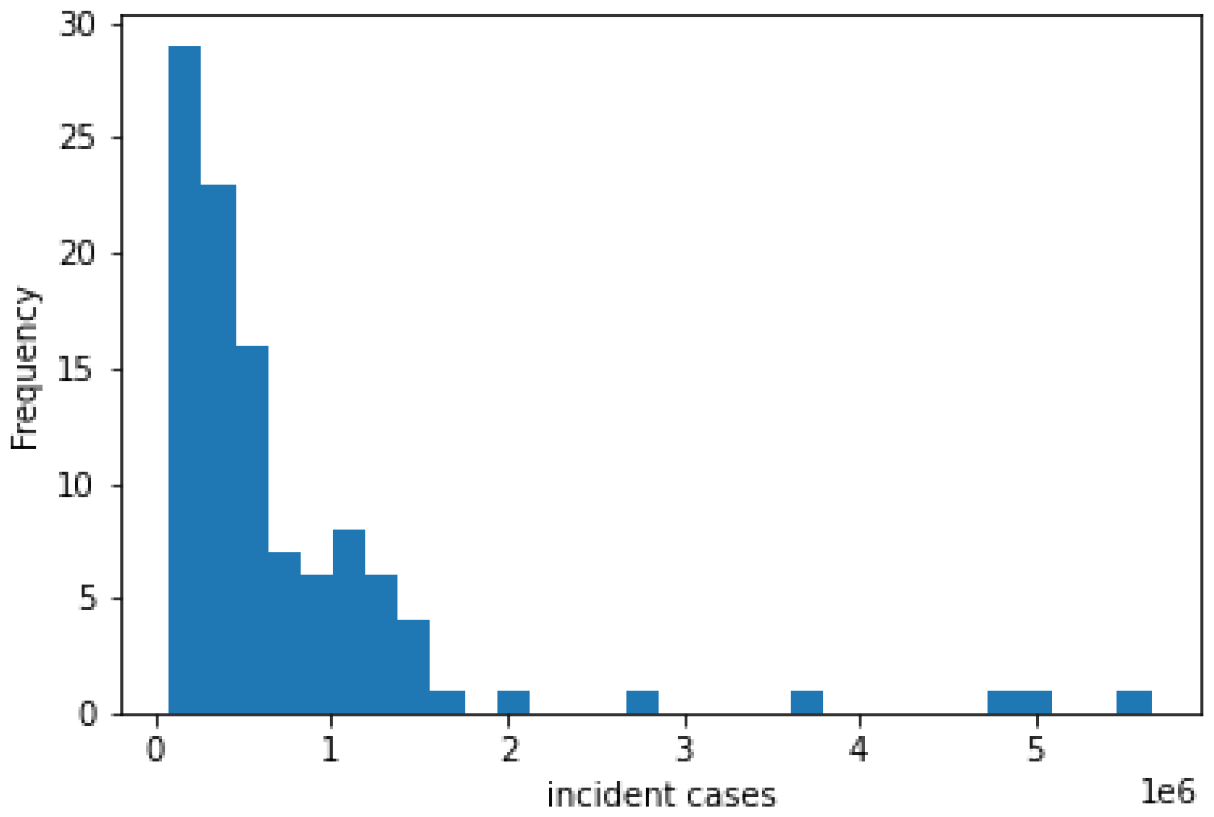
Distribution of Weekly Incident Cases.

**Figure 27.**
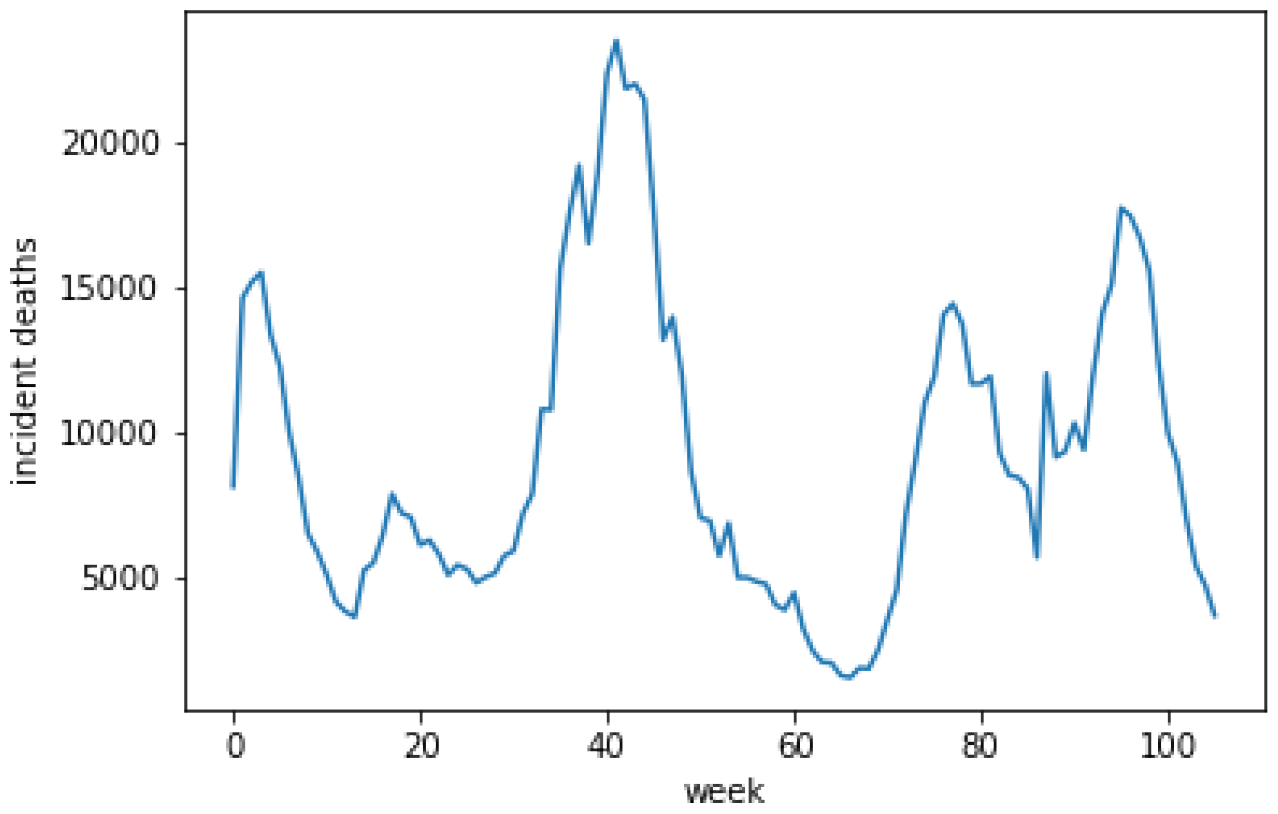
Time Series of Weekly Incident Deaths.

**Figure 28.**
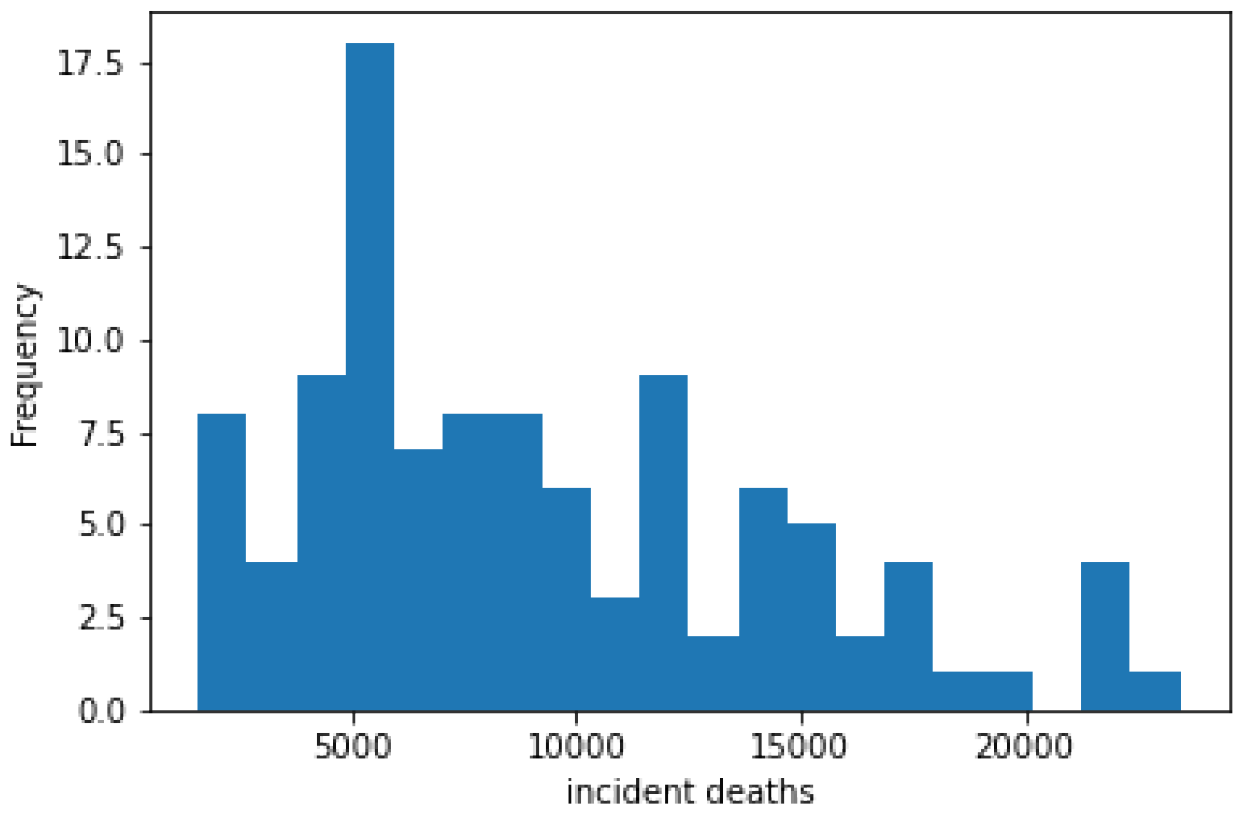
Distribution of Weekly Incident Deaths.

**Figure 29.**
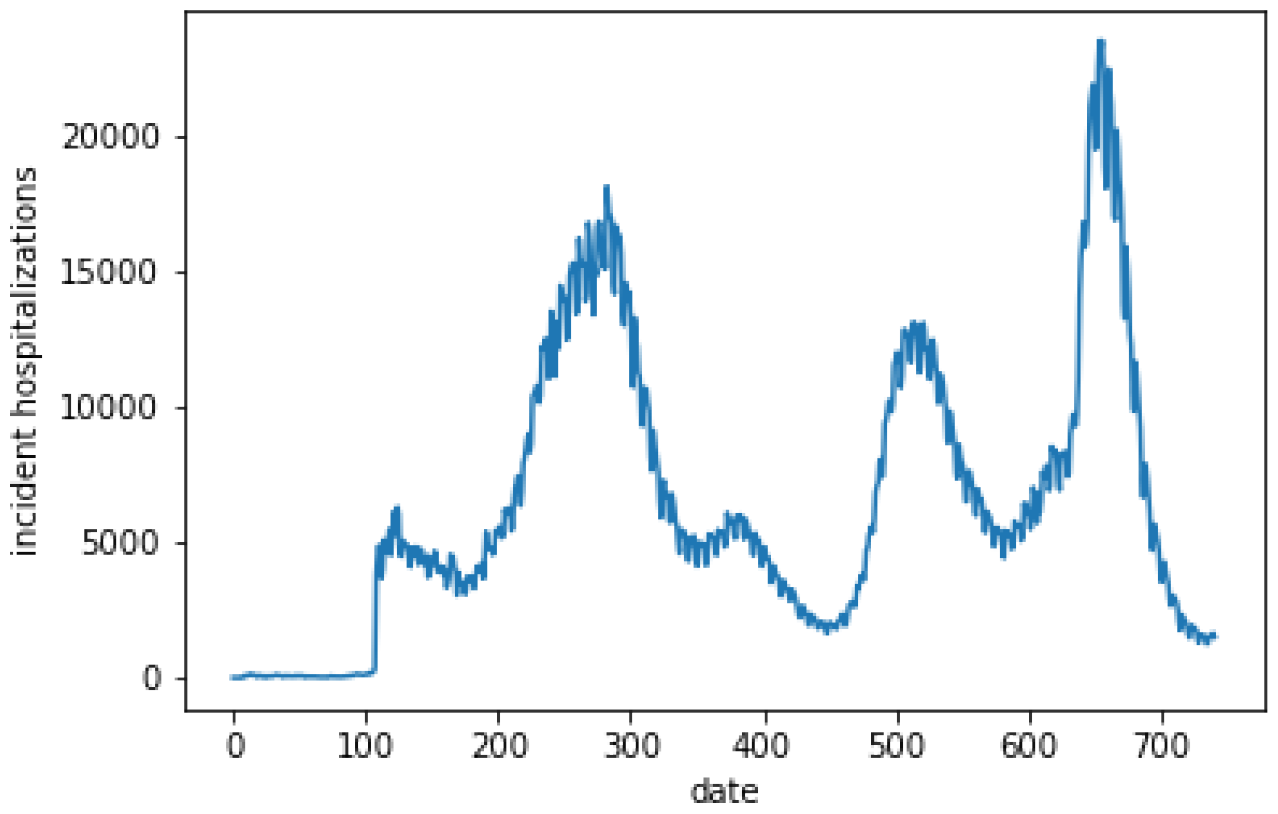
Time Series of Daily Incident Hospitalizations.

**Figure 30.**
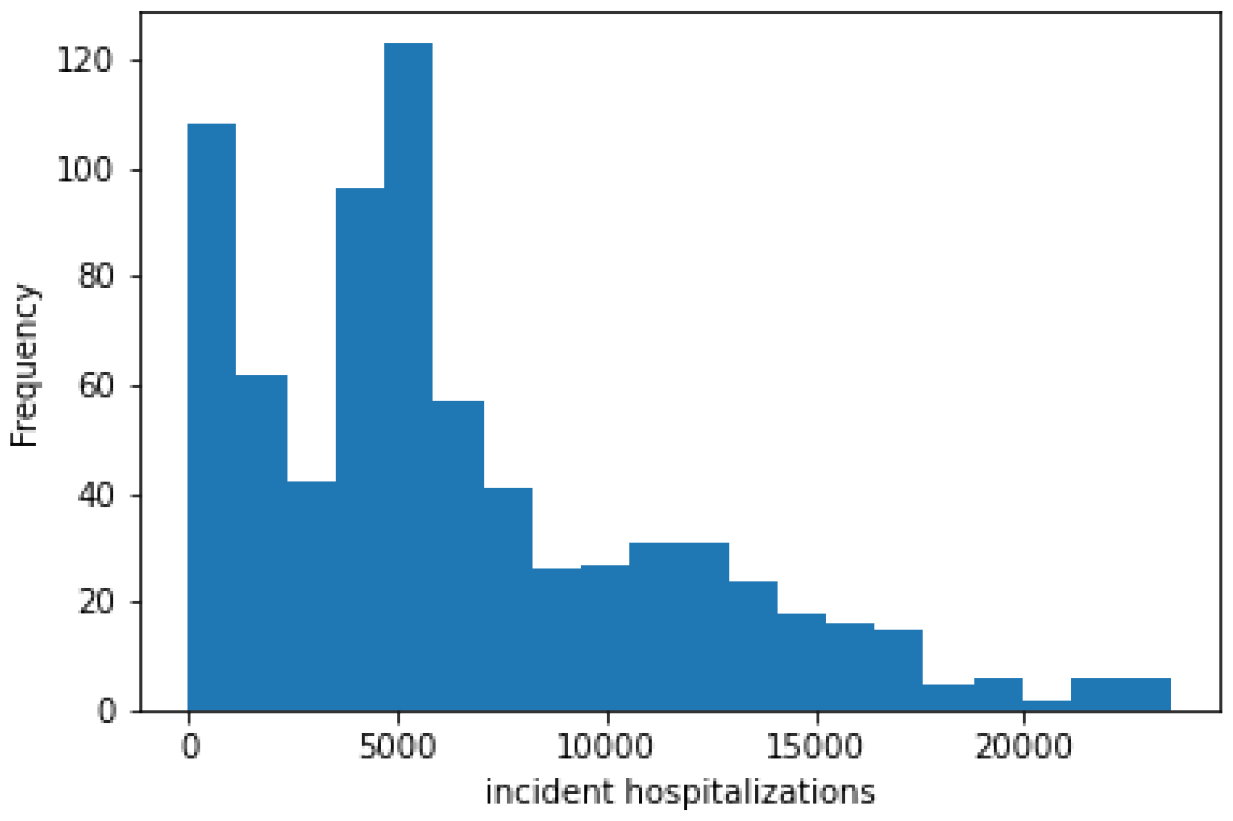
Distribution of Daily Incident Hospitalizations.

**Figure 31.**
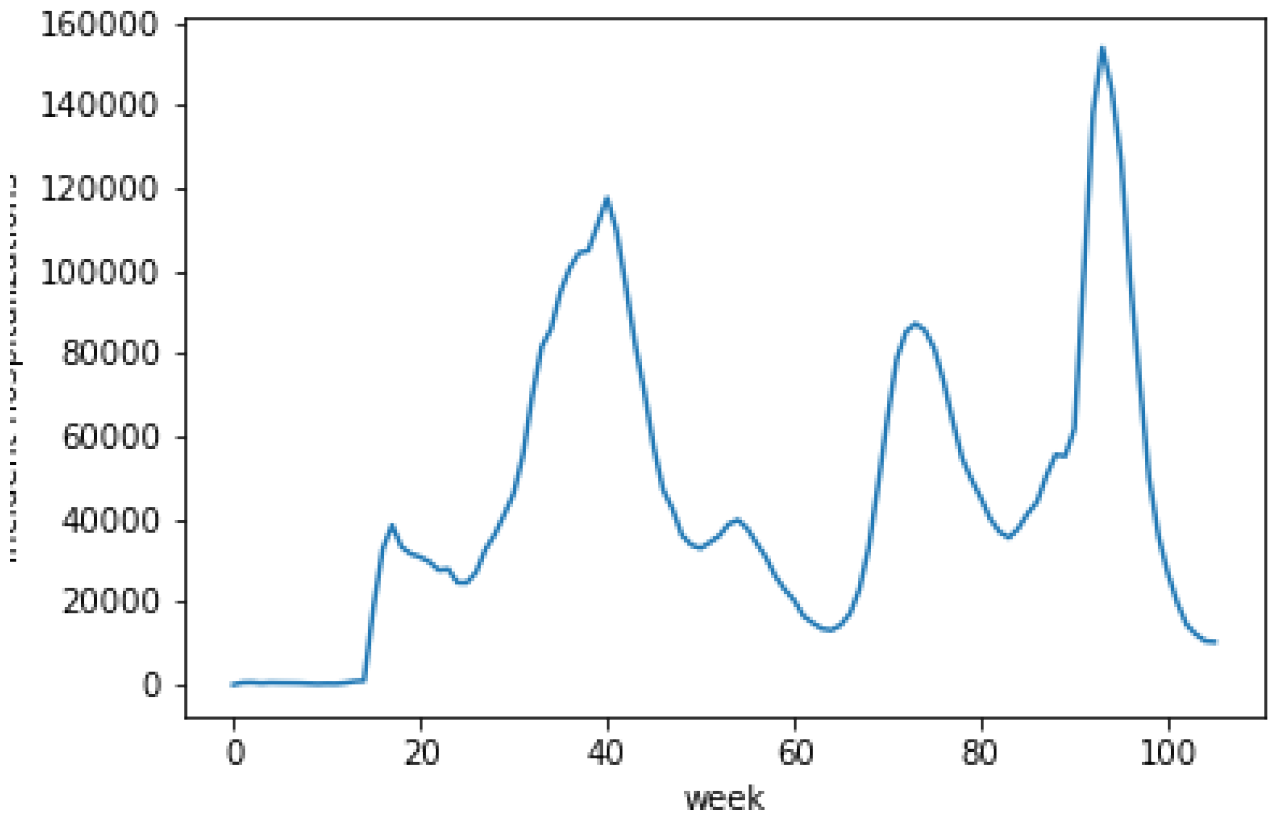
Time Series of Weekly Incident Hospitalizations.

**Figure 32.**
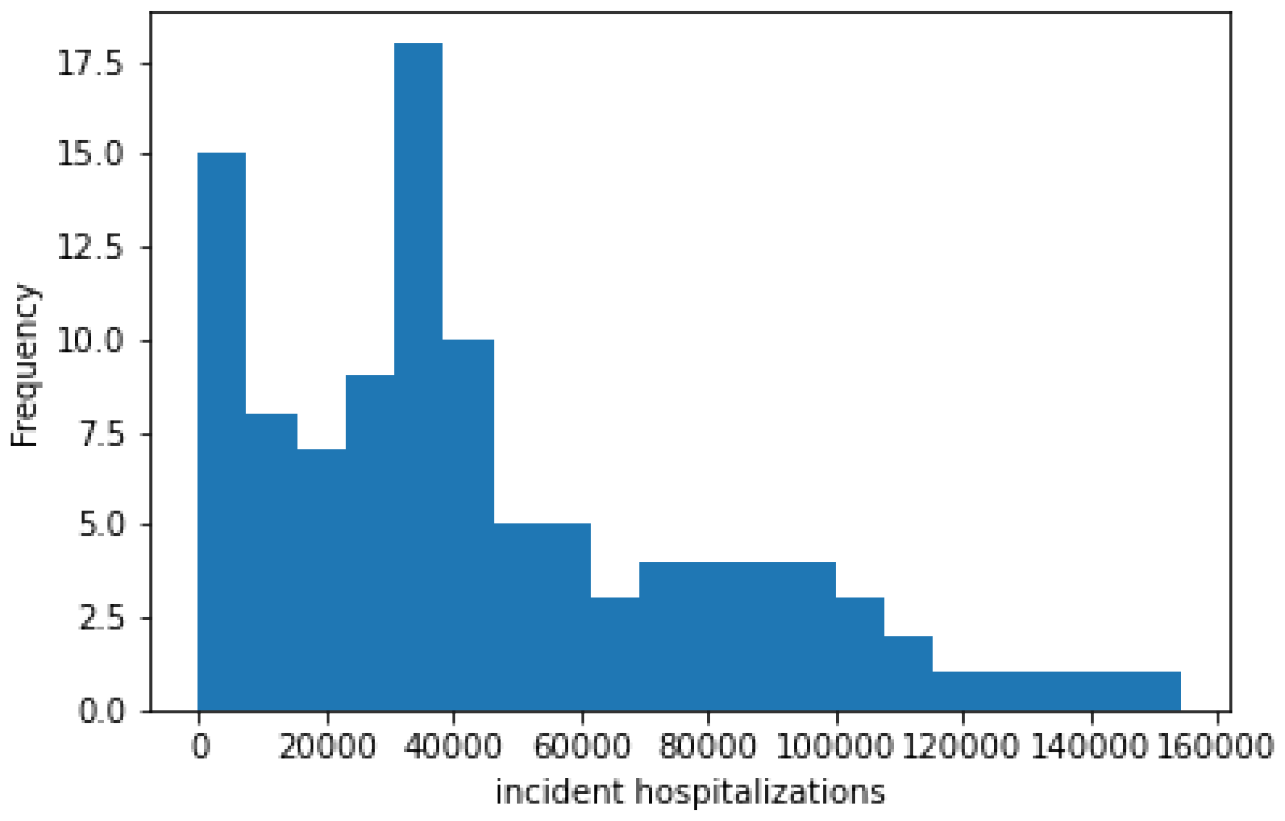
Distribution of Weekly Incident Hospitalizations.

**Figure 33.**
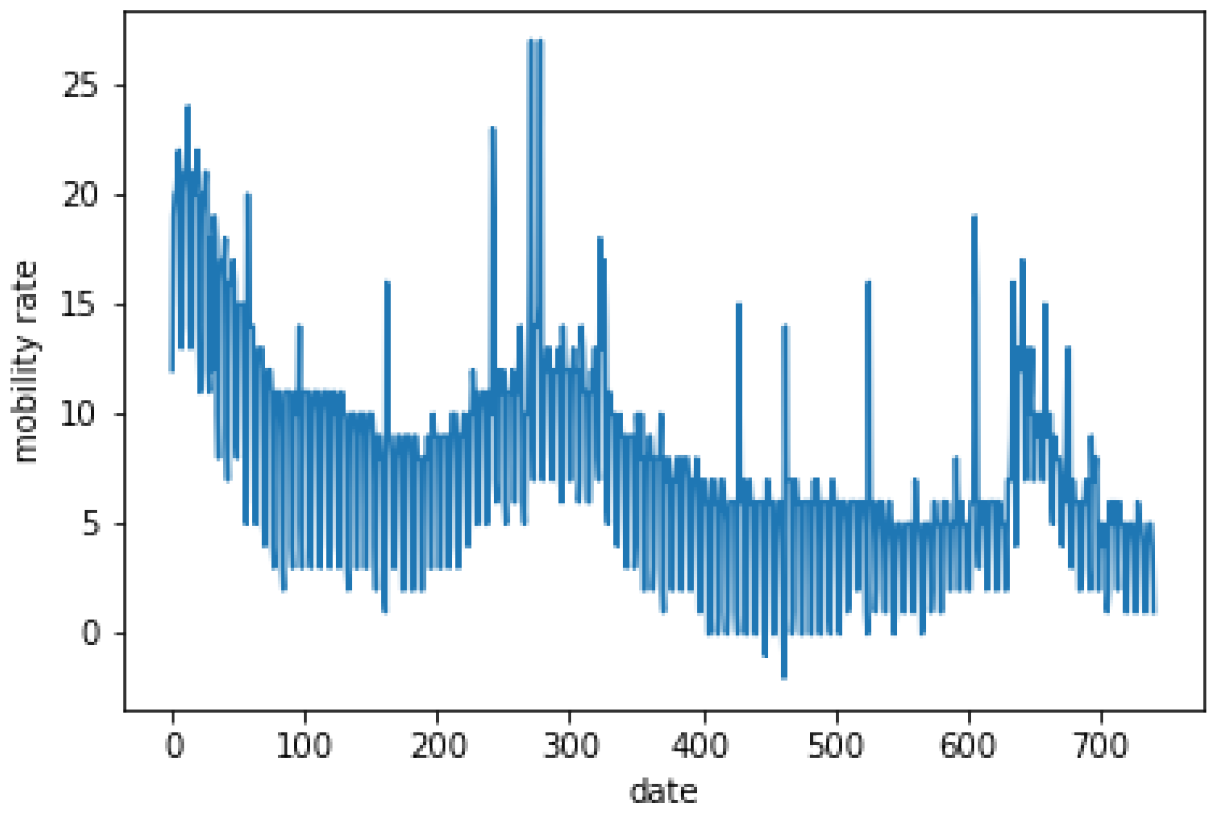
Time Series of Daily Mobility Rate.

**Figure 34.**
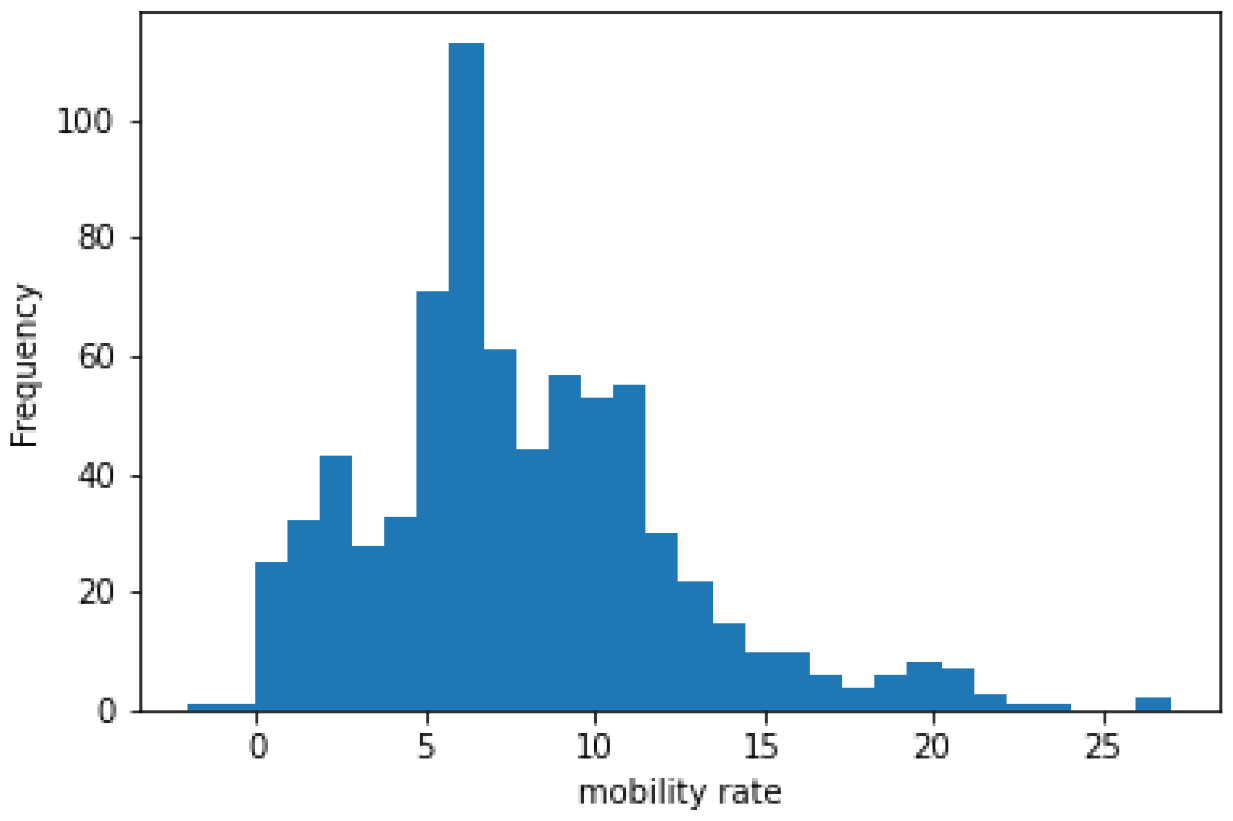
Distribution of Daily Mobility Rate.

**Figure 35.**
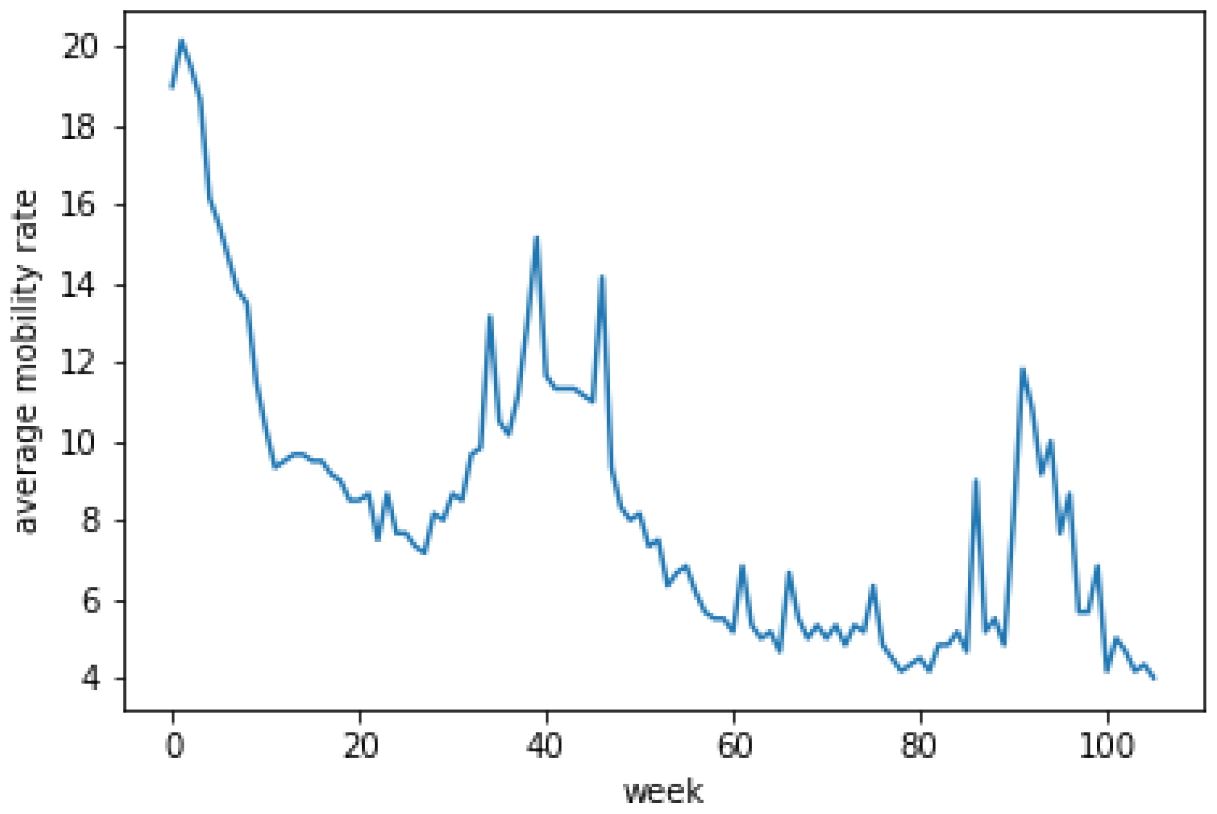
Time Series of Weekly Mobility Rate.

**Figure 36.**
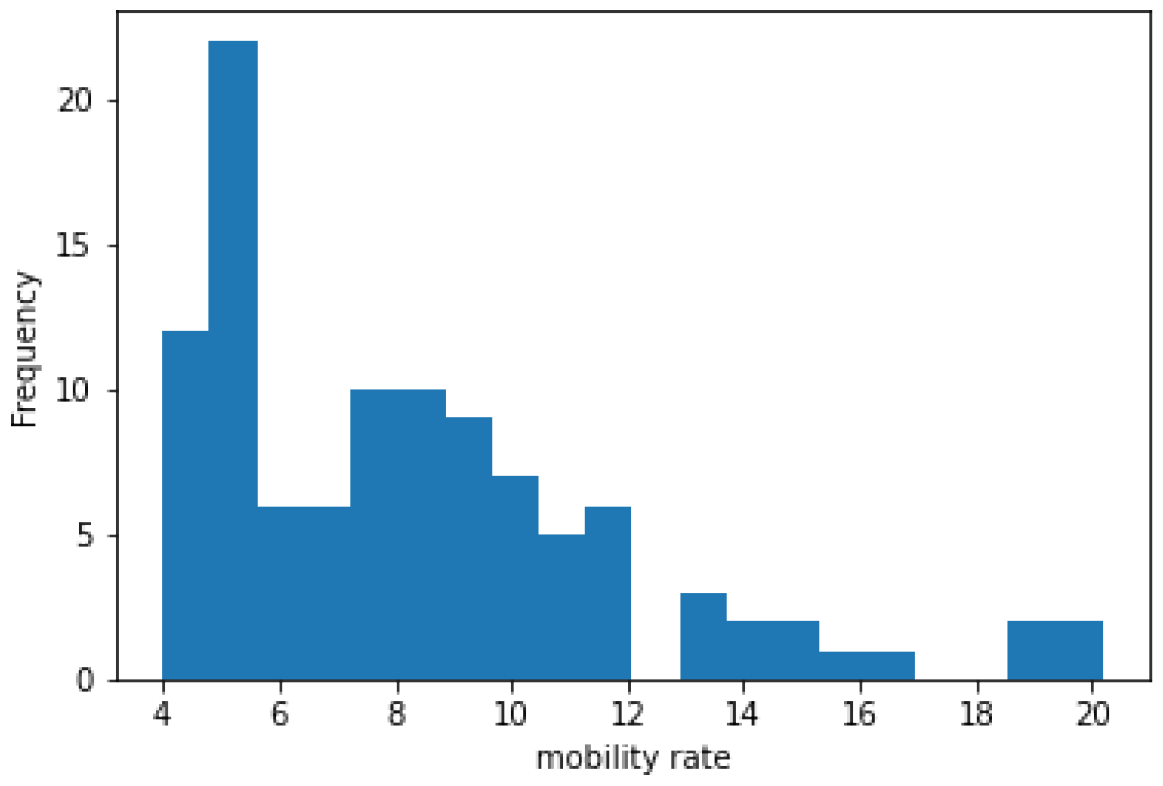
Distribution of Weekly Mobility Rate.

## Results for individual states

**New York**

**California**

**Wisconsin**

**Arizona**

**Oklahoma**

**US excluding mobility data**

**Figure 37.**
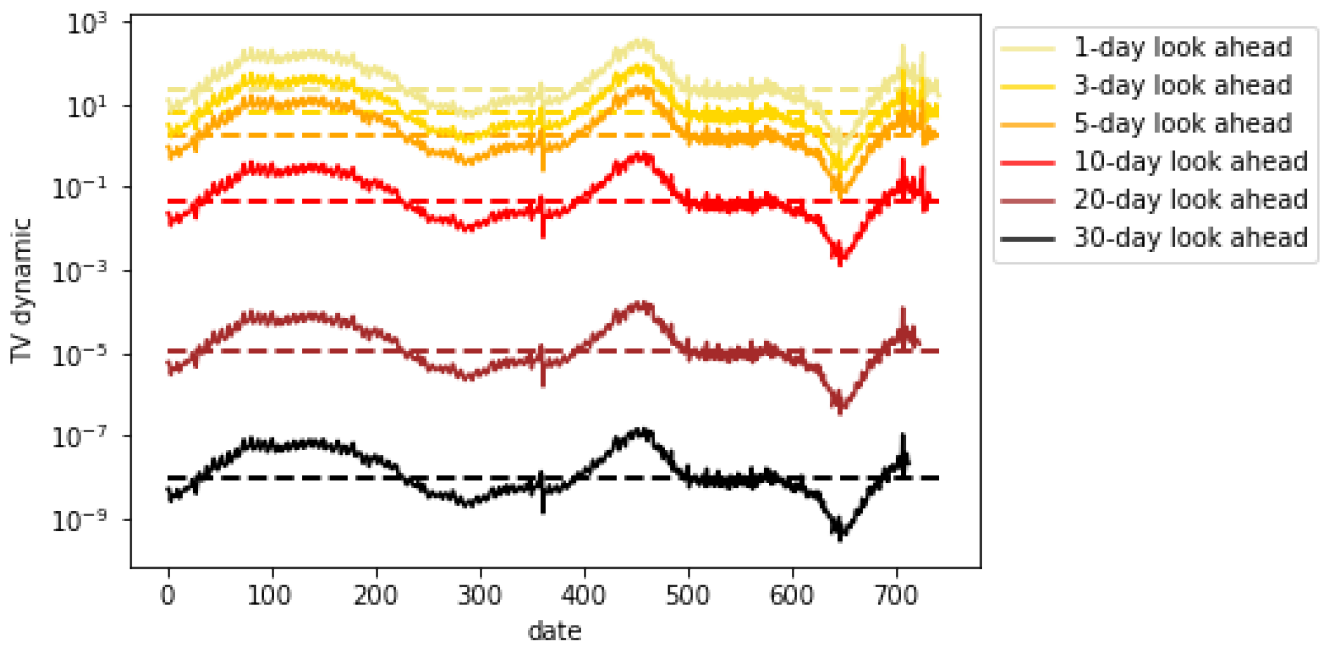
*TV* ^*dynamic*^ for daily cases (NY)

**Figure 38.**
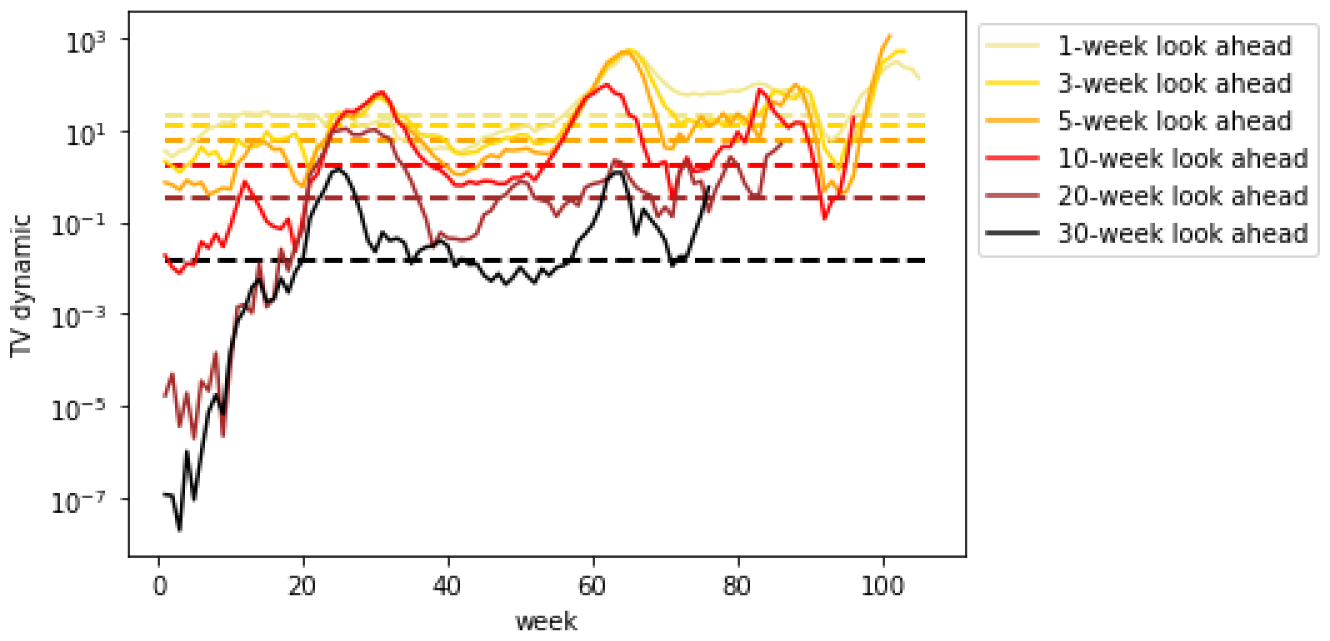
*TV* ^*dynamic*^ for weekly cases (NY)

**Figure 39.**
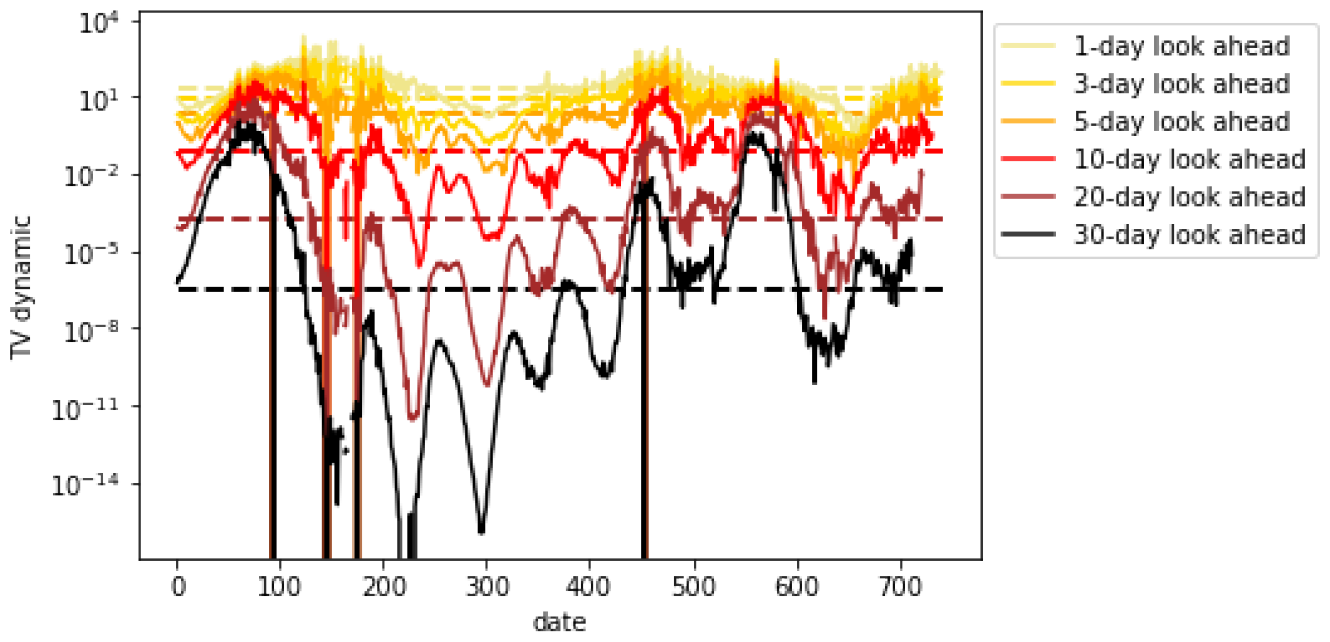
*TV* ^*dynamic*^ for daily mortality (NY)

**Figure 40.**
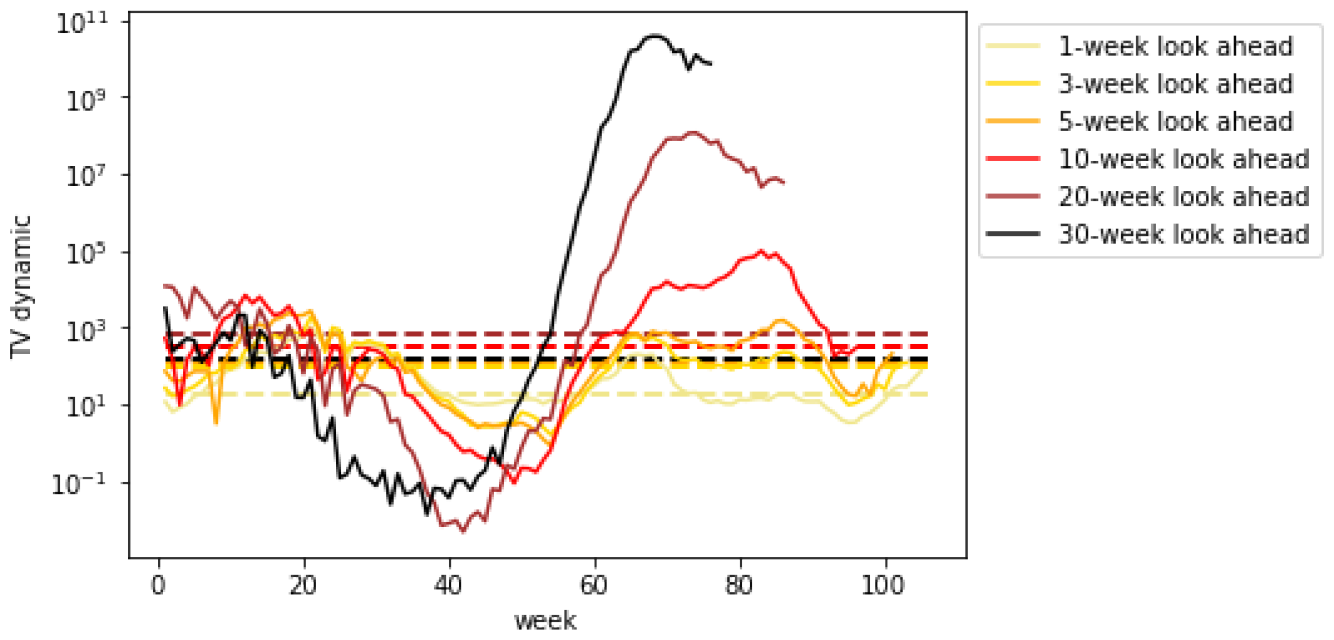
*TV* ^*dynamic*^ for weekly mortality (NY)

**Figure 41.**
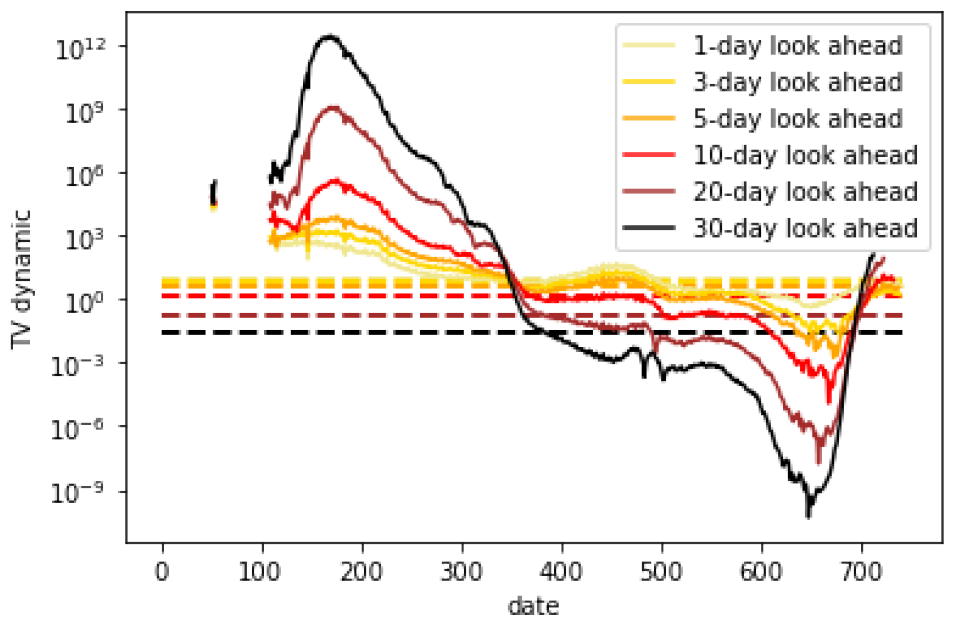
*TV* ^*dynamic*^ for daily hospitalizations (NY)

**Figure 42.**
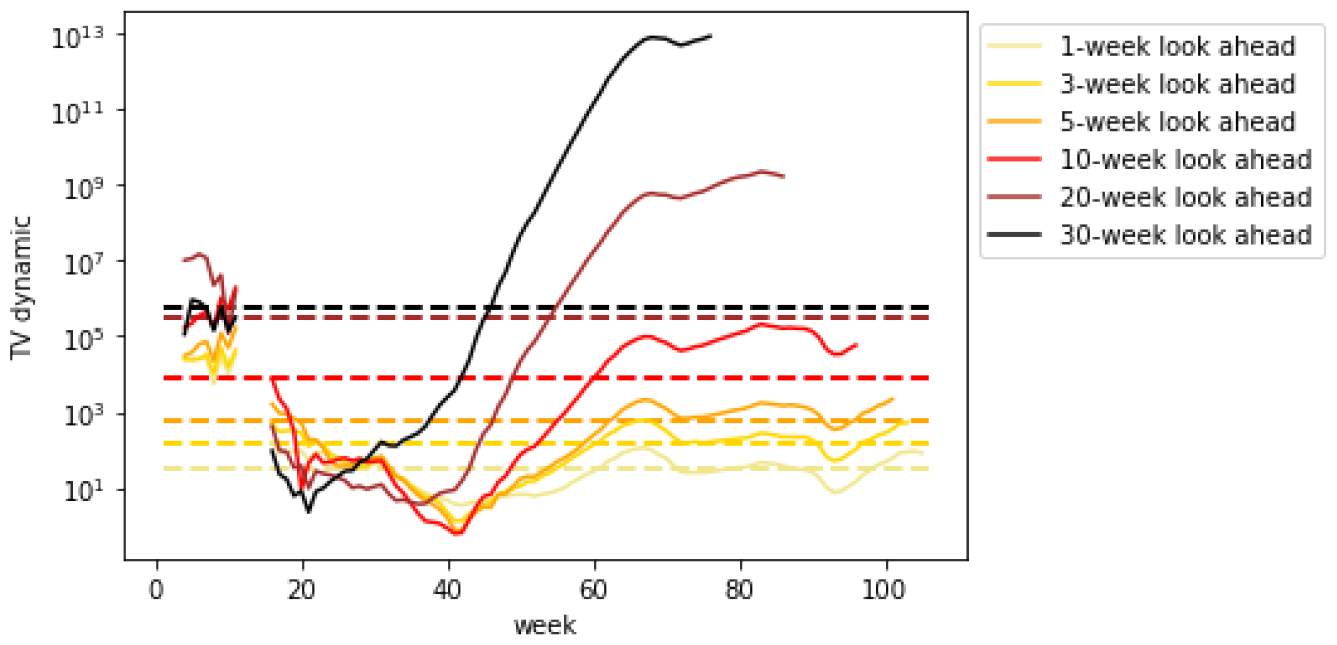
*TV* ^*dynamic*^ for weekly hospitalizations (NY)

**Figure 43.**
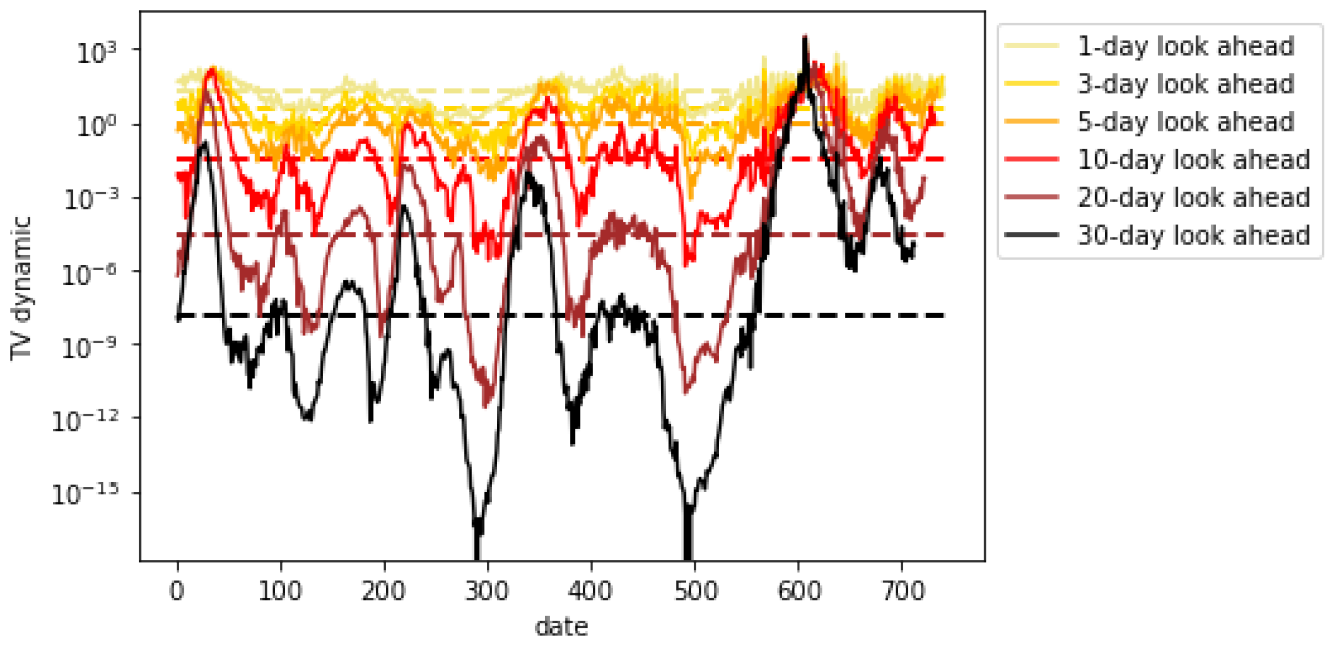
*TV* ^*dynamic*^ for daily cases (CA)

**Figure 44.**
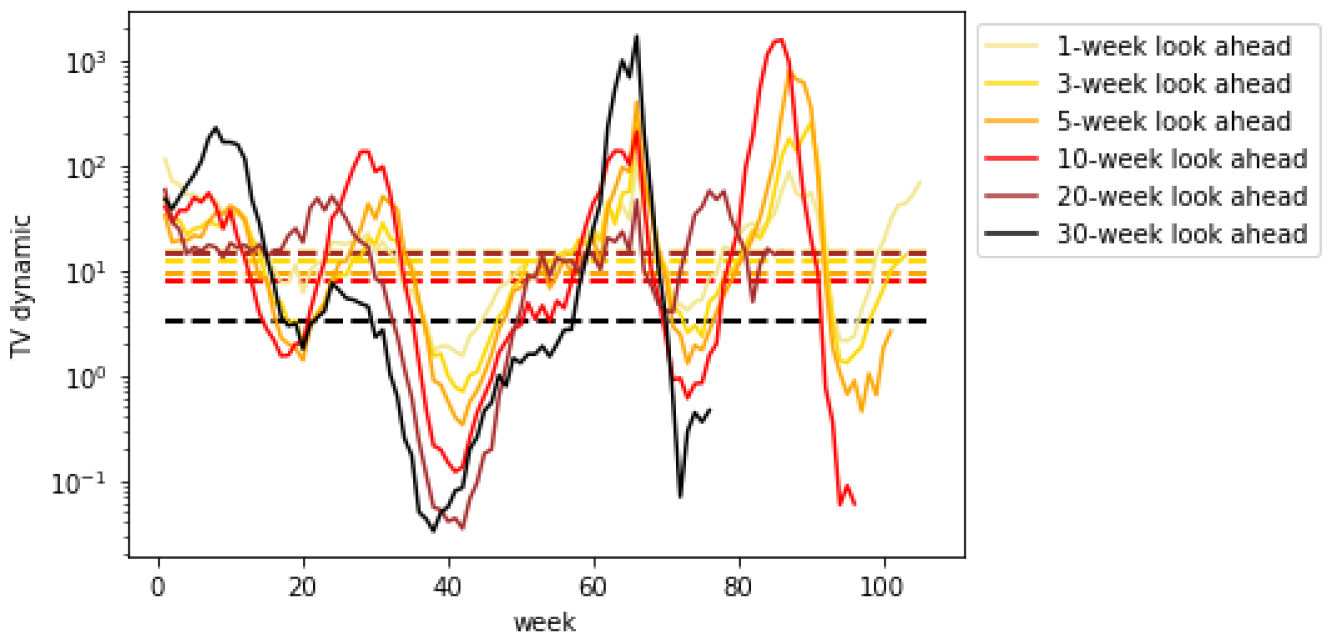
*TV* ^*dynamic*^ for weekly cases (CA)

**Figure 45.**
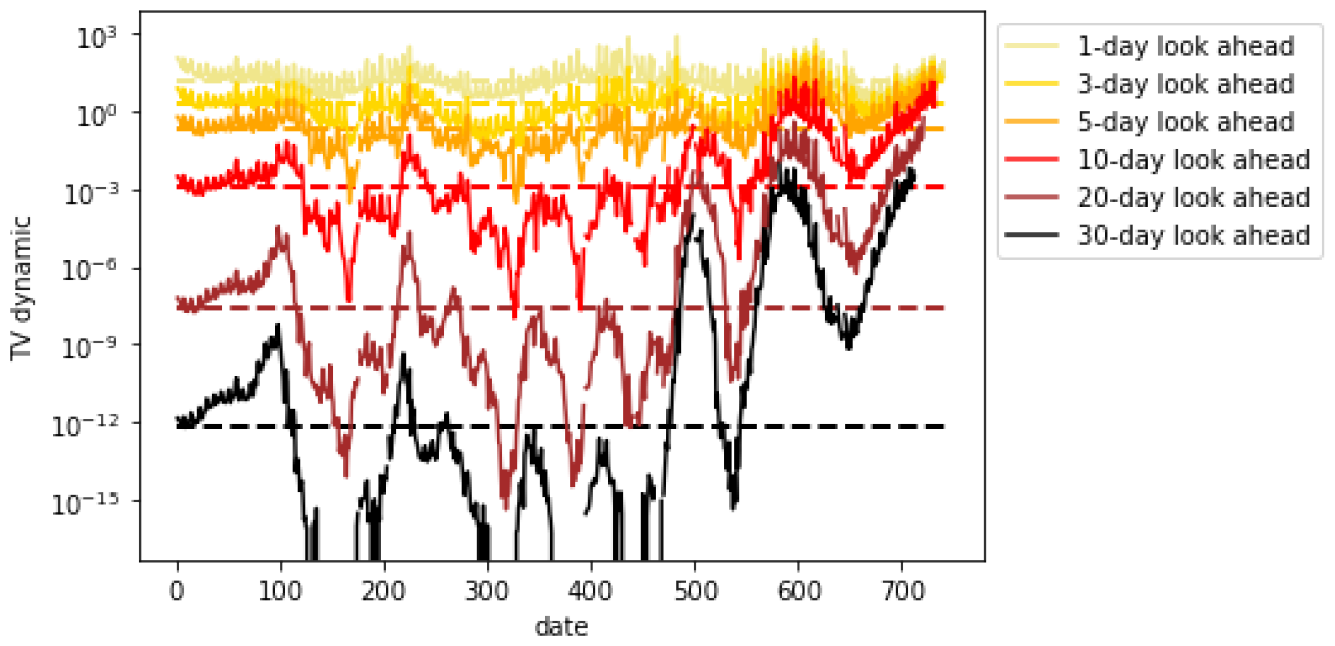
*TV* ^*dynamic*^ for daily mortality (CA)

**Figure 46.**
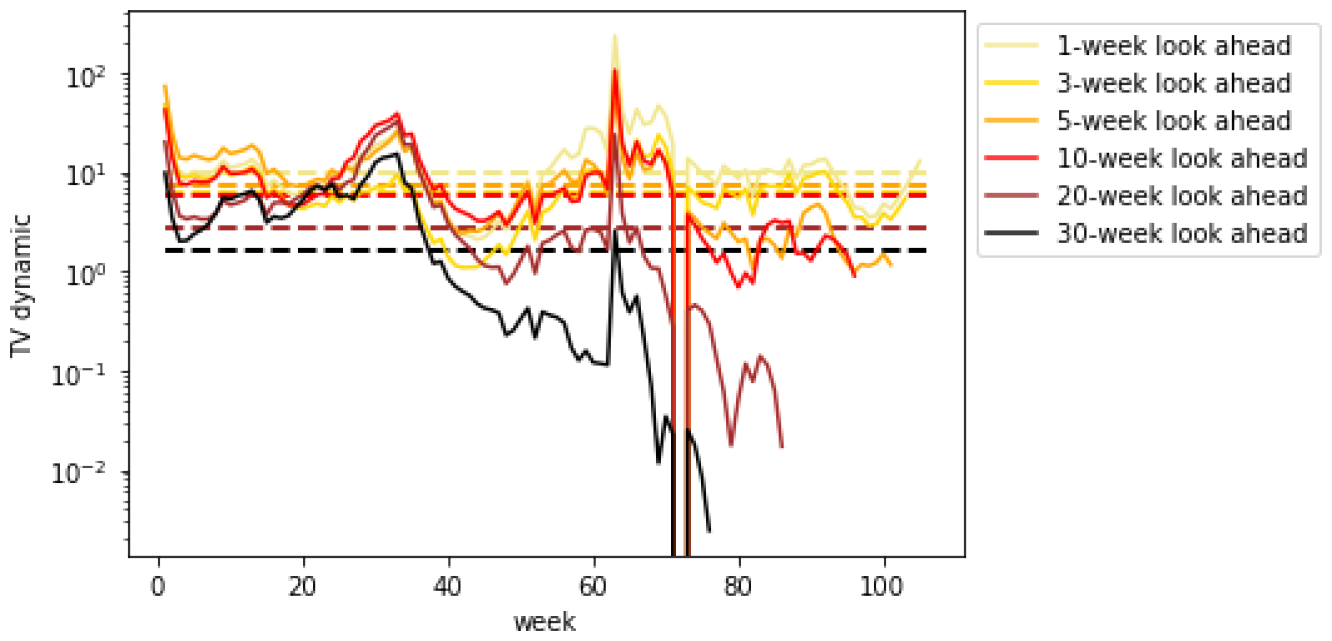
*TV* ^*dynamic*^ for weekly mortality (CA)

**Figure 47.**
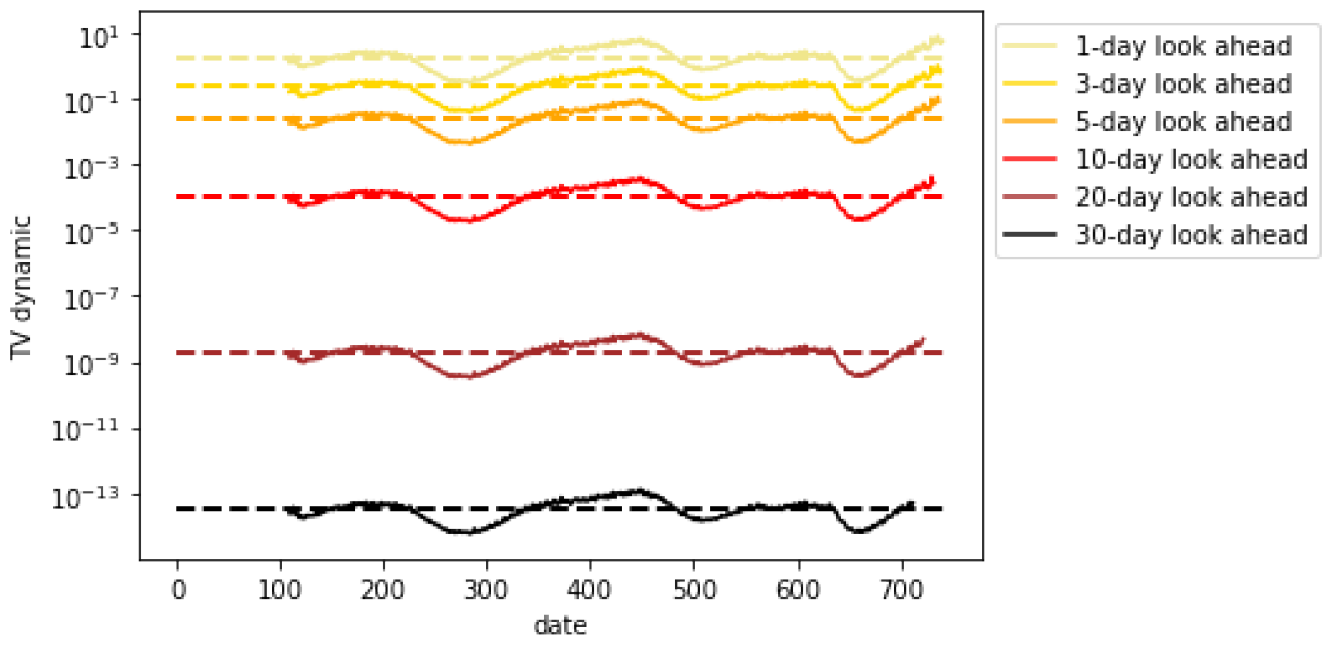
*TV* ^*dynamic*^ for daily hospitalizations (CA)

**Figure 48.**
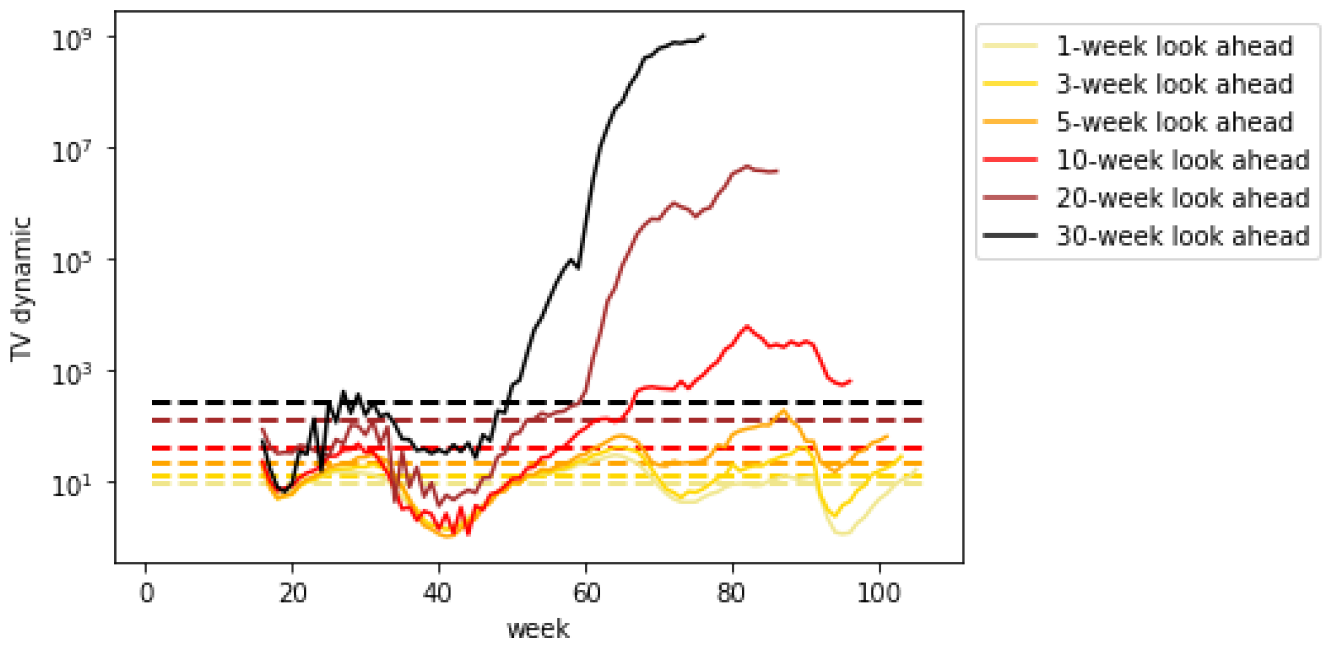
*TV* ^*dynamic*^ for weekly hospitalizations (CA)

**Figure 49.**
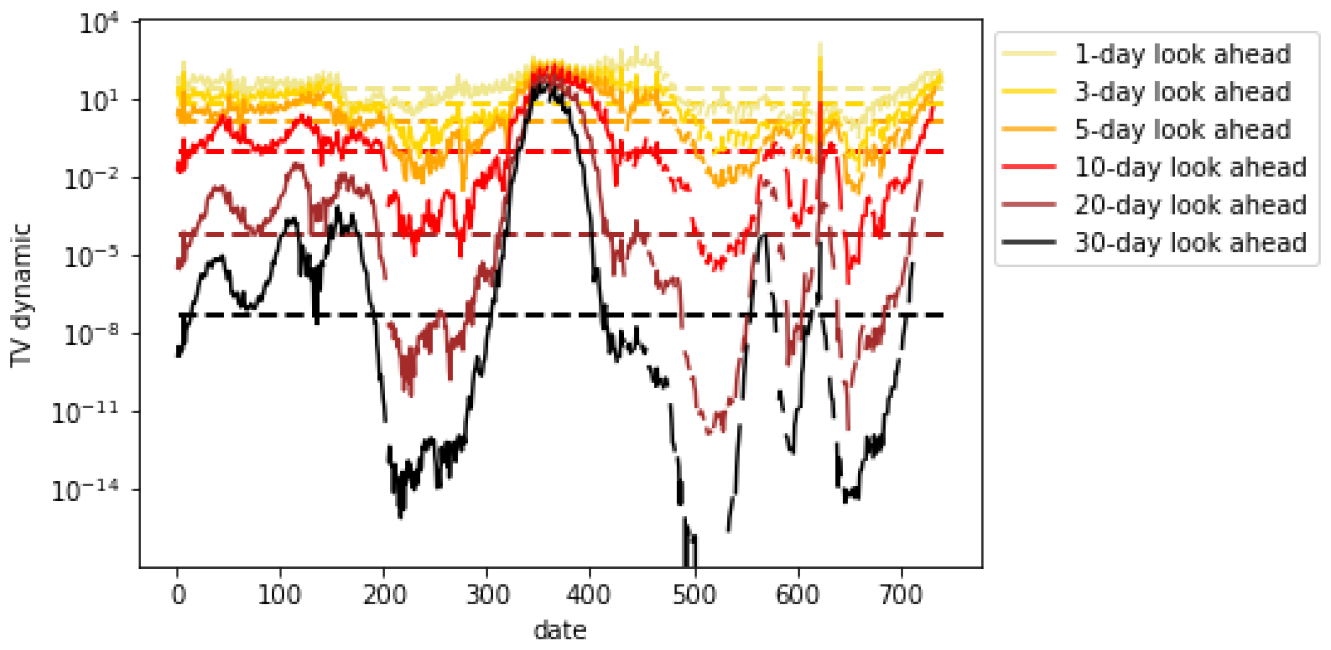
*TV* ^*dynamic*^ for daily cases (WI)

**Figure 50.**
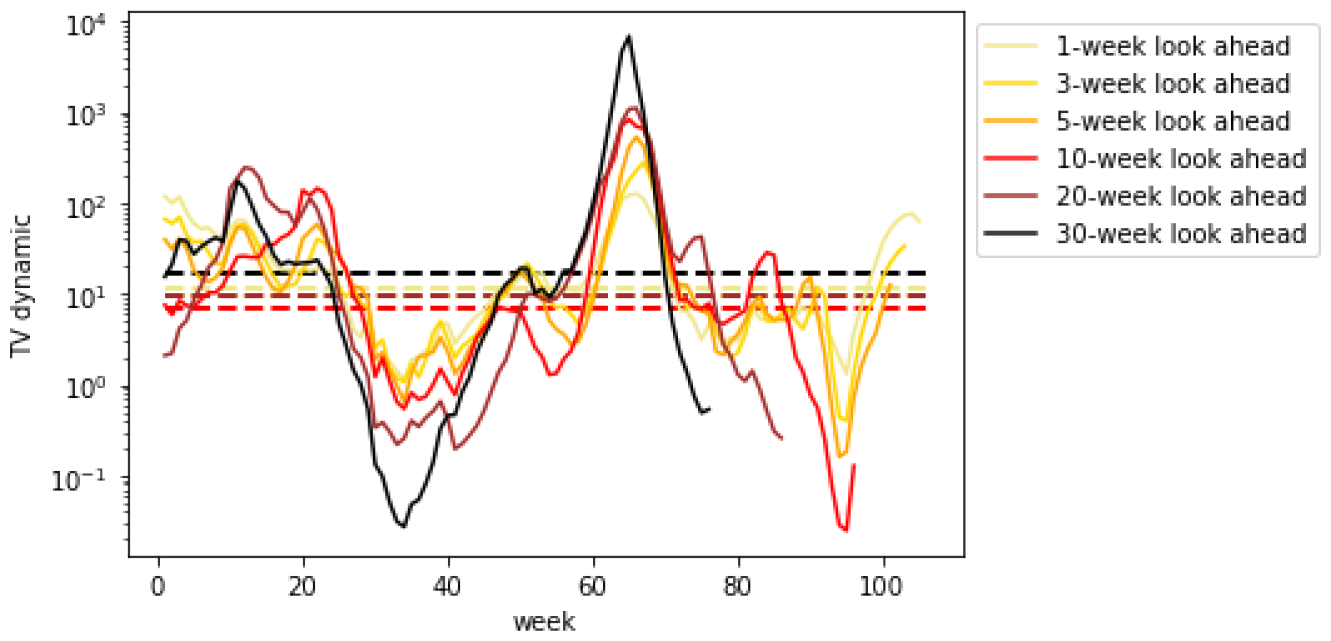
*TV* ^*dynamic*^ for weekly cases (WI)

**Figure 51.**
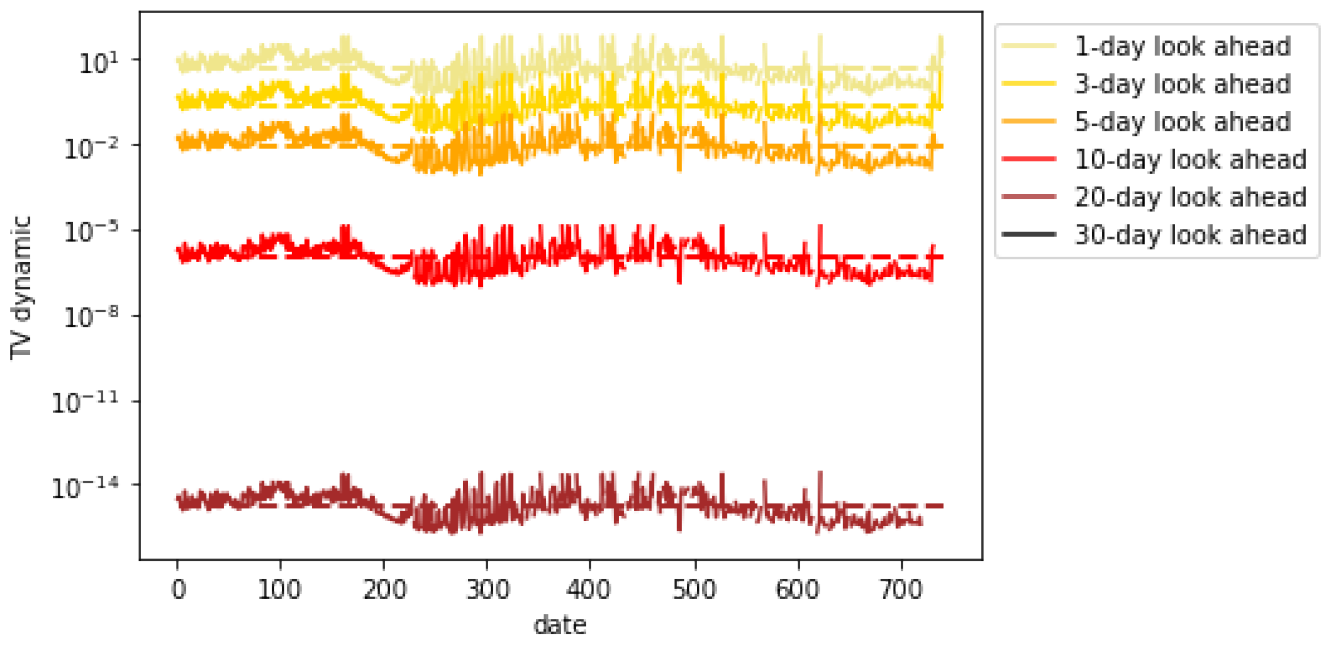
*TV* ^*dynamic*^ for daily mortality (WI)

**Figure 52.**
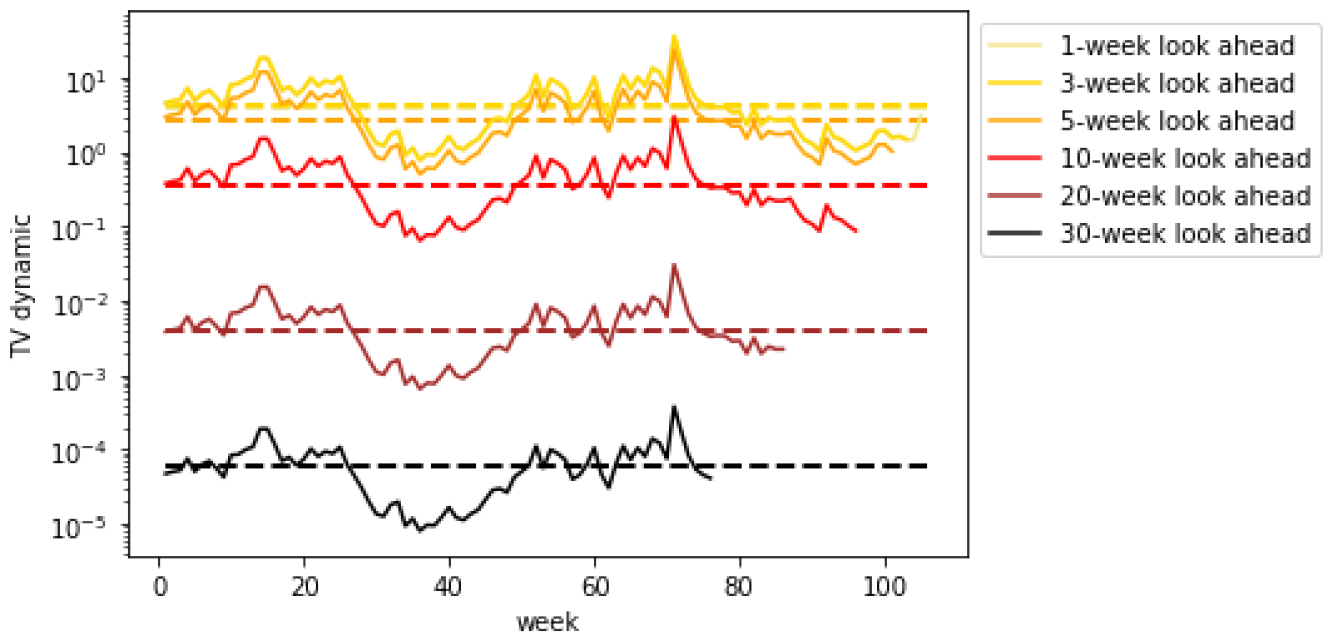
*TV* ^*dynamic*^ for weekly mortality (WI)

**Figure 53.**
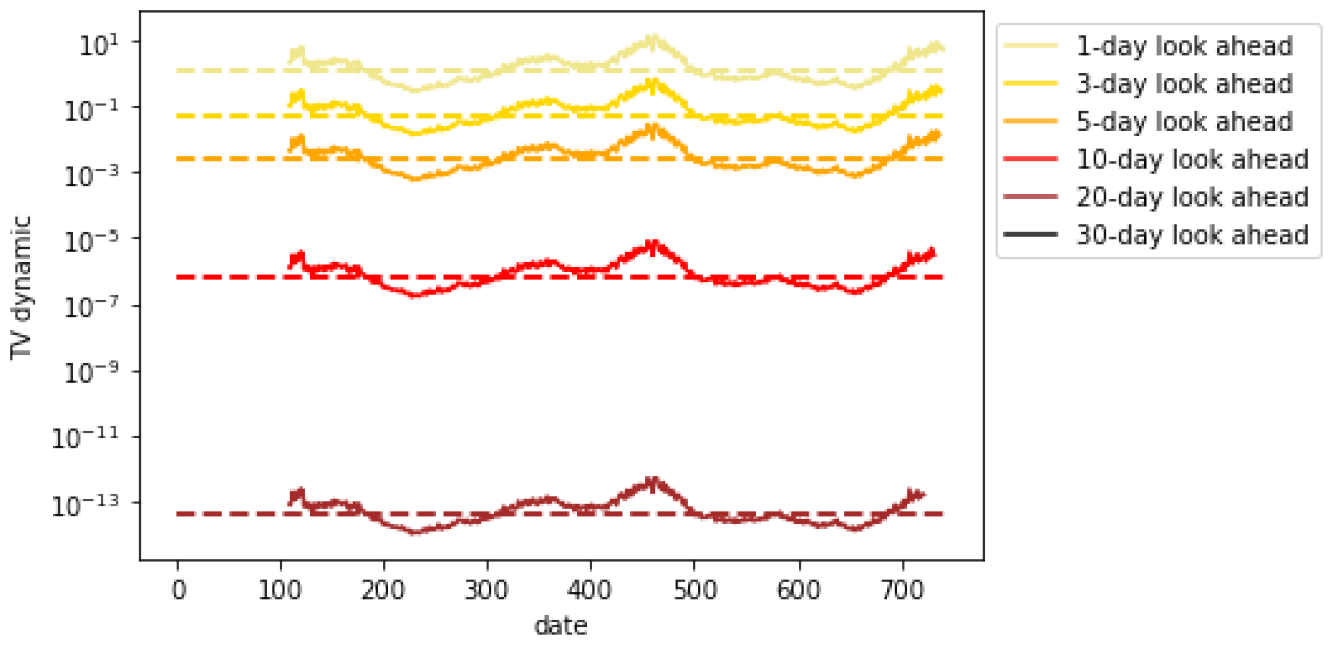
*TV* ^*dynamic*^ for daily hospitalizations (WI)

**Figure 54.**
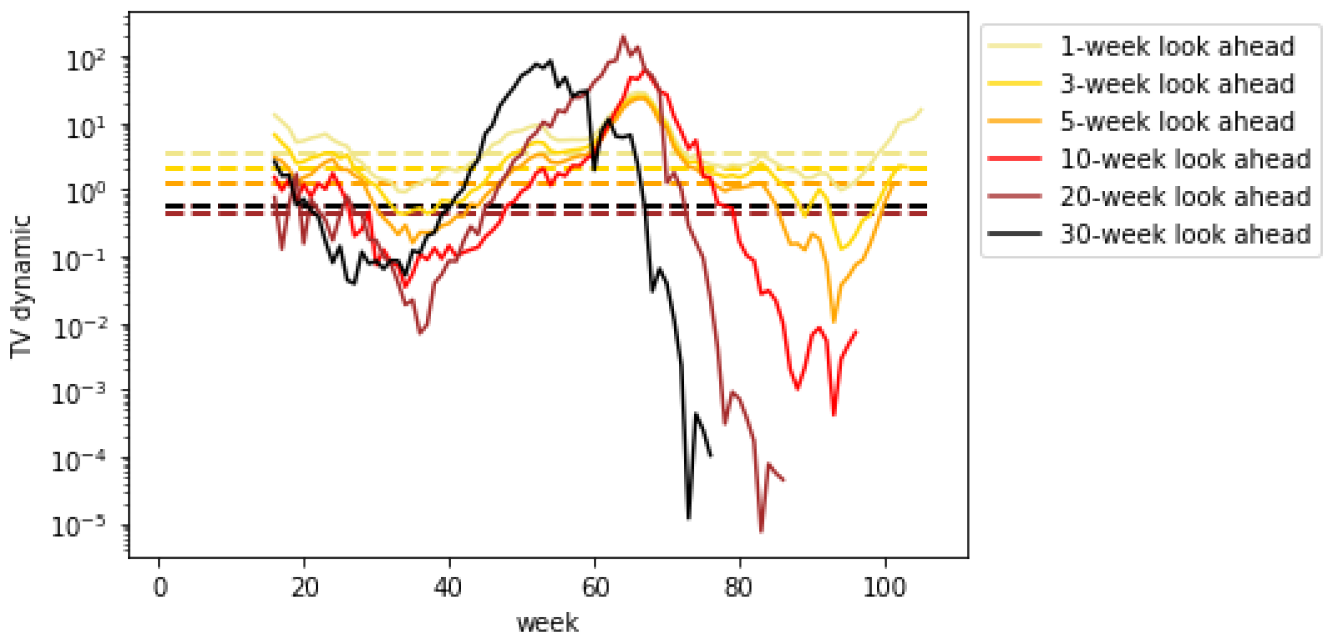
*TV* ^*dynamic*^ for weekly hospitalizations (WI)

**Figure 55.**
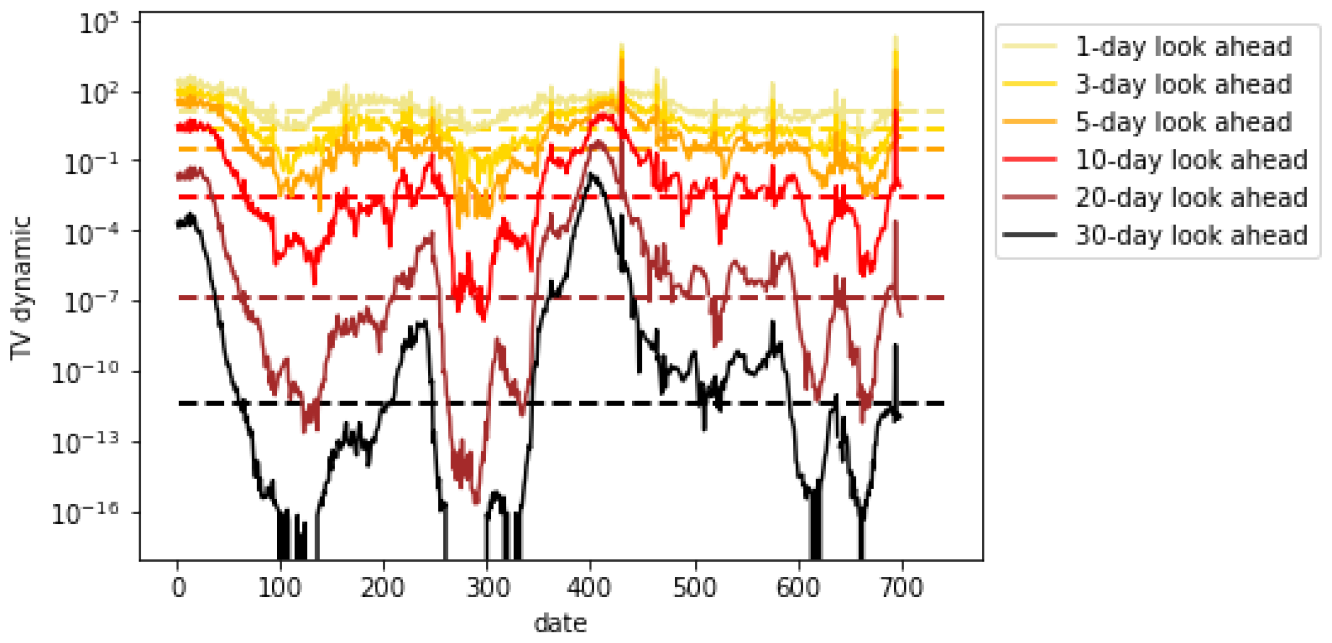
*TV* ^*dynamic*^ for daily cases (AZ)

**Figure 56.**
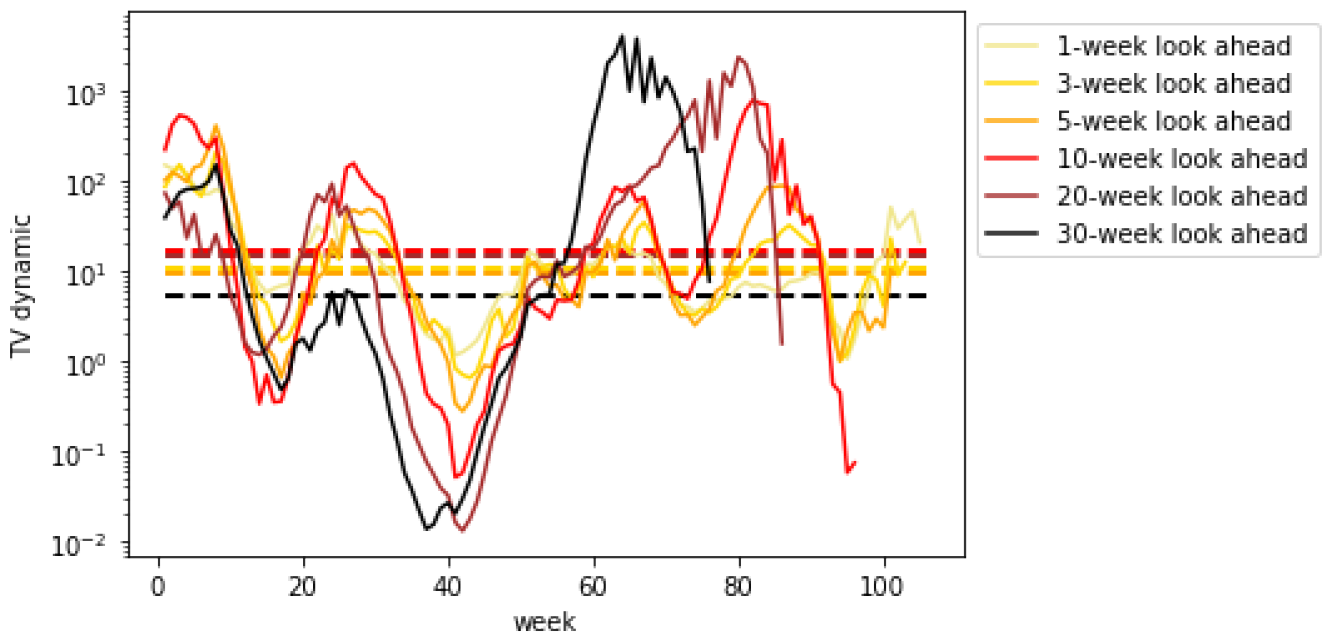
*TV* ^*dynamic*^ for weekly cases (AZ)

**Figure 57.**
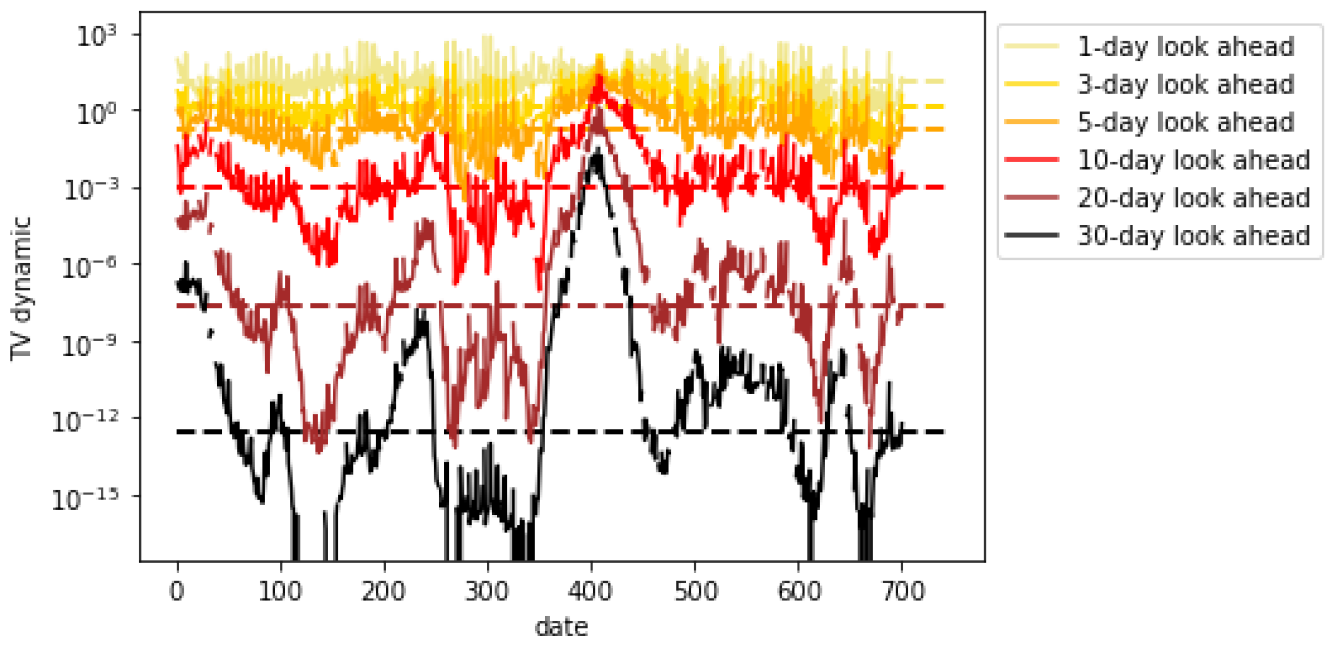
*TV* ^*dynamic*^ for daily mortality (AZ)

**Figure 58.**
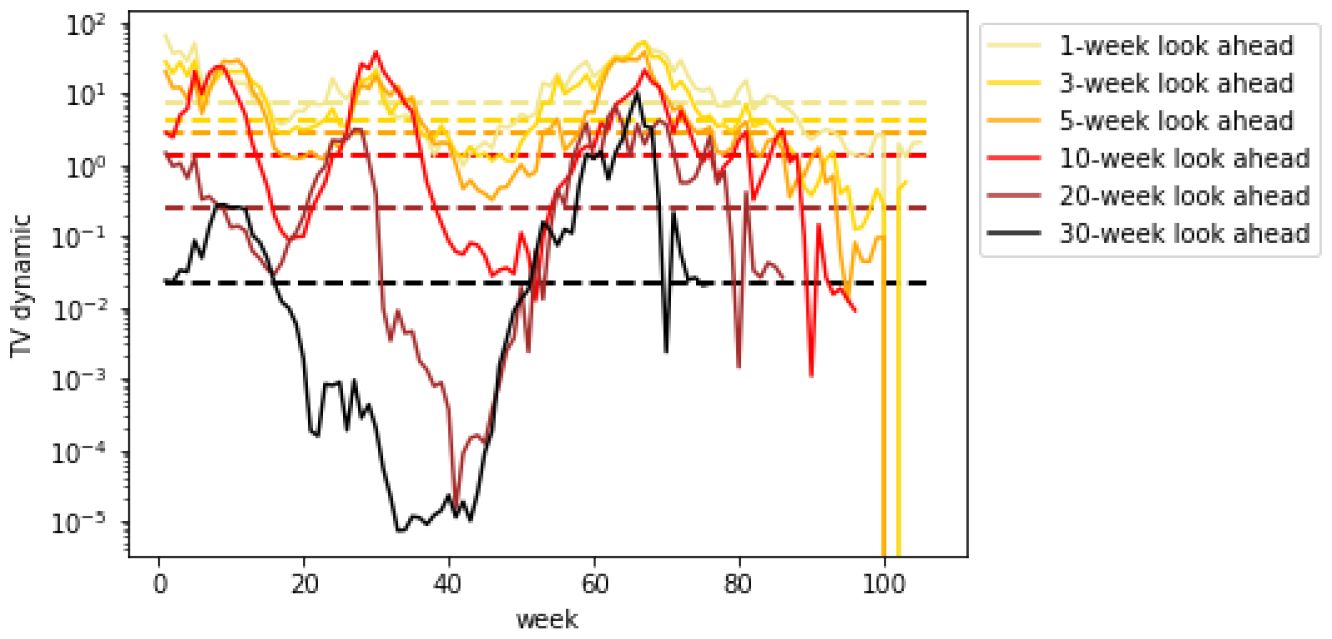
*TV* ^*dynamic*^ for weekly mortality (AZ)

**Figure 59.**
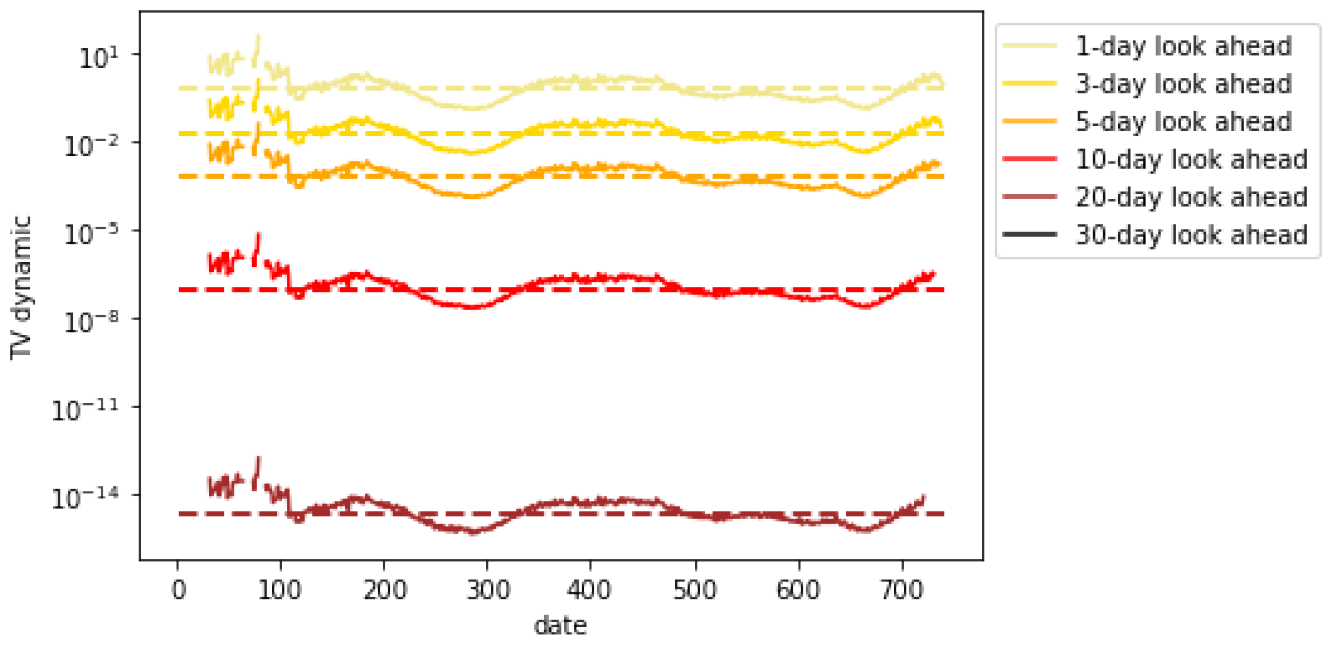
*TV* ^*dynamic*^ for daily hospitalizations (AZ)

**Figure 60.**
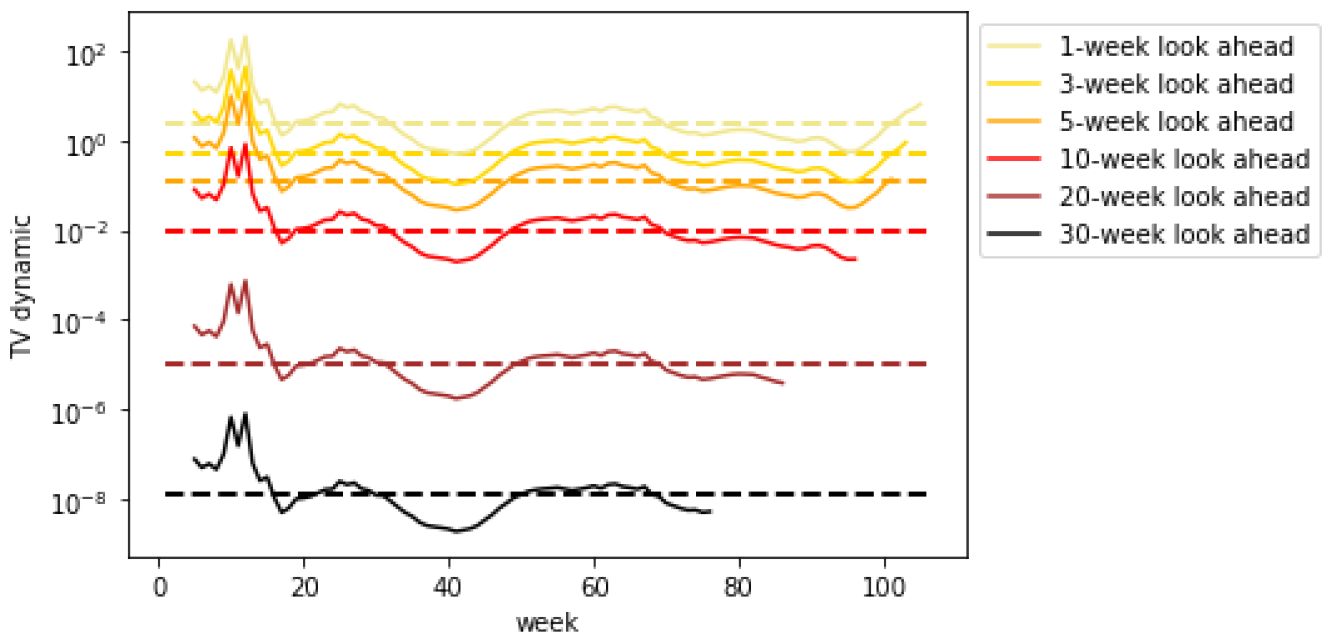
*TV* ^*dynamic*^ for weekly hospitalizations (AZ)

**Figure 61.**
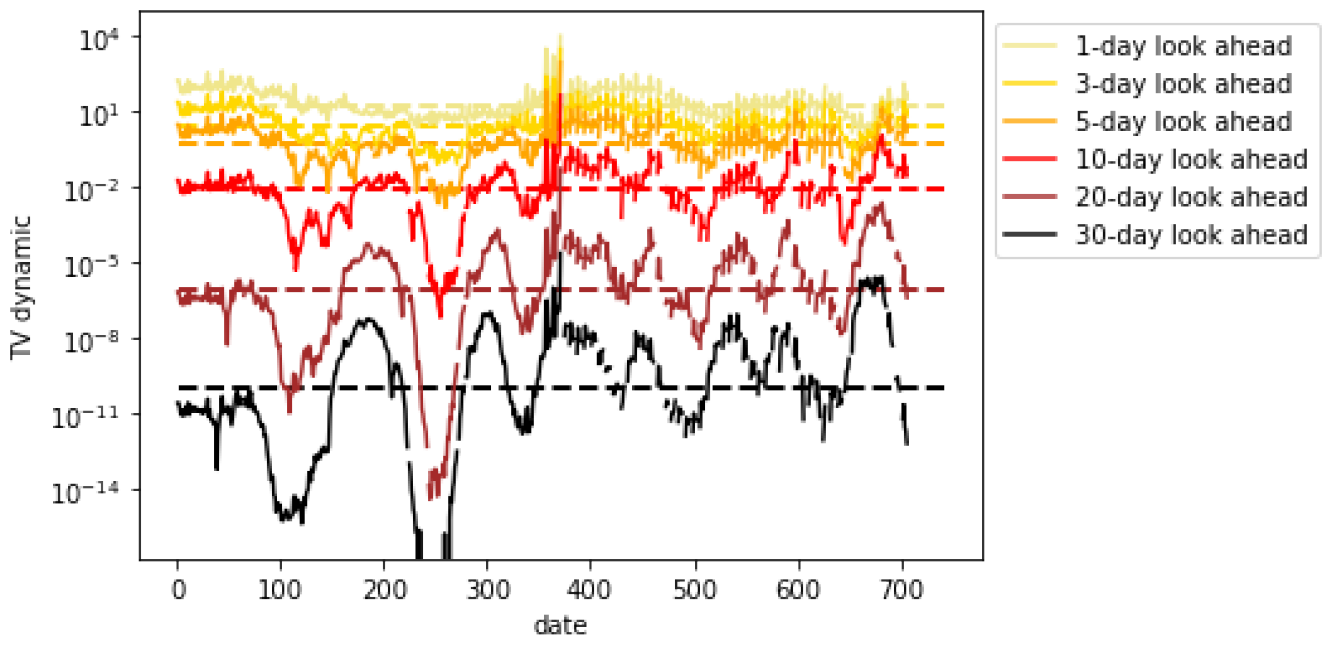
*TV* ^*dynamic*^ for daily cases (OK)

**Figure 62.**
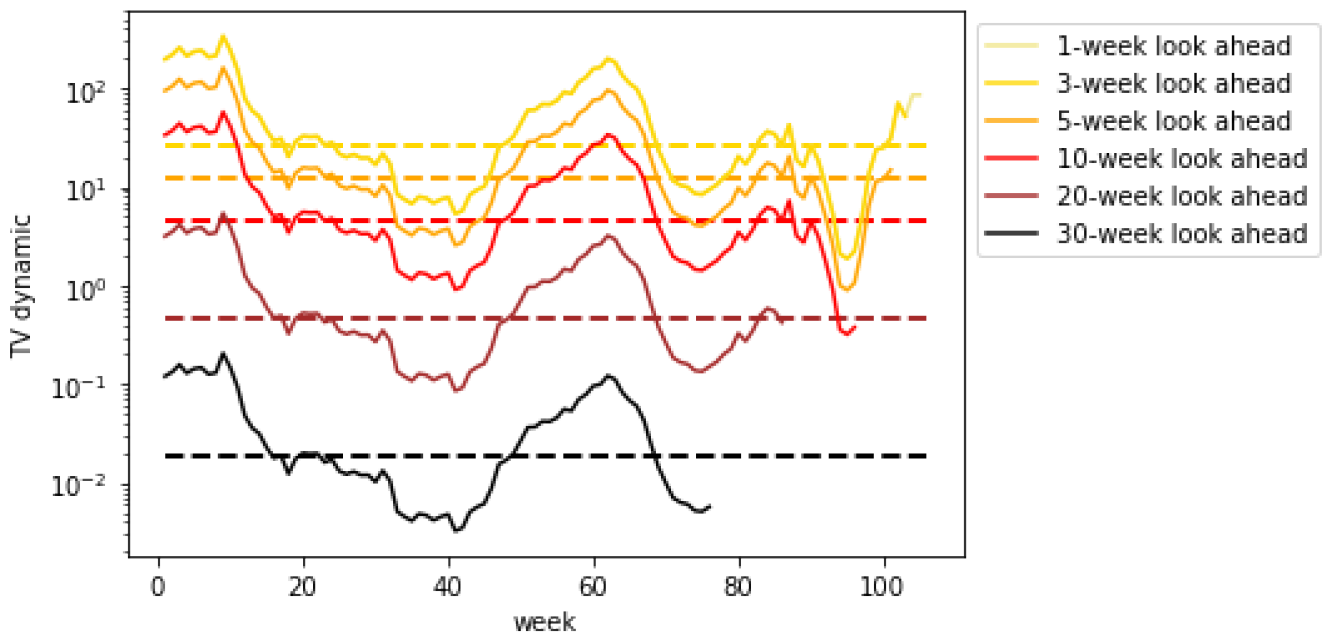
*TV* ^*dynamic*^ for weekly cases (OK)

**Figure 63.**
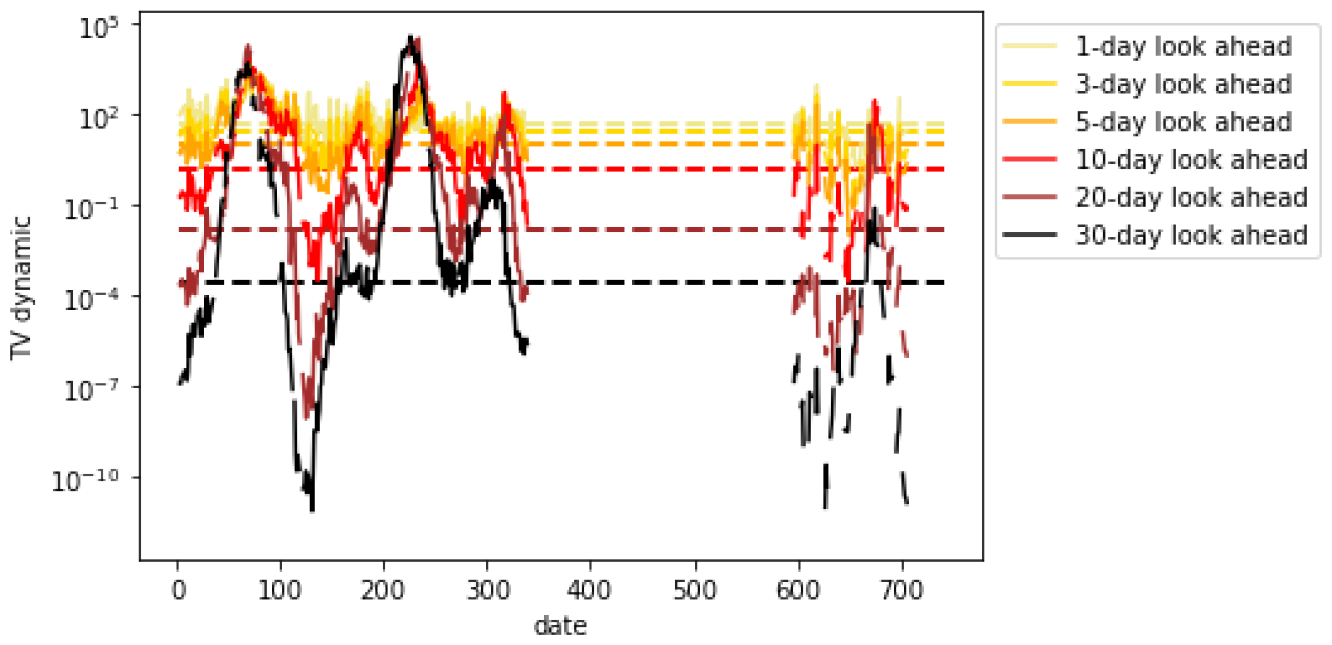
*TV* ^*dynamic*^ for daily mortality (OK)

**Figure 64.**
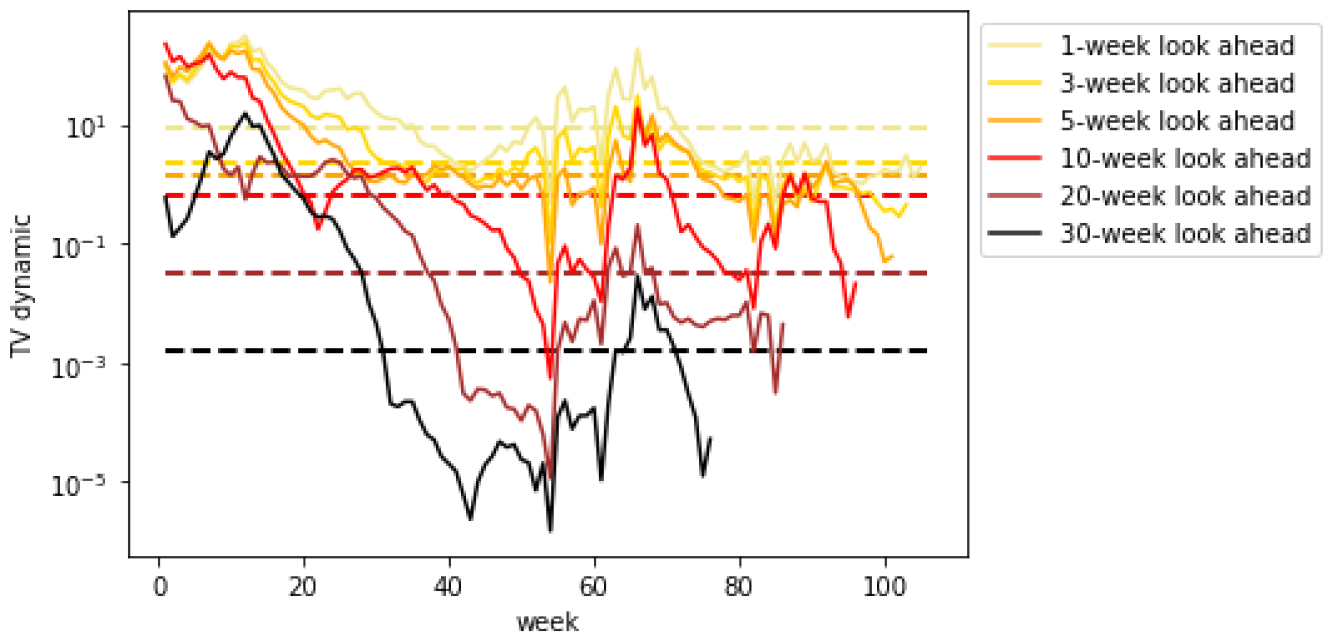
*TV* ^*dynamic*^ for weekly mortality (OK)

**Figure 65.**
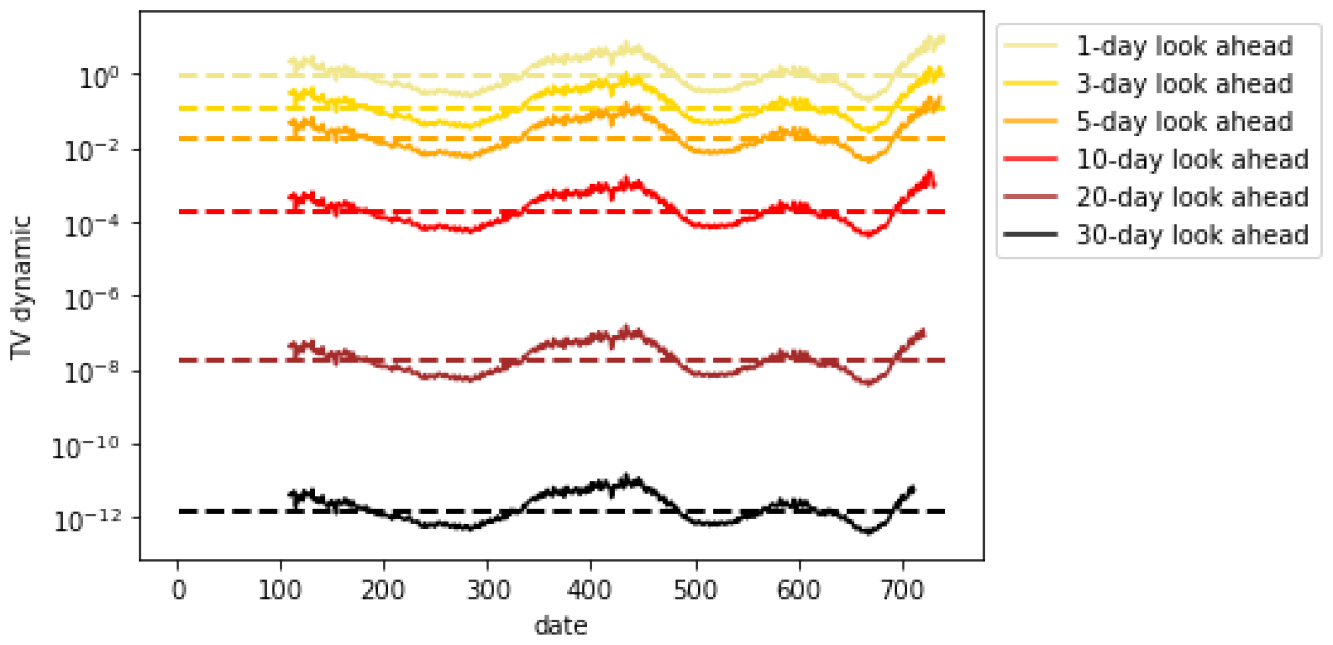
*TV* ^*dynamic*^ for daily hospitalizations (OK)

**Figure 66.**
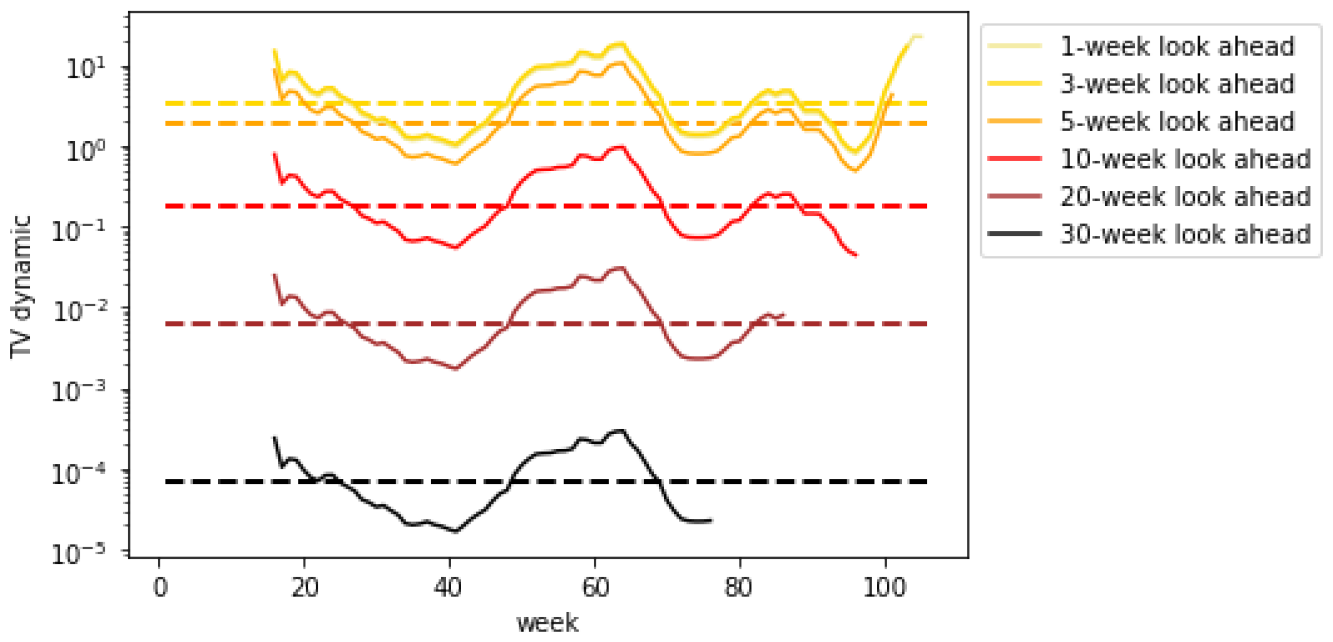
*TV* ^*dynamic*^ for weekly hospitalizations (OK)

**Figure 67.**
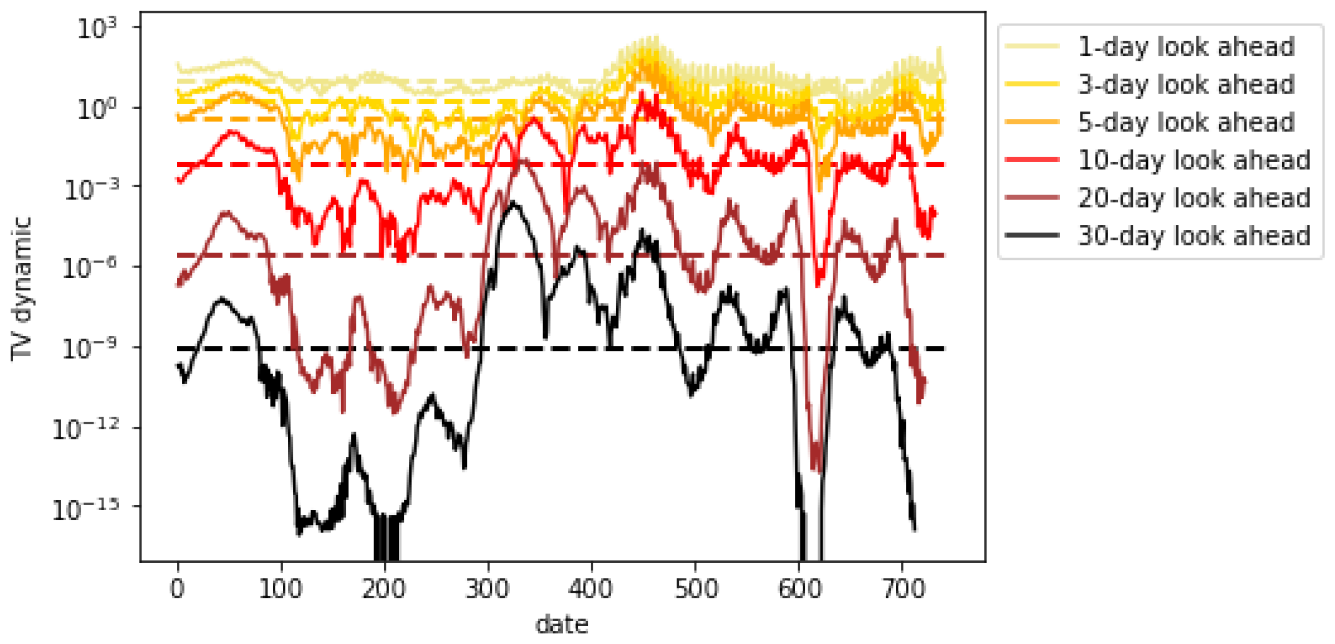
*TV* ^*dynamic*^ for daily cases (US excluding mobility data)

**Figure 68.**
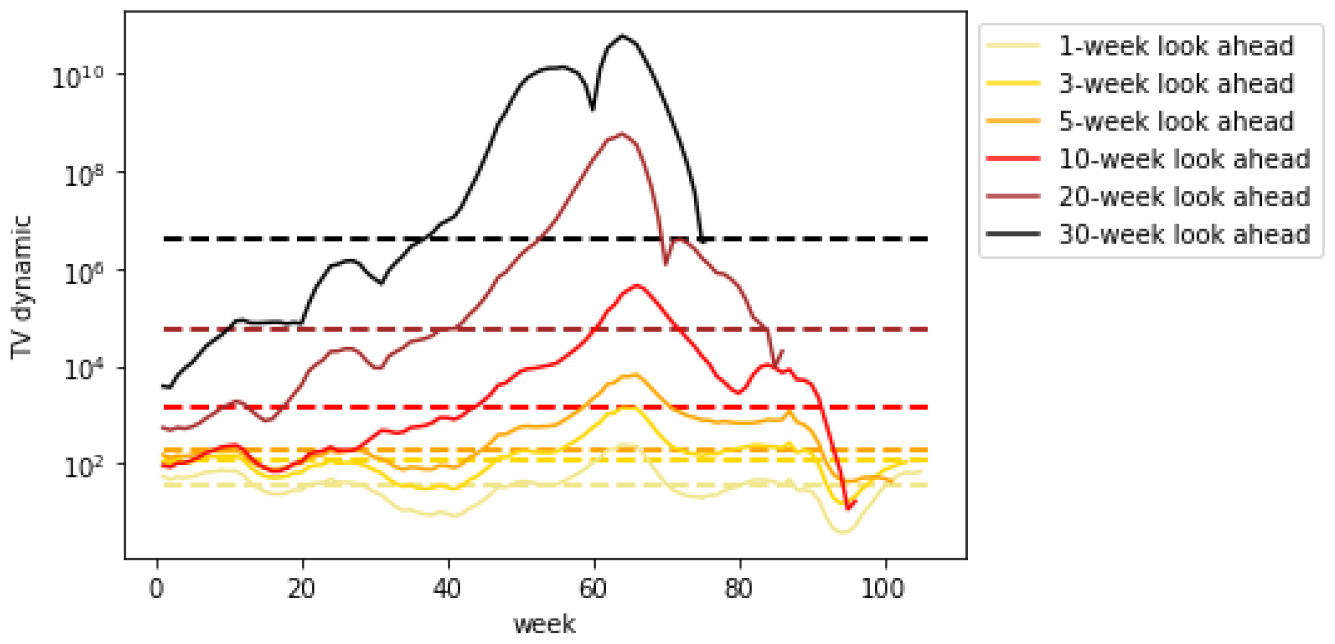
*TV* ^*dynamic*^ for weekly cases (US excluding mobility data)

**Figure 69.**
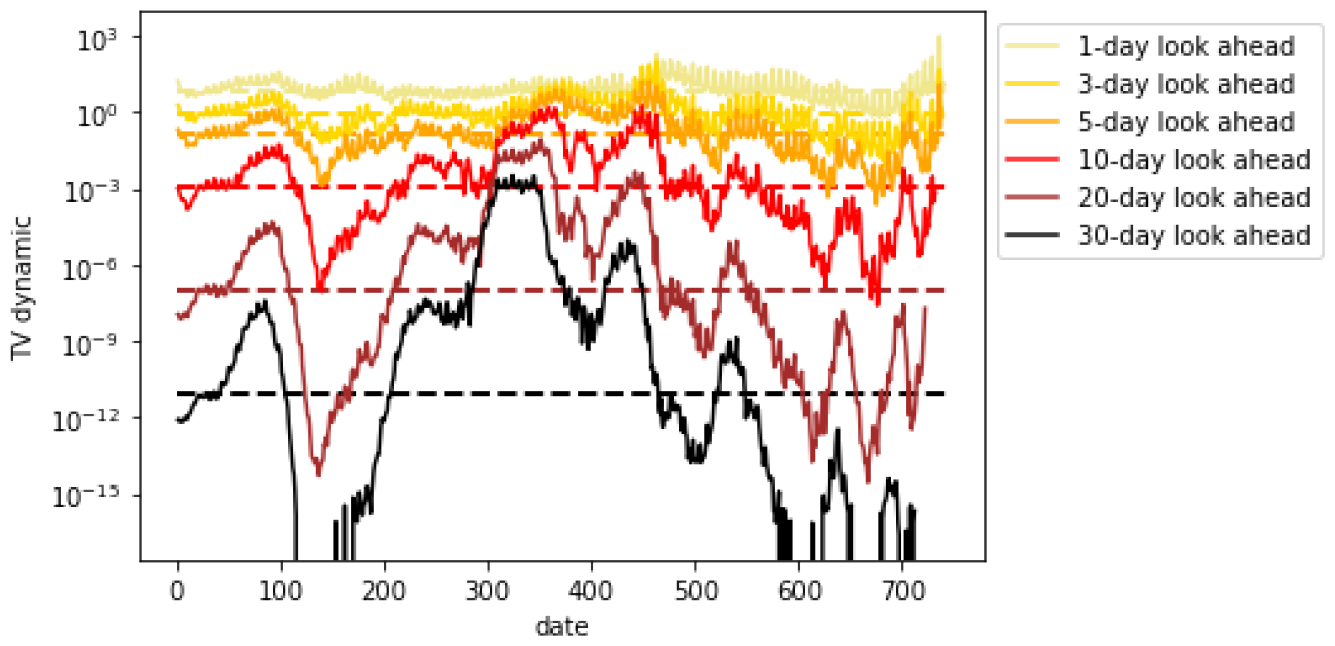
*TV* ^*dynamic*^ for daily mortality (US excluding mobility data)

**Figure 70.**
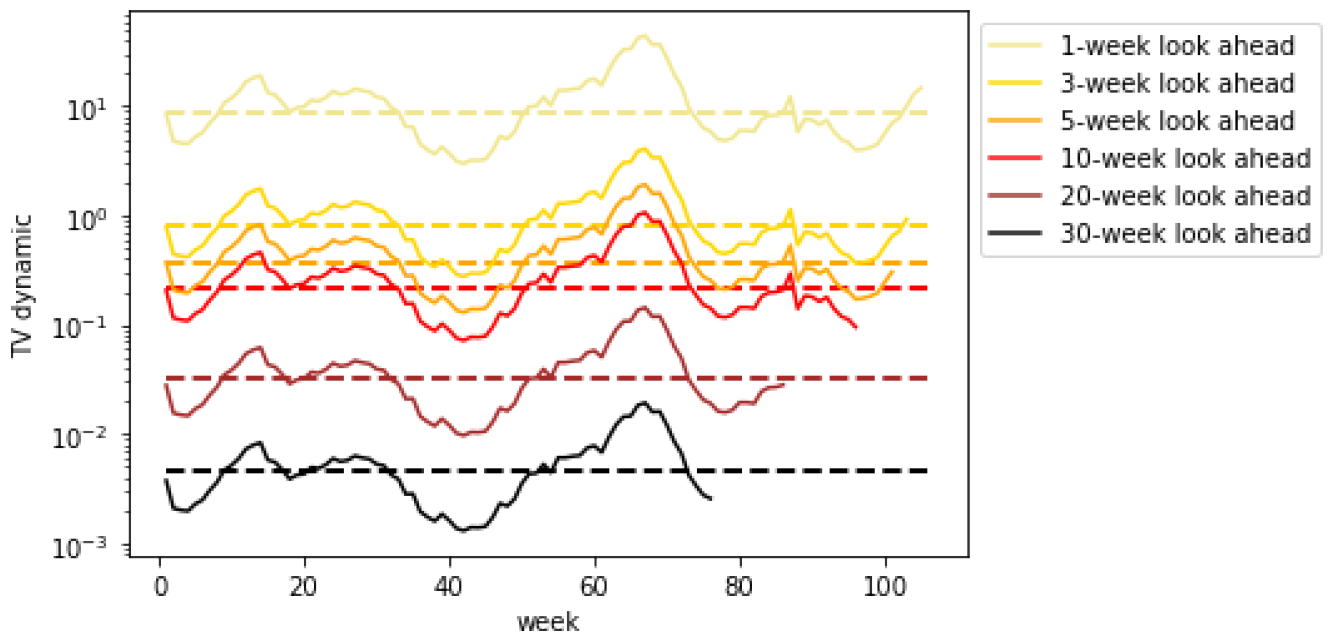
*TV* ^*dynamic*^ for weekly mortality (US excluding mobility data)

**Figure 71.**
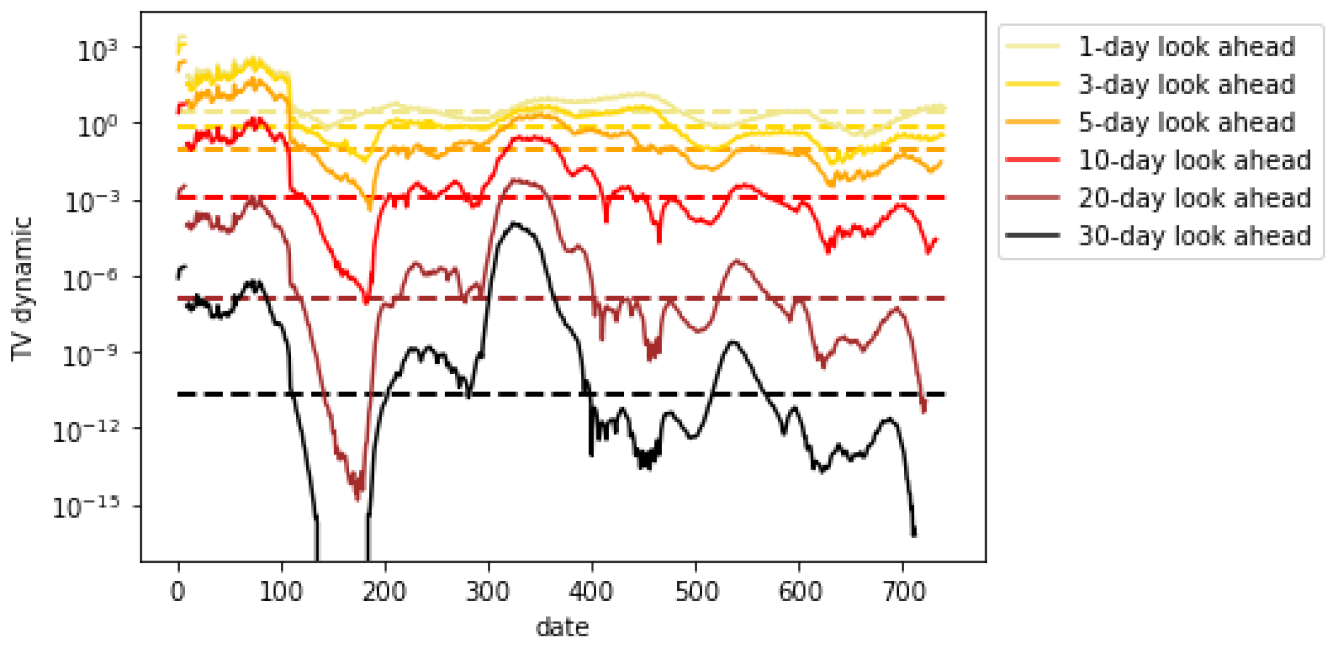
*TV* ^*dynamic*^ for daily hospitalizations (US excluding mobility data)

**Figure 72.**
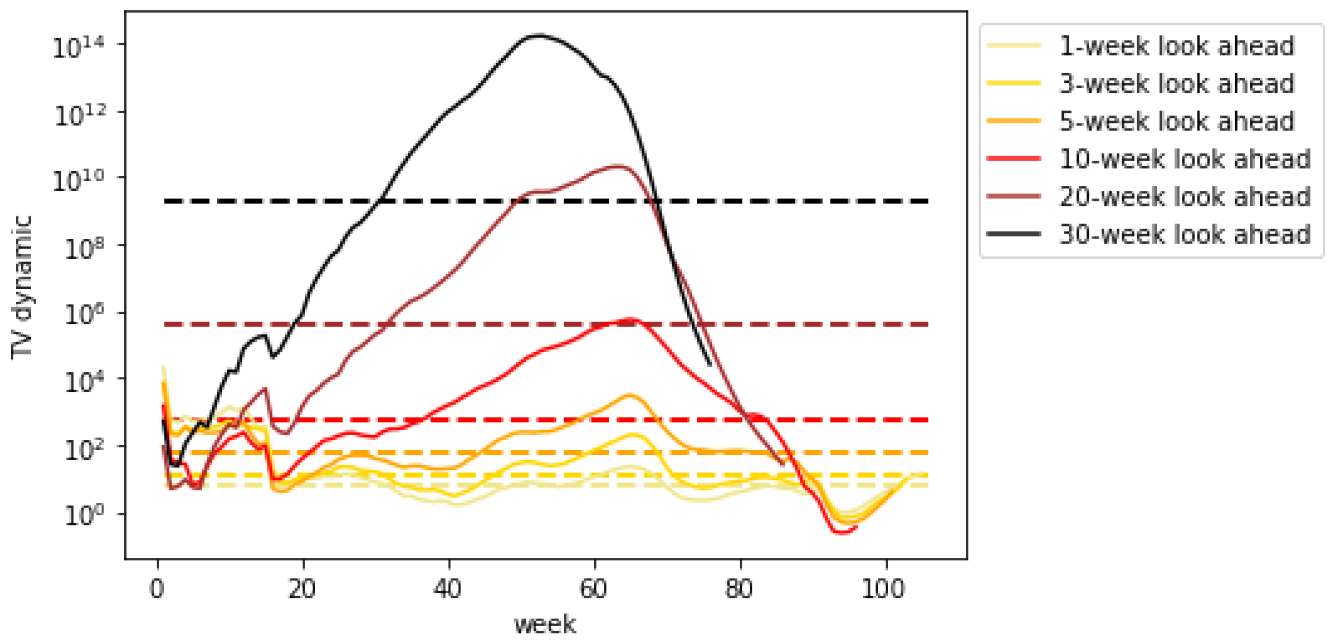
*TV* ^*dynamic*^ for weekly hospitalizations (US excluding mobility data)

